# Responsiveness to pulmonary rehabilitation in COPD is associated with changes in microbiota

**DOI:** 10.1101/2022.10.28.22281668

**Authors:** Sara Melo-Dias, Miguel Cabral, Andreia Furtado, Sara Souto-Miranda, Maria Aurora Mendes, João Cravo, Catarina R. Almeida, Alda Marques, Ana Sousa

## Abstract

**Background:** Pulmonary Rehabilitation (PR) is one of the most cost-effective therapies for chronic obstructive pulmonary disease (COPD) management. There are, however, people who do not respond to PR and reasons for non-response are mostly unknown. PR is likely to change the airway microbiota and this could play a role in its responsiveness. In this study we have explored the association between PR effectiveness and specific alterations in oral microbiota and inflammation.

**Methods:** A prospective longitudinal study was conducted. Data on exercise capacity, dyspnoea, impact of disease and 418 saliva samples were collected from 76 patients, half of whom participated in a 12-weeks PR programme. Responders and non-responders to PR (dyspnoea, exercise-capacity and impact of disease) were defined based on minimal clinically important differences.

**Results:** Changes in microbiota, including *Prevotella melaninogenica* and *Streptococcus* were observed upon PR. *Prevotella*, previously found to be depleted in severe COPD, increased during the first month of PR in responders. This increase was negatively correlated with *Streptococcus* and *Lautropia*, known to be enriched in severe cases of COPD. Simultaneously, an anti-inflammatory commensal of the respiratory tract, *Rothia,* correlated strongly and negatively with several pro-inflammatory markers, whose levels were generally boosted by PR.

Conversely, in non-responders, the observed decline in *Prevotella* correlated negatively with *Streptococcus* and *Lautropia* whose fluctuations co-occurred with several pro-inflammatory markers.

**Conclusions:** PR is associated with changes in oral microbiota. Specifically, PR increases salivary *Prevotella melaninogenica* and avoids the decline in *Rothia* and the increase in *Streptococcus* and *Lautropia* in responders, which may contribute to the benefits of PR.

**Notation of prior abstract publication/presentation:** S. Melo-Dias, M. Cabral, A. Furtado, S. Souto-Miranda, J. Cravo, M. A. Mendes, C. R. Almeida, A. Marques, <u>A. Sousa</u>, 2022, **“Responsiveness to pulmonary rehabilitation is related with changes in oral microbiota of people with COPD”** (Oral communication in 38° Congresso de Pneumologia, <u>November 2022</u>)

<u>S. Melo-Dias</u>, M. Cabral, A. Furtado, S. Souto-Miranda, J. Cravo, M. A. Mendes, C. R. Almeida, A. Marques, A. Sousa, 2022, **“Pulmonary rehabilitation changes the oral microbiota of people with COPD”** (Oral communication in <u>ERS International Congress, September 2022</u>)

<u>S. Melo-Dias</u>, M. Cabral, A. Furtado, S. Souto-Miranda, C. R. Almeida, A. Marques, A. Sousa, 2022, **“The Effectiveness of pulmonary rehabilitation in COPD is associated with specific shifts in oral microbiota”** (Poster presentation in the <u>7^th^ meeting of International Society for Evolution, Medicine and Public Health, July 2022</u>)

<u>A. Furtado</u>, S. Melo-Dias, M. Cabral, A. Marques, A. Sousa, 2021, **“The effect of pulmonary rehabilitation in salivary microbiota of people with chronic obstructive pulmonary disease: A longitudinal study”** (Poster presentation in <u>IV International Conference - “Microbiota and Health”, 22 October 2021</u>)

## Introduction

Chronic obstructive pulmonary disease (COPD) is heterogenous and complex and therefore, difficult to treat and manage. Pulmonary rehabilitation (PR) is a grade A non-pharmacological therapy and one of the most cost-effective approaches for the management of COPD [1]. Compared to pharmacological treatment, PR is three to five times more effective in improving exercise capacity, dyspnoea, and quality of life [2]. Response to PR is however multidimensional and heterogeneous, which means that for the same outcome patients are not equally responsive [3]. Reasons behind non-response are mostly unknown [3,4].

An association between the airway microbiota, including the oral microbiota [5], and disease severity has been extensively established in people with COPD [6–8]. However, the impact of PR on the airway microbiota has not yet been investigated, mainly because of the difficulty in collecting airway samples, such as sputum and bronchoalveolar lavage (BAL), routinely. Considering that only around 30% of people with COPD have productive cough [1], inducing sputum production becomes the only viable alternative. Unfortunately, this is challenging to collect outside the hospital context and requires both specialized personnel and equipment. The acquisition of BAL samples is even more challenging, as it is limited to hospital centres and bronchoscopy in patients with COPD carries a significantly higher risk of complications such as pneumonia, respiratory failure and desaturation compared with those with normal lung function [9]. In this scenario, saliva emerges as a viable non-invasive alternative to sampling the airway microbiota, which collection is simple and can be performed in different settings, namely at-home. Moreover, the oral and lower airway microbiotas are highly correlated [10], given the topological continuity between the niches, implying oral bacteria as the major colonizers of the lungs through aspiration [10–12].

An additional reason to choose saliva to evaluate the effect of PR in the airway microbiota, is the role of oral bacteria in nitrate metabolism. Exercise training, one of the main components of PR, stimulates the synthesis of nitric oxide by the human body, which is a key regulator of skeletal muscle blood flow, contractility, and mitochondrial function [13]. Oral microbiota has been strongly implicated in exercise performance, mainly due to its essential role in nitrate-nitrite-nitric oxide pathway [14,15], with recent studies showing nitrate oral supplementation to enhance PR effectiveness [16]. The oral microbiota although acknowledged as essential for this positive effect, was never studied.

On the other hand, microbiota modifications are frequently connected with an inflammatory response, which is well known to be modulated by exercise [17] and affected by PR, although with inconsistent results [18–20].

Here, we have explored the association between PR and changes in oral microbiota and inflammatory markers of people with COPD to propose that PR effectiveness could be related, at least partially, with bacterial-driven immune regulation.

## Methods

A prospective longitudinal cohort study was conducted. Ethical approvals were obtained from Administração Regional de Saúde Centro (64/2016), Centro Hospitalar do Baixo Vouga (including Estarreja’s Hospital; 08-03-17) and Agrupamento dos Centros de Saúde do Baixo Vouga. Written informed consent was obtained from all participants.

The study was reported following the “Strengthening the Reporting of Observational Studies in Epidemiology” statement [21]. Participants were identified and referenced by clinicians who briefly explained the purposes of the study. Participants were eligible if diagnosed with COPD according to the Global Initiative for Chronic Obstructive Lung Disease (GOLD) criteria [1] and stable with no acute exacerbations in the month prior to enrolment. Exclusion criteria were presence of severe cardiac, musculoskeletal, or neuromuscular diseases, signs of cognitive impairment, active neoplasia or immune diseases, that could hinder their participation in PR.

The opportunity to be integrated in the PR programme was offered to all participants. Those who did not accept to participate in the intervention but agreed with being monitored during the 5-month period were included in the control group. The intervention group (n=38) undertook a 12-week community-based PR program, whereas the control group (n=38) did not.

Sociodemographic (age, sex), anthropometric (height and weight) and general clinical (smoking habits; medication, long-term oxygen use; number of acute exacerbations and hospitalizations in the past year and comorbidities - Charlson Comorbidity Index) data were collected with a structured questionnaire. Lung function was assessed with spirometry as recommended. Exercise capacity was assessed with the six-minute walk test (6MWT), a self-paced test of walking capacity. Impact of the disease was assessed with the COPD assessment test (CAT), a 8-item questionnaire, each assessed with a 6-point Likert scale. Dyspnoea at rest was assessed with the modified Borg Scale (mBorg), a 10-item scale. Clinical data (pre-post PR for the intervention group and M0-M3 for the control group) was collected with a structured protocol adapted from the team published work [22].

Saliva samples were collected monthly with the passive drool method. Prior to sample collection, patients were advised to drink a glass of water (especially if they had recently drunk coffee or citrus juice) and to provide 3-4 mL of saliva using a labelled sample collection cup. Subsequently, the sample was transported in a cooler to the lab as quickly as possible and preserved at −80°C until DNA extraction and/or inflammatory markers’ quantification (stored at −80°C for 1 to 6 months).Response/non-response to PR was determined based on published minimal clinical importance differences (MCIDs), i.e., −1 point for mBorg [23]; 25m for 6MWT [24] and −2 points for CAT [25].

Deep sequencing of the V4 hypervariable region of 16S rRNA gene (F515/R806 primer pair) was performed for all samples. The bead assay LEGENDplex™ Human Inflammation Panel 1 (13-plex) (BioLegend, San Diego, CA, USA) was used to quantify inflammatory markers (IL-1β, IFN-α2, IFN-γ, TNF-α, MCP-1, IL-6, IL-8, IL-10, IL-12p70, IL-17A, IL-18, IL23, and IL-33) in both groups in three timepoints, M0, M1 and M3. QIIME2 2020.8 [26,27] was used to perform microbiota analyses. All statistical analyses were performed in GraphPad Prism 8 [28] and R software v 3.6.1 [29]. Longitudinal models (*lme4* package [30]) and repeated-measures correlations (*rmcorr* package [31]) were performed in R software v 3.6.1 [29]. The full description of the methodology used, including all steps of data collection, processing, and data analyses, are available in Supplementary file 1.

## Results

### Cohort characterisation

Seventy-six individuals with COPD were included in this study, 38 (29 male, 72±9y, BMI: 26±4kg/m^2^, FEV_1_pp 49.2±16 % predicted, GOLD A-8, B-20, C-0, D-10) in the intervention group and the remaining 38 patients (31 male, 70±8y, BMI: 26.4±4.8kg/m^2^, FEV_1_pp 52.3±19.8 % predicted, GOLD A-17, B-12, C-1, D-8) in the control group. Table 1 summarizes the baseline characteristics of the two groups. No significant differences were found between groups (see Table 1), and therefore, no adjustment for confounding factors was performed.

**Table 1.** Sociodemographic, anthropometric, and clinical characteristics of individuals with chronic obstructive pulmonary disease at beginning of the study (n=76).

| Characteristics | Intervention group<br>(n=38) | Control group<br>(n=38) | p-value |
| --- | --- | --- | --- |
| Age (years), mean±SD | 72±9 | 70±7.60 | 0.19<sup>1</sup> |
| Gender, n (%) |  |  | 0.57<sup>2</sup> |
| Male | 29 (76%) | 31 (82%) |  |
| Female | 9 (24%) | 7 (18%) |  |
| BMI (kg/m<sup>2</sup>), mean±SD | 26±4 | 26.4±4.8 | 0.72<sup>3</sup> |
| FEV<sub>1pp</sub>, mean±SD | 49.2±16.0 | 52.3±19.8 | 0.53<sup>1</sup> |
| FVC<sub>pp</sub>, mean±SD | 77.8±17.3 | 76.9±27.9 | 0.68<sup>1</sup> |
| Pack-years, mean±SD | 42.4±43.0 | 53.0±51.2 | 0.27<sup>1</sup> |
| Comorbidities (CCI), mean±SD | 4.2±1.4 | 3.8±1.3 | 0.10<sup>1</sup> |
| GOLD grade, n (%) |  |  | 0.22<sup>4</sup> |
| 1 | 3 (8%) | 6 (16%) |  |
| 2 | 14 (37%) | 12 (32%) |  |
| 3 | 20 (53%) | 15 (39%) |  |
| 4 | 1 (3%) | 5 (13%) |  |
| GOLD group, n (%) |  |  | 0.069<sup>4</sup> |
| A | 8 (21%) | 17 (45%) |  |
| B | 20 (53%) | 12 (32%) |  |
| C | 0 (0%) | 1 (3%) |  |
| D | 10 (26%) | 8 (21%) |  |
| Medication, n (%) |  |  |  |
| ICS | 21 (55%) | 19 (50%) | 0.8<sup>4</sup> |
| LABA | 33 (87%) | 30 (79%) | 0.36<sup>4</sup> |
| LAMA | 24 (63%) | 25 (66%) | >0.99 |
| LTRA | 2 (5%) | 2 (5%) | >0.99<sup>4</sup> |
| SABA | 9 (24%) | 5 (13%) | 0.37<sup>4</sup> |
| SAMA | 2 (5%) | 0 (5%) | 0.49<sup>4</sup> |
| Macrolides in stability | 0 (0%) | 0 (0%) | >0.99 |
| Number of exacerbations in the last year, n (%) |  |  | 0.60<sup>2</sup> |
| 0 or 1 | 27 (71%) | 29 (76%) |  |
| ≥2 or 1 with a hospitalisation | 11 (29%) | 9 (24%) |  |
| Number of hospitalisations in the last year due to COPD, n (%) |  |  | 0.67<sup>4</sup> |
| 0 | 32 (84%) | 34 (89%) |  |
| 1 | 2 (5%) | 3 (8%) |  |
| 2 | 3 (8%) | 1 (3%) |  |
| Missing values | 1 (3%) | 0 (0%) |  |
| mBORG, mean±SD | 0.8±1.3 | 0.5±1.0 | 0.3<sup>1</sup> |
| CAT, mean±SD | 17.1±8.1 | 11.7±7.4 | 0.003<sup>1</sup> |
| Walked distance in 6MWT, mean±SD | 389.0±133.9 | 457.5±82.8 | 0.045<sup>1</sup> |

SD: standard deviation, n (%): absolute and relative frequency, respectively; BMI: body mass index, FEV<sub>1pp</sub>: forced expiratory volume in 1 second percent predicted, CCI: charlson comorbidity index, GOLD: global initiative for chronic obstructive lung disease, ICS: inhaled corticosteroids, LABA: long-acting beta agonists, LAMA: long-acting muscarinic agonists, LTRA: leukotriene receptor antagonists, SABA: short-acting beta agonists, SAMA: short-acting muscarinic agonists, <sup>1</sup> p-value obtained with Mann Whitney U-test, <sup>2</sup> p-value obtained with Chi-square test, <sup>3</sup> p-value obtained with unpaired t-test with Welch's correction, <sup>4</sup> p-value obtained with Fisher's exact test.

Oral microbiota was similar between groups, being composed by two major phyla, Firmicutes (∼43%) and Bacteroidetes (∼24%), followed by Proteobacteria, Fusobacteria, Actinobacteria and ten low abundant phyla (<2%) (Supplementary figure 1).

### Changes in oral microbiota and inflammatory response are associated with PR

We followed the temporal dynamics of beta-diversity (global rate of change (grc) of Weighted-Unifrac distance) in patients for a period of five months, including the 12 weeks of PR, to understand the impact of PR in microbiota composition. As a control for steady-state microbiota fluctuations, an independent cohort of patients who did not undergo PR, was surveyed for an equivalent amount of time.

The dynamics of microbiota composition was significantly different between patients undergoing PR and controls (Figure 1A) (lmer on Weighted-Unifrac distance (grc), Group:Time-point, F= 7.58, p<0.001).

**Figure 1.**
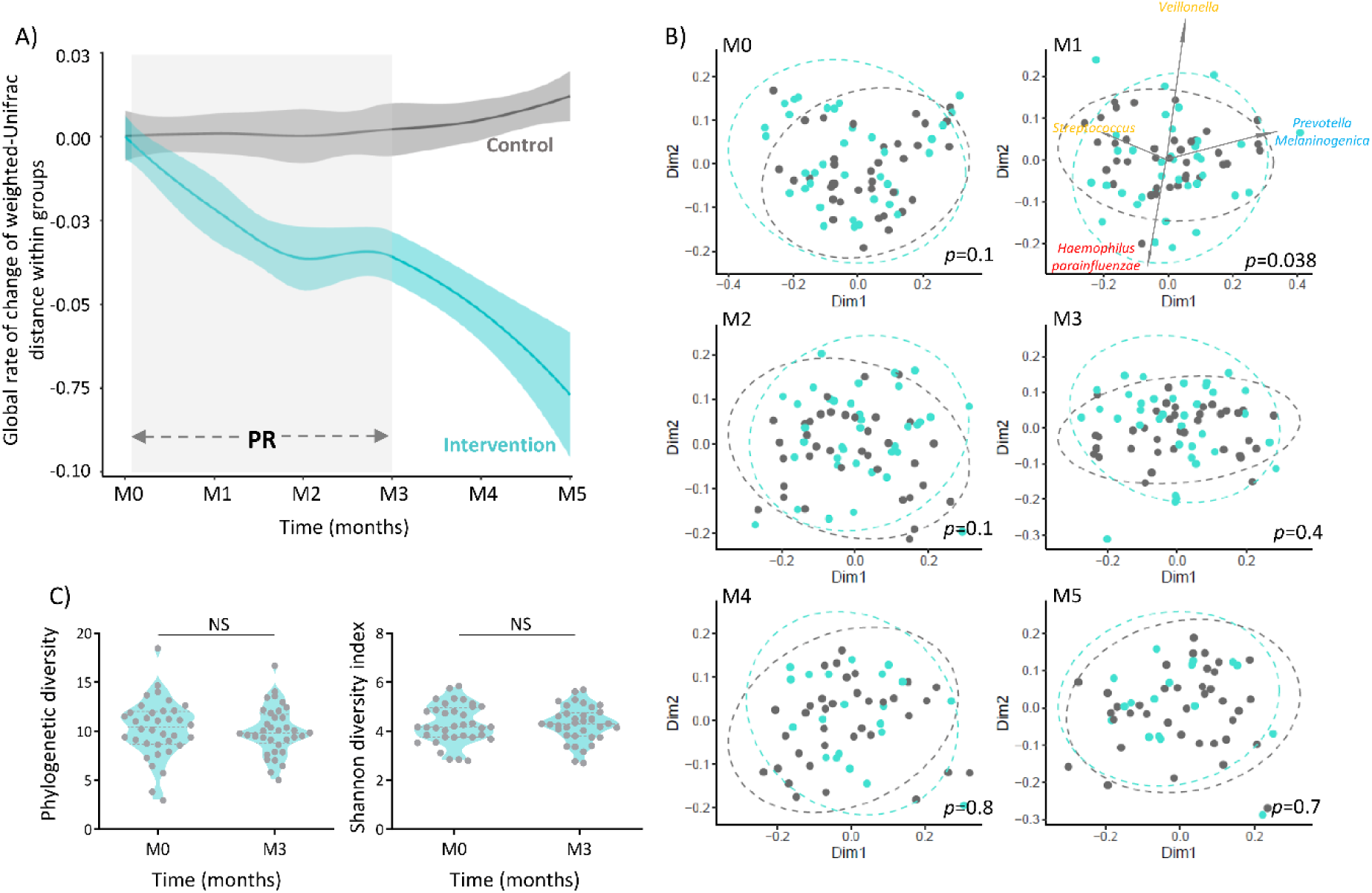
Pulmonary rehabilitation modulates the microbiota composition of people with COPD, but does not affect alpha diversity. **A)** Global rate of change of beta diversity was estimated by subtraction of baseline values at all timepoints. Blue and grey loess lines were fitted over the data points representing beta-diversity relative to baseline along time in the intervention and control groups, respectively. The blue and grey areas represent the 0.95 confidence intervals. (M0=immediately prior PR; M1, M2, M3= 1,2,3 months after initiating PR; M4, M5=1 and 2 months after finishing PR). Intervention and control group presented different microbiota dynamics over the 5-month follow-up (LMER on Weighted Unifrac distance matrix, Group:Time-Point, p<0.001). The grey rectangle indicates the time interval spanning PR. **B)** Principal coordinate analysis of Weighted Unifrac distance between groups in each timepoint. (M0, M1, M2, M3, M4 and M5). Significant differences in microbiota composition between groups were observed in M1 (PERMANOVA, F=2.27, *p*=0.038). Blue and grey dots and ellipses represent intervention and control groups, respectively. The biplot represented in M1 show the top ASVs contributing for sample segregation (*5608c3e6c9de9ceb79610e7786bd0ac4: Veillonella, d0b698c7298bf04110a6d2f220879bfb: Prevotella melaninogenica, e27680d4009f98f30248d823bc17fb8e: Haemophilus parainfluenzae, a5189f77a2cfeab3bc1602ff5c8ac3e9: Streptococcus*). **C)** No pre-post differences were observed in alpha diversity (Phylogenetic index and Shannon diversity index) in the Intervention group (Wilcoxon test: phylogenetic diversity: W= −85. p=0.48; Shannon diversity index: W=-1 p>0.99). \**p*<0.05, \*\**p*<0.01, \*\*\**p*<0.001

Principal coordinate analysis of pairwise distances (Weighted Unifrac) between groups in each timepoint showed significant differences in microbiota composition between groups upon one month of PR (Figure 1B) (PERMANOVA, F=2.3, *p*=0.038). The top four amplicon sequence variants (ASVs) responsible for individual’s separation after the first month of PR were identified as *Veillonella*, *Prevotella melaninogenica*, *Haemophilus parainfluenzae* and *Streptococcus*.

Regarding microbiota diversity within individuals (alpha diversity), no significant changes were observed upon PR (Figure 1C) (Wilcoxon test on phylogenetic diversity: W= −85. p=0.48; Wilcoxon test on Shannon diversity: W= −1. p>0.99).

Next, we compared the microbiota composition of the intervention group at baseline with that at M1, M2 and M3, to identify which taxa could have been affected by PR. PERMANOVA (M0vsM1: F=0.6, *p_adjust_*>0.99; M0vsM2: F=1.3, *p_adjust_*>0.99; M0vsM3: F=1.2, *p_adjust_*>0.99; M0vsM4: F=1, *p_adjust_*>0.99; M0vsM5: F=1.4, *p_adjust_*=>0.99), ANCOM and LEfSe detected no significant differences between pre- and post-PR microbiota composition. This is compatible with the inherent correlation among the groups (composed by the same individuals) being stronger than differences introduced by PR.

To overcome this limitation, given the longitudinal design of the experiment, we next tested for differences in taxa dynamics between the intervention and control group. This was performed using linear mixed-effect models. The temporal dynamics of the phylum Fusobacteria (lmer, Group:Time-point, F= 5.9, p=0.02) and several oral genera, such as *Fusobacterium*, *Streptococcus*, *Dialister* and *Selenomonas* were observed to be associated with PR (Table 2). Target testing for the top four ASVs distinguishing patients on M1 was also performed. Remarkably, significant alterations in the relative abundances of *Prevotella melaninogenica* and *Streptococcus*, two ASVs previously associated with disease severity in people with COPD [5], were also found (lmer, *P. melaninogenica:* Group:Time-point, F= 4.34, p=0.04; lmer, *Streprococcus:* Group:Time-point, F= 4.96, p=0.03).

**Table 2.** Summary table of significant linear mixed effect models established to assess differences in microbiota dynamics between intervention and control groups.

| OTUs/ASVs longitudinal dynamics | test's statistics | p-value (TP:Group) |
| --- | --- | --- |
| *Fusobacteria* | 5.86 | 0.016 |
| *Atopobium* | 5.51 | 0.019 |
| *Streptococcus* | 5.03 | 0.026 |
| *Dialister* | 6.39 | 0.012 |
| *Parvimonas* | 4.75 | 0.030 |
| *Fusobacterium* | 7.00 | 0.009 |
| *a5189f77a2cfeab3bc1602ff5c8ac3e9: Streptococcus* | 4.96 | 0.027 |
| *d0b698c7298bf04110a6d2f220879bfb: Prevotella melaninogenica* | 4.34 | 0.038 |
| *e27680d4009f98f30248d823bc17fb8e: Haemophilus parainfluenzae* | 0.18 | 0.673 |
| *5608c3e6c9de9ceb79610e7786bd0ac4: Veillonella* | 3.30 | 0.070 |

Models were produced based on the longitudinal relative frequencies of phyla, genera, and ASVs *a5189f77a2cfeab3bc1602ff5c8ac3e9*, *d0b698c7298bf04110a6d2f220879bfb*, *e27680d4009f98f30248d823bc17fb8e* and *5608c3e6c9de9ceb79610e7786bd0ac4*. Data upon relative frequencies was transformed with arcsine and square root transformation. TP: time-point

Considering that changes in microbiota are expected to be related with the inflammatory response, we further quantified a panel of 13 cytokines in three time points: M0, M1 and M3 in both intervention and control groups (Figure 2 and Supplementary figure 2). Significant changes in the inflammatory profile of patients undergoing PR were observed (Figure 2). Specifically, upon one month of PR (M1) significant increases in the amounts of IL-10 and IL-18 were observed (Wilcoxon signed-rank: IL-10, W=163 *p_adjust_*=0.038; IL-18, W=165 *p_adjust_*=0.035). No significant changes were observed in the same period in control group. After three months of PR (M3), TNF-α, IL-1β, IL-18, IL-10 and IL-6 were also significantly higher than at baseline (M0) (Wilcoxon signed-rank: IL-1β, W=205 *p_adjust_*=0.016; TNF-α, W=251 *p_adjust_*=0.002; IL-10, W=263 *p_adjust_*=0.0008; IL-6, W=189 *p_adjust_*=0.03; IL-18, W=233 *p_adjust_*=0.004). IL-10 and IL-6 also increased in the control group over the same time interval (Wilcoxon signed-rank: IL-10, W=225 *p_adjust_*=0.018; IL-6, W=224 *p_adjust_*=0.019), suggesting that these cytokines may be less stable.

**Figure 2.**
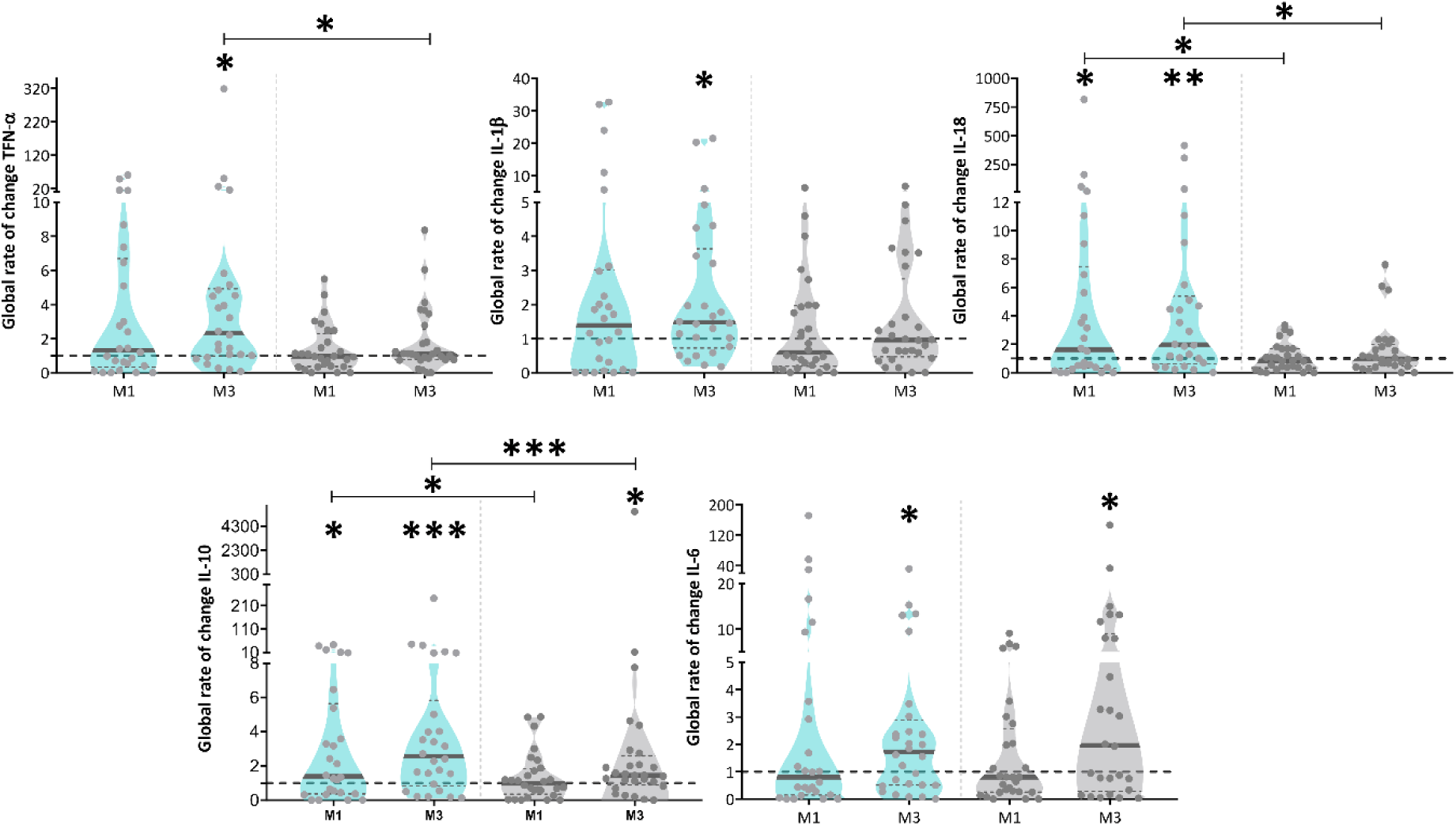
The inflammatory profile of people with COPD changes upon pulmonary rehabilitation. Global rate of change represents the ratio between cytokine values measured at baseline and M1 or M3 (*i.e.* M1/M0 and M3/M0). Differences between M1 and the baseline and between M3 and the baseline were assessed by Wilcoxon signed-rank, in each group (intervention (blue) and control (grey)). Differences between groups at M1 and M3 were assessed by Mann-Whitney U-test. p<0.05, **p<0.01, **p<0.001. All cytokines for which a significant shift was observed are represented in this figure. Supplementary figure 2 shows the data for the remaining 8 cytokines.

Regarding comparisons between the groups, shifts in IL-10 and IL-18 were significantly higher after 1 month of PR than the equivalent shifts in the control group (Mann-Whitney U-test, IL-10, U=180 *p*=0.043; IL-18, U=173 *p*=0.03). Over 3 months of PR, TNF-α, IL-18 and IL-10 had significantly higher shifts in the intervention than in the control group (Mann-Whitney U-test, TNF-α, U=246 *p*=0.04; IL-10, U=196 *p*=0.0004; IL-18, U=249 *p*=0.046). No additional significant differences were found in both groups (Supplementary figure 2).

Interestingly, the variance associated with cytokine shifts over time was generally higher in the intervention than in the control group (Supplementary table 2). This is compatible with PR generally influencing cytokine levels.

### Oral microbiota and inflammatory response vary differentially among responders and non-responders during PR

PR improved all outcomes significantly (Wilcoxon test, p < 0.05), except for BMI (W=-114, p=0.31) and mBorg (W=37, p=0.5) (Supplementary table 3). Mean improvements only exceeded the MCID for exercise capacity (6MWT, mean-diff: 45.43; Cohen’s d=0.34) (Supplementary table 2). Supplementary figure 3 represents the overlap between responders (R) and non-responders (NR) to PR in exercise capacity (6MWT), impact of disease (CAT) and dyspnoea (mBorg). From 38 patients, 24% responded in dyspnoea, 63% responded in exercise capacity and 63% responded in the impact of disease. 16% of patients responded simultaneously to all three domains and 16% failed to respond to any domain.

We analysed differences in microbiota composition among R and NR at M0 to query for a possible relationship between microbiota composition before PR and its effectiveness (Figure 3A). Principal coordinate analysis of pairwise distances (Weighted-Unifrac) showed significant differences between R and NR to dyspnoea (PERMANOVA p = 0.04) and captured 66% of total diversity (Figure 3B). Furthermore, the ASV responsible for the biggest separation between individuals in this analysis, was *P. melaninogenica*, whose frequency was below average in 75% of R. In accordance, the differential abundance analysis (LEfSe) showed that *Prevotella* (Figure 3C) was significantly enriched in NR to dyspnoea, with an effect-size of 9. No significant differences were observed for the other domains.

**Figure 3.**
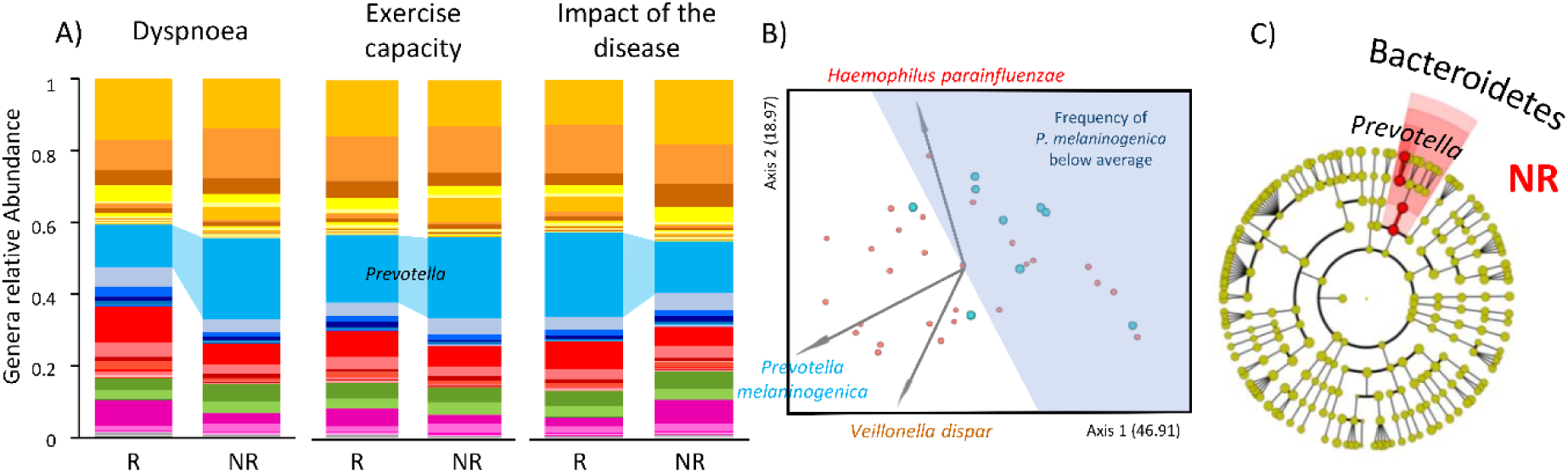
Responders and non-responders present distinct microbiota profiles prior to pulmonary rehabilitation. **A)** Mean frequency of phyla and genera of bacteria present in Responders (R) and non-responders (NR) to dyspnoea, exercise capacity and impact of the disease. **B)** R and NR to dyspnoea showed distinct microbiota composition prior to PR (PERMANOVA, p=0.04). PCoA analysis using Emperor on Weighted UniFrac distance. The biplot (grey arrows) represent the 3 most relevant ASVs for R and NR segregation. *Prevotella melaninogenica* (ASV: d0b698c7298bf04110a6d2f220879bfb) was the major contributor for segregation of R and NR followed by *Veillonella dispar* (ASV: 5608c3e6c9de9ceb79610e7786bd0ac) and *Haemophilus parainfluenzae* (ASV: e27680d4009f98f30248d823bc17fb8e). In blue is represented the area where samples presented a mean frequency of *P. melaninogenica* below the average of the dataset. 78% of R showed reduced mean frequencies of *P. melaninogenica* prior to PR while only 37% of NR followed the same trend. **C)** Cladogram highlighting differentially abundant genera between R and NR to dyspnoea prior to PR, inferred by linear discriminant analysis (LEfSe) at a significance cut-off of 3. *Prevotella* is significantly enriched in NR to dyspnoea, with an effect-size of 9.

Next, we used linear mixed effect models to compare the microbiota temporal dynamics between R and NR. Despite being differentially represented in R and NR (to dyspnoea) before PR, the frequency trajectory of *P. melaninogenica* was not significantly different among the two sets of patients during the intervention. Instead, several other taxa were found to have significantly different trajectories between the groups (Table 3 and Supplementary figure 4). Regarding the exercise capacity, the frequency trajectory of *Lautropia* (family *Burkholderiaceae*) was significantly different between R and NR (lmer, Group:Time-point, F= 2.9, p=0.02), reaching higher values in NR by the end of PR (Supplementary figure 3). The dynamics of *P. melaninogenica* was distinct among R and NR (lmer, Group:Time-point, F= 2.6, p=0.03) to impact of disease (Supplementary figure 4).

**Table 3.** Summary table of significant linear mixed effect models established to assess differences in microbiota dynamics between responders and non-responders to dyspnoea, exercise capacity and impact of the disease.

| OTUs/ASVs longitudinal dynamics | test's statistics | p-value (TP:Group) |
| --- | --- | --- |
| Response to dyspnoea during exercise (Borg Modified Scale of Dyspnoea) MCID: -1 point |  |  |
| *Spirochaetes* | 2.33 | 0.046 |
| TM7 | 5.40 | 0.000 |
| *Oribacterium* | 2.68 | 0.024 |
| *Dialister* | 2.69 | 0.023 |
| *Schwartzia* | 2.55 | 0.036 |
| [*Mogibacteriaceae*] | 2.45 | 0.036 |
| *Bulleidia* | 3.08 | 0.011 |
| *Treponema* | 2.33 | 0.046 |
| Response to exercise capacity (6-minute walk test) MCID: +25m |  |  |
| *Lautropia* | 2.87 | 0.017 |
| Response to impact of the disease (COPD assessment test) MCID: -2 points |  |  |
| *d0b698c7298bf04110a6d2f220879bfb: Prevotella melaninogenica* | 2.58 | 0.029 |

Models were produced based on the longitudinal relative frequencies of phyla, genera, *a5189f77a2cfeab3bc1602ff5c8ac3e9* and *d0b698c7298bf04110a6d2f220879bfb*. Data upon relative frequencies was transformed with arcsine and square root transformation. TP: time-point

We compared the salivary levels of cytokines present in R and NR to question whether responsiveness to PR was related with the inflammatory response. Distinct patterns of inflammatory response were observed between R and NR, in all domains (Supplementary figure 5). Significant shifts during PR were always towards values higher than the baseline and occurred mainly in NR patients. Specifically, after PR, levels of TNFα, IL-1β, IL-18 and IL-10 were higher in NR of all domains, whereas only TNF-α and IL-10 showed significant changes in R patients (please see Supplementary figure 5 for details on the statistical tests results). Interestingly, the variance in cytokine shifts after PR was also mostly higher in NR than in R (Supplementary tables 4–6).

### PR effectiveness is related with specific bacteria-inflammation correlation signatures

Independent analyses revealed an association between PR effectiveness and either changes in oral microbiota or inflammatory markers. We explored the patterns of longitudinal correlation between the two to understand in what extent PR effectiveness is linked with specific bacteria-inflammation associations. Figure 4 represents the network of significant correlations between inflammatory markers and bacterial genera inferred for the three domains. Remarkably, R and NR presented distinct patterns of bacteria-cytokine correlation (Figure 4).

**Figure 4.**
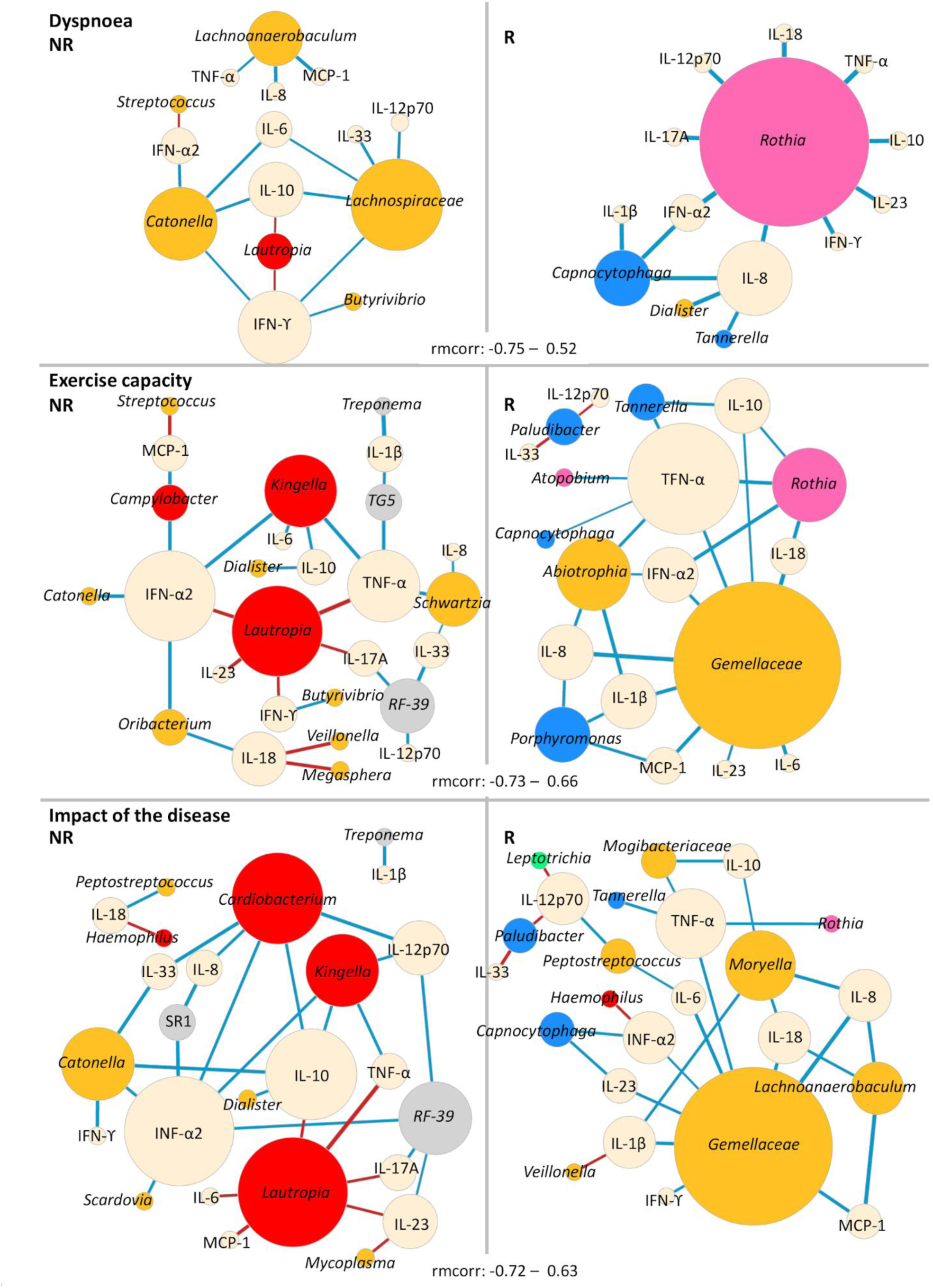
Responders and non-responders to dyspnoea, exercise capacity and impact of the disease present distinct patterns of longitudinal bacteria-inflammatory markers’ correlation. Correlation networks representing the repeated-measures correlations between bacteria and inflammatory markers in non-responders and responders to **A)** dyspnoea (rmcorr: [-0.75 – 0.52]) **B)** exercise capacity (rmcorr: [-0.73 – 0.66]) and **C)** impact of the disease (rmcorr: [-0.72 – 0.63]). Prior to the analysis, genera and ASVs: d0b698c7298bf04110a6d2f220879bfb and *a5189f77a2cfeab3bc1602ff5c8ac3e9* relative frequencies were transformed with arcsine square root transformation and inflammatory markers’ concentration was transformed with log_10_. The diameter of the nodes is proportional to the number of connections. Bacterial nodes are coloured with the colour code of each bacterial phyla (Firmicutes: yellow, Bacteroidetes: blue; Proteobacteria: red; Fusobacteria: green Actinobacteria: pink; Other phyla: grey). Inflammatory markers’ nodes are represented in beige. Positive correlations are represented in red, while negative correlations are represented in blue. The width of edges is proportional to the correlation coefficient.

In all groups of NR, *Lautropia* showed a strong positive correlation with several pro-inflammatory cytokines, being the strongest with TNF-α in NR to exercise capacity (rmcorr = 0.65, *p=*0.002) and impact of the disease (rmcorr = 0.63, *p=*0.002). Conversely, *Rothia* presented a negative correlation with several pro-inflammatory markers in all groups of R, with the strongest correlation with TNF-α (rmcorr = −0.73, *p=*0.007). Furthermore, *Gemellaceae*, was also negatively correlated with several inflammatory markers (strongest with IL-8, rmcorr = −0.69, *p<*0.0001) in R to exercise capacity and impact of the disease. Strikingly, except for a positive correlation between *Haemophilu*s and IFN-α2 in R to impact of the disease (rmcorr = 0.43, *p=*0.03), all significant correlations with Proteobacteria were found in NR.

We then performed a longitudinal correlation analysis between bacterial frequencies to gain further insight on bacterial interactions that might be related with PR responsiveness but are not necessarily linked with the immune response (Figure 5). In an effort to be conservative, since compositional data are intrinsically correlated, we chose a subset of taxa for this analysis. We included the major hubs found in the correlation between bacteria and inflammatory markers (*Lautropia*, *Rothia*, *Gemellaceae* and *Kingella*), plus five other oral taxa previously associated [5] with severity (*Prevotella*, *P. melaninogenica*, *Streptococcus*, *Streptococcus* sp., *Haemophilus* and *Porphyromonas*).

**Figure 5.**
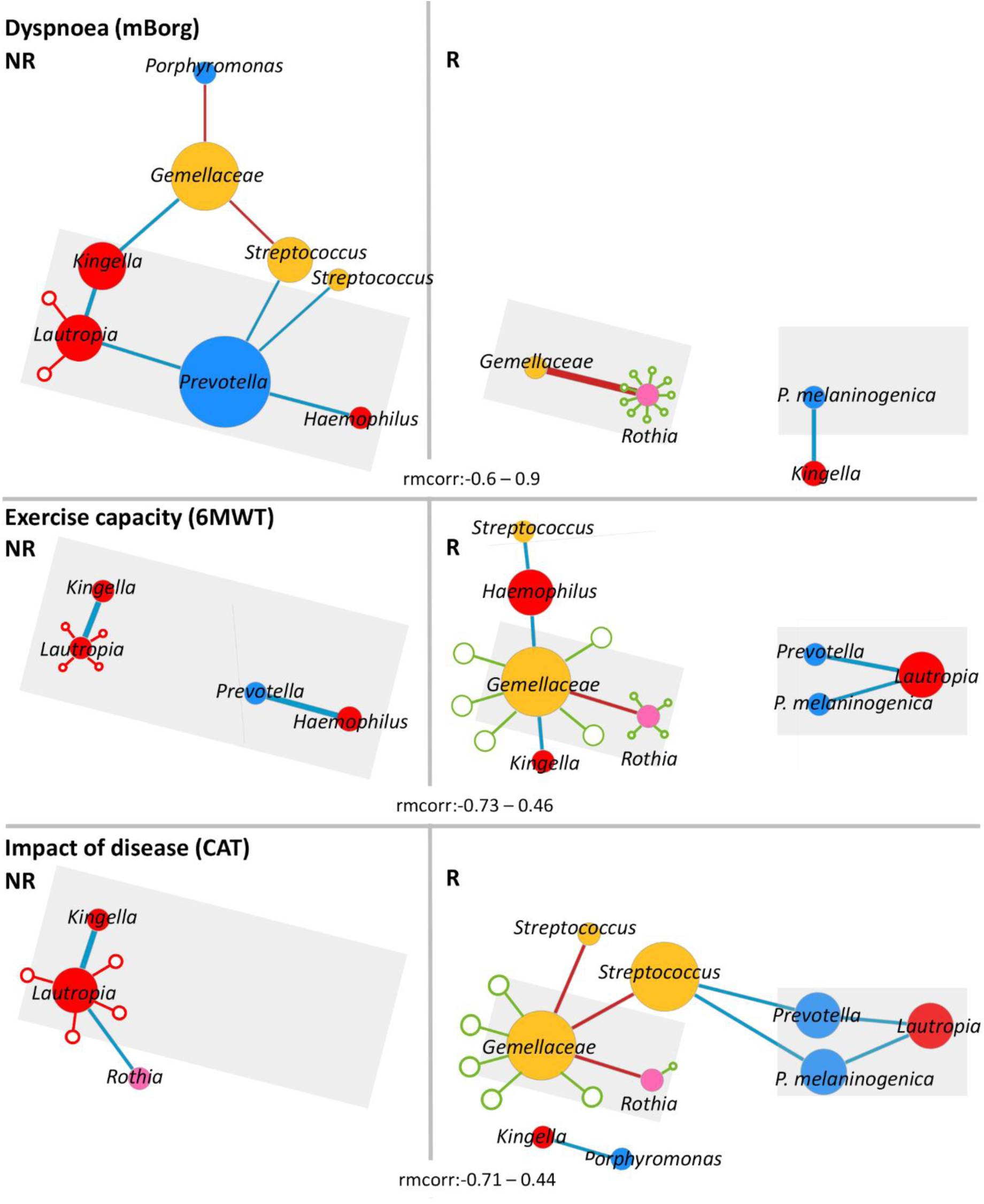
Responders and non-responders to dyspnoea, exercise capacity and impact of the disease present distinct patterns of longitudinal bacteria-bacteria correlation. Correlation networks representing the longitudinal correlations between a subset of bacterial genera and ASVs: d0b698c7298bf04110a6d2f220879bfb and *a5189f77a2cfeab3bc1602ff5c8ac3e9* non-responders and responders to **A)** dyspnoea (rmcorr: [-0.6 – 0.9]), **B)** exercise capacity (rmcorr: [-0.73 – 0.46]), and **C)** impact of the disease (rmcorr: [-0.71 – 0.44]). Grey triangles highlight common patterns of bacterial correlations between domains. Empty blue and red circles represent cytokines with which the correspondent taxon negatively and positively correlates, respectively. These correlations were inferred through the correlation analysis between bacteria-inflammatory markers and are represented here to facilitate the interpretation of the bacteria-bacteria correlation network. Prior to the analysis, genera and ASVs: d0b698c7298bf04110a6d2f220879bfb and *a5189f77a2cfeab3bc1602ff5c8ac3e9* relative frequencies were transformed with arcsine square root transformation. The diameter of the nodes is proportional to the number of connections. Bacterial nodes were coloured according to the colour code of each bacterial phyla (Firmicutes: yellow, Bacteroidetes: blue; Proteobacteria: red; Fusobacteria: green Actinobacteria: pink; Other phyla: grey). Positive correlations were represented in red, while negative correlations were represented in blue. The width of edges is proportional to the correlation coefficient.

In this analysis, *Prevotella*, whose frequency drops during PR in all NR (Supplementary figure 6 A-C), was negatively correlated with *Haemophilus* (rmcorr = −0.46, *p=*0.005), *Streptococcus* (rmcorr = −0.37, *p=*0.03) and *Lautropia* (rmcorr = −0.41, *p=*0.01) in NR to dyspnoea and with *Haemophilus* (rmcorr = −0.73, *p=*0.0004) in NR to exercise capacity. Conversely, during the first month of PR, the frequency of *P. melaninogenica* increased in all R (Supplementary figure 6A-C) and was negatively correlated with *Kingella* (R to dyspnoea, rmcorr = −0.59, *p=*0.04), *Lautropia* (R to exercise capacity, rmcorr = −0.52, *p=*0.004; R to impact of the disease, rmcorr = −0.43, *p=*0.03) and *Streptococcus* (R to impact of the disease, rmcorr = −0.41, *p=*0.04). *Gemellaceae* and *Rothia* were strongly positively correlated in all R (strongest in dyspnoea, rmcorr = 0.9, *p<*0.0001).

## Discussion

We show, for the first time, that oral microbiota changes with PR and that shifts in specific taxa appear to be strongly related with its effectiveness. By tracking two independent groups of patients for almost half a year we were able to distinguish between steady-state microbiota fluctuations and those possibly induced by PR. Contrary to the simple expectation that a well-established therapy should positively impact on patients’ microbiota, a more complex reality was unravelled. Overall, PR was not able to boost microbiota diversity within patients, a hallmark of dysbiosis, causing only low magnitude changes in a few taxa, and elicited the secretion of several pro-inflammatory cytokines.

However, when considering separately R and NR, taxa potentially related with responsiveness emerges and a differential inflammatory response becomes apparent. Previously, we have observed that low frequency of *Prevotella*, and in particular *P. melaninogenica*, was associated with a recent history of severe exacerbation [5]. Here we observed that in the first month of PR *Prevotella* was boosted in R of all domains. We hypothesize that this boost could be directly or indirectly related with the ability to respond. For example, by directly inducing the reduction of lung epithelial cell permeability (by modulating expression of tight junction proteins) [32], or indirectly by promoting lung innate immune responses, particularly against *Streptococcus pneumoniae* [33] and *Lautropia,* to which it was negatively correlated.

In fact, it has been recently observed that *P. melaninogenica* is able to promote *S. pneumoniae* rapid clearance from the lung via TLR2 activation, recruitment of neutrophils and upregulation of TNFα and IL-10 [34]. Interestingly, it was also observed that the aspiration of a mixture of 3 species of human oral commensals in mice, including *P. melaninogenica*, decreased host’s susceptibility to *S. pneumoniae* [33].

Supporting the hypothesis that, by counteracting *Lautropia*, *Prevotella* could be positively associated with PR effectiveness, is the observation that *Lautropia* is enriched in the oral microbiota of more severe cases of COPD [5] and co-occur with high levels of inflammatory markers in the eosinophilic COPD [35], an endotype that has been suggested to be predictive of worse outcome upon PR [36].

Conversely, in NR to dyspnoea and exercise capacity, the opposite scenario was observed, that is, a clear decline in *Prevotella* upon PR, negatively correlated with *Haemophilus* (exercise capacity) and with *Streptococcus* and *Lautropia* (dyspnoea). A pro-inflammatory role in COPD has been repeatedly attributed to the first two taxa [37] which could likely contribute to the lack of response to this therapy. Besides this, *Lautropia* positively correlated with several pro-inflammatory markers in all groups of NR.

Another striking observation, possibly related with the success of PR across the three domains, was the co-occurrence of *Rothia* and *Gemellacea* (in R) and its negative correlation with several pro-inflammatory markers during PR. *Rothia mucilaginosa* (the most abundant species of *Rothia* genus in our dataset) has been identified as an anti-inflammatory bacterium when present in the lungs (even in low abundance) of people with chronic respiratory diseases [38]. This has been attributed to its ability to reduce the levels of several pro-inflammatory cytokines and NF-kB activation in response to immune stimulation by pathogenic bacteria like *Pseudomonas aeruginosa* and *Staphylococcus aureus* [38].

Considering the impact of PR on individual species, this was significant on the longitudinal dynamics of two particular ASVs, *P. melaninogenica* and *Streptococcus*, previously suggested to be associated with COPD severity [5]. Moreover, the longitudinal dynamics of *P. melaninogenica* was distinct among R and NR to impact of the disease, supporting its importance to PR effectiveness.

Besides its immunomodulatory properties, *P. melaninogenica* could have an additional role in PR effectiveness, since it belongs to the most prevalent genus of oral nitrate-reducing bacteria [39]. These are essential for the nitrate-nitrite-nitric oxide pathway which is implicated in exercise performance and recovery [14,15]. Coherently, it has been shown that nitrate oral supplementation increased the beneficial effects of PR [16].

Several inconsistencies have been reported regarding the effects of PR in the inflammatory status of people with COPD. Although it is unlikely that PR increases inflammation in the long-term, our findings corroborate the previous reported increase of TNF-α in plasma of people with COPD upon PR [23]. The observed increase of IL-10 suggest an attempt to counterbalance the rise of pro-inflammatory cytokines during PR, possibly via inhibition of nuclear factor kappa B (NF-kB) and the synthesis of pro-inflammatory cytokines [24].

Furthermore, it has been observed that an inflammatory response (measured by an increase in serum levels of IL-1β, TNF-α and IL-10) is necessary for recovery from exercise-induced muscle damage [27,28]. Given the overlap between inflammatory profiles of saliva and serum [25,26], our results could reflect a similar process.

Some limitations of our study need to be acknowledged. First, the use of saliva for COPD assessment is still exploratory. Although the microbiota of upper and lower airways is highly correlated, with oral bacteria being the major colonizers of the lungs through microaspiration [10–12], more studies, besides Melo-Dias et al [5] validating its biological relevance in the context of COPD are needed. Additionally, despite longitudinal studies including PR being very resource demanding, further works with similar design should be carried out, ideally including multicentric trials with larger cohorts to evaluate the robustness of the findings. This is particularly important to assess the association between responsiveness and microbiota modulation. Second, clinical data collection was performed pre-post intervention and therefore it is not possible to adjust longitudinal models for multiple confounders such as occurrence of exacerbations, treatment with inhaled-corticosteroids and/or antibiotics.

Overall, besides responsiveness to PR being multidimensional and heterogeneous, giving rise to a moderate overlap in individuals response across domains, PR-induced changes in microbiota revealed surprisingly consistent patterns among R and NR. Furthermore, our findings suggest that PR effectiveness could be associated with a controlled inflammatory response to exercise, *i.e.*, the inflammatory response to exercise occurs accompanied by efficient regulatory mechanisms. These mechanisms can be mediated by bacteria and could be tested *in vitro*.

Future studies should address the implications and stability of these findings, clarifying the role of oral microbiota both as a biomarker of PR responsiveness and as a therapeutic target.

## Data Availability

The dataset supporting the conclusions of this article is included within the article (and its additional file(s)). Furthermore, raw sequencing data was deposited in National Centre for Biotechnology Informations (NCBI) Sequence Read Archive (SRA) (BioProject PRJNA872131).

https://www.ncbi.nlm.nih.gov/bioproject/PRJNA872131

## Author contributions

S. Melo-Dias and A. Sousa had full access to all data in the study and take responsibility for the integrity of the data and the accuracy of the data analysis. A. Marques was responsible for ethical approval. A. Marques and A. Sousa were responsible for obtaining the funding, conceiving and designing the study. M. A. Mendes, J. Cravo and S. Souto-Miranda provided a substantial contribution for clinical data acquisition. S. Melo-Dias A. Furtado conducted sample processing, DNA extraction procedures and inflammatory markers quantification. S. Melo-Dias, M. Cabral, A. Furtado, C. R. Almeida and A. Sousa conducted bioinformatics and statistical analyses. S. Melo-Dias and A. Sousa drafted the manuscript. All authors critically revised the manuscript, ensured accuracy and integrity of the work and approved the final version to be published.

## Availability of data and materials

The dataset supporting the conclusions of this article is included within the article (and its additional file(s)). Furthermore, raw sequencing data was deposited in National Centre for Biotechnology Information’s (NCBI) Sequence Read Archive (SRA) (BioProject PRJNA872131). Scripts for data analyses are provided in supplementary file 1.

## Ethics approval and consent to participate

A cross-sectional study was conducted. Ethical approvals were obtained from Administração Regional de Saúde Centro (64/2016) and from Centro Hospitalar do Baixo Vouga (08-03-17). Written informed consent was obtained from all participants.

## Consent for publication

Not applicable.

## Acknowledgements

We are grateful to all people with COPD who accepted to participate in this study and to the team of Respiratory Research and Rehabilitation Laboratory, School of Health Sciences (Lab3R-ESSUA) for their support.

## SUPPLEMENTARY MATERIAL

### Methods

#### Participants and sample collection

The pulmonary rehabilitation (PR) program was composed by twice a week 60-minute sessions of moderate exercise training and psychoeducational sessions once every other week. Detailed description of the PR program can be found elsewhere [1]. Patients were followed monthly for 5 consecutive months. Thirty-eight patients undertook a 12-week community-based PR program, intervention group, and the remaining 38 integrated the control group. In intervention group the time frame encompasses 1 month before PR, 3 months during PR and 2 months after PR. The PR program was delivered by a multidisciplinary team of healthcare professionals. During this period sociodemographic, anthropometric, clinical data were collected at M0 and M3 (corresponding to pre-post intervention) and saliva samples (monthly, passive drool method) were collected using a structured protocol [1].

Sociodemographic (age, sex, educational level), anthropometric (weight and height to compute body mass index), clinical (smoking habits, number of exacerbations and hospitalizations in the past year, past 3 months and past month, medication used, long-term oxygen, comorbidities - Charlson Comorbidity Index [2], level of airway obstruction-spirometry (FEV_1_, FVC, FEV_1_pp) (MicroLab 3535, CareFusion, Kent, UK) [3,4], medication including long term oxygen therapy, impact of the disease – COPD Assessment Test (CAT) [5,6], exercise capacity – six-minute walk test (6MWT) [7,8] data were collected following a published structured protocol of the team [1]. Dyspnoea at rest was assessed with the modified Borg Scale (mBorg) which is a 10-item scale. GOLD grades were defined according to FEV1 percentage predicted for each individual. GOLD groups were defined combining the number of exacerbations and hospital admissions of each patient in the year before enrolment with their CAT scores. From the initially predicted 456 samples a total of 418 saliva samples were collected, the number of samples collected per group and timepoint can be found in supplementary table 1. That is, our study has a 9% rate of missing saliva samples, particularly in the last timepoints M4 and M5. This can be largely explained by difficulties in reaching patients and keeping them motivated to collaborate in data collection for a long period.

##### Response to PR

Response/non-response to PR was determined with the published minimal clinical importance differences (MCIDs) for the modified Borg scale-Dyspnoea (mBorg), 6MWT and COPD assessment test (CAT) were: −1 point [9], 25m [10] and −2 points [11], respectively.

#### DNA extraction

Prior to DNA extraction, samples were thawed at room temperature and centrifuged at 10,000xg for 10 minutes. Supernatants were saved for subsequent quantification of inflammatory markers and pellets were used for DNA extraction following QIAamp DNA Mini Kit (Qiagen, Hilden, Germany) protocol with minor modifications: initial sample volume was set to 400µL and the volumes of buffers and Qiagen protease were adjusted. Elution volume was reduced to a quarter of the recommended. Thirty-eight negative controls where saliva was replaced by phosphate-buffered saline were performed in order to control for background bacterial contamination. Quality and quantity of the extracted DNA was assessed in Denovix DS-11 spectrophotometer, with OD260/280 and OD260/230 ratios.

#### 16S rRNA gene amplification and sequencing

16S rRNA gene amplification and sequencing was carried out at the Gene Expression Unit from Instituto Gulbenkian de Ciência following the implemented protocol. Briefly, for each sample, the hypervariable V4 region of 16S rRNA gene was amplified, using universal pair of primers F515 (5′-CACGGTCGKCGGCGCCATT-3′) / R806 (5′-GGACTACHVGGGTWTCTAAT-3′). Samples were then pair-end-sequenced on an Illumina MiSeq Benchtop Sequencer, following Illumina recommendations.

#### Quantification of inflammatory markers in saliva samples

The bead assay LEGENDplex™ Human Inflammation Panel 1 (13-plex) with V-bottom Plate (BioLegend, San Diego, CA, USA), previously used successfully with saliva samples [12,13], was used to quantify inflammatory markers (IL-1β, IFN-α2, IFN-γ, TNF-α, MCP-1, IL-6, IL-8, IL-10, IL-12p70, IL-17A, IL-18, IL-23, and IL-33) in oral samples from intervention and control groups.. Specifically, the supernatants obtained after sample centrifugation for 10 min at 10.000g, from the intervention (at baseline (M0), after 1 month of PR (M1), and after 3 months of PR (M3)) and control groups (in the timepoints M0, M1 and M3), were used for cytokine quantification following manufacturer’s recommendations [14]. Data acquisition was performed with BD AccuriTM C6 Plus flow cytometer and analysed with online version of LEGENDplex™ Data Analysis Software Suite [14]. For values below the detection limit an estimate was obtained using the standard curve. For values above the threshold of detection, the value corresponding to the maximum detection limit was used to replace cytokine values.

#### Microbiota, inflammatory markers, and statistical analyses

##### Sample characterisation (relative to Table 1)

Descriptive statistics was used to characterize clinical date of intervention and control groups at baseline. Data normality was assessed with Shapiro-Wilk and D’Agostino-Pearson omnibus normality tests, ensuring the assumptions of parametric statistics approach. Comparisons between intervention and control group were conducted with unpaired t-test with Welch’s correction, when quantitative data followed normal distribution, Mann-Whitney U-test, when quantitative data violated the assumptions of parametric tests and Chi-square test, when qualitative data was considered (statistical analyses were performed in GraphPad Prism 8 [15] and R software v 3.6.1 [16]). Statistical significance was considered for p-values below 0.05.

##### Analysis of illumina paired-end reads

QIIME2 2020.8 [11,17] was used to perform microbiota analyses. Demultiplexed 16s paired-end sequences were imported and q2-vsearch plugin [18] was applied to join forward and reverse reads. Quality control was assessed via q-score base filtering, chimera removing and 16S-denoising with Deblur [19,20]. Next, to exclude bacterial background contaminations, we used the *DECONTAM* package [21,22] from *R* software [16] with the prevalence method and a threshold of 0.5, meaning that ASVs were classified as contaminants if present in a higher fraction of negative controls than true samples. This allowed us to identify 235 contaminant ASVs (available in supplementary file 2), that together with ASVs from *mitochondria*, *chloroplasts* and *cyanobacteria*were removed from the dataset prior to conducting subsequent analyses. Results from previous steps were summarized in a feature table. Q2-phylogeny plugin [23] was next employed to produce a MAFFT alignment [24] of ASVs which was later used to construct a rooted phylogeny with FastTree2 [25] for subsequent applications.

Taxonomy assignment of ASVs was performed with q2-feature-classifier plugin [26,27], through classify-sklearn method with pre-trained Naïve Bayes classifier against 99%-eHOMD_v15.1 reference database [28] (sequences trimmed to only include 250bp of V4 region, bound by the F515/R806 primer pair).

Categorized samples’ metadata used for analyses is supplied in supplementary file 3. Subsequent analyses were performed upon ASVs. Differential abundance analyses were conducted with on both ASVs and OTUs at taxonomic level 6.

##### Diversity analyses

Alpha- and beta-diversities were estimated with q2-diversity plugin [29] after rarefaction to 8000 sequences per sample (subsample without replacement). Spatial dissimilarities between bacterial communities of different groups and/or periods were assessed with principal coordinate analyses (PCoA) and biplots based on Weighted Unifrac distance. Wilcoxon test and Friedman’s test with Dunn’s correction were employed to compare alpha diversity among groups, periods and/or timepoints (statistical analyses were performed in *GraphPad Prism 8* [15] and *R stats* package [30] from *R* software [16]). Statistical significance was considered for p-values below 0.05.

Differences in beta-diversity between groups, periods and/or time points were quantified by permutational multivariate analysis of variance (PERMANOVA) [31] conducted with *adonis2* function [32] (*vegan* package [33] from *R* software [16]). For all statistical analyses a p-value of ≤0.05, corrected for multiple testing whenever necessary, was considered statistically significant.

##### Differential abundance analyses of OTUs

Analysis of composition of microbiomes (ANCOM) [34] and Linear discriminant effect size (LEfSe) analysis [35] were performed to identify differentially abundant OTUs between groups, periods and/or time points. LEfSe is an algorithm for high-dimensional biomarker discovery that uses linear discriminant analysis to estimate the effect size of each taxon and does not account for the compositional nature of the microbiota. ANCOM uses a log ratio analysis to make point estimates of the variance and mean, taking into consideration the compositional nature of the data. These analyses were conducted with the feature table collapsed at genus taxonomic level (L6). LEfSe was performed in the online version [36] with an Linear Discriminant Analysis score of 3 for significance. ANCOM was performed in *R* software with *ANCOM 2.0* script [37] with taxa-wise multiple correction and a W cut-off of significance of 0.7, both recommended by the developer, based on simulation data.

##### Longitudinal analyses (linear mixed-effects models)

• ***Longitudinal differential abundance analyses of OTUs with ANCOM II***

Longitudinal differential abundance analyses were carried out between patients undergoing PR and controls, or R and NR to exercise capacity, dyspnoea and impact of the disease, with analysis of composition of microbiomes (ANCOM II) [38]. Similar to classic ANCOM, ANCOM II uses a log ratio analysis to make point estimates of the variance and mean [39], taking into consideration the compositional nature of the data, but comprises an additional step to deal with different types of zeros, which was further described by Kaul et al [38]. These analyses were conducted with the feature table collapsed at genus taxonomic level (L6) as well as with the feature table not collapsed, e.g., using only ASVs. ANCOM II was performed in R software [16] using ANCOM 2.1 script [40] with taxa-wise multiple correction and a W cut-off of 0.7 significance (both recommended by the developer, based on simulation data).

• ***Data normalization and transformation***

To evaluate the global rate of change of beta diversity of patients undergoing PR and controls, all the timepoints of both groups were normalized through subtraction of baseline values.

- Differences in the longitudinal dynamics of OTUs/ASVs’ frequencies between groups and between R and NR to exercise capacity, dyspnoea, and impact of the disease were assessed based on the arcsine square root transformed relative frequencies in each timepoint. Additionally, raw data was filtered so that the analysis included only ASVs/OTUs that were present in ≥20% of the samples. *Loess Lines*

Loess Lines plots were produced to observe dissimilar tendencies during the 5 consecutive months (M0 to M5) of the study between patients undergoing PR and controls or R and NR to exercise capacity, dyspnoea and impact of the disease. In brief, Loess regressions were fitted over longitudinal data beta-diversity and ASVs/OTUs’ relative frequencies and then plotted with ggplot2 package from R software [16].

• ***Linear mixed-effects models***

Longitudinal analyses with linear mixed-effects models, were performed to compare the longitudinal dynamics of beta-diversity (Weighted Unifrac distances) and relative frequencies of genera and 4 ASVs (top 4 ASVs responsible for patients’ separation in principal coordinate analysis), between patients undergoing PR and controls. Differences in the longitudinal trajectories of genera/ASVs’ relative frequencies between R and NR to exercise capacity, dyspnoea and impact of the disease were also evaluated. Regarding the models for beta diversity, independent variables included the timepoints (5 months) and the experimental design groups (intervention and control). Subjects were incorporated in the model as random factors, using *lmer* function of the *lme4* package [41] of R software [16]. P-values were obtained using the *Anova* function from *R-stats* package [30] and statistically significant results were considered valid if the assumptions of the model were validated.

To assess the effect of PR in the dynamics of each OTU/ASVs, the arcsine square root transformed frequencies of each OTU/ASV were defined as dependent variable in the correspondent model. Moreover, the experimental design groups (intervention and control or R and NR to exercise capacity, dyspnoea and impact of the disease) and timepoints were set as independent variables. Subjects were adjusted as a random factor. P-values were obtained using the *Anova* function and statistically significant results were considered valid if the assumptions of the model were validated. Additionally, contrast analysis with the Bonferroni correction was executed to identify the timepoints in which the mean abundance of relevant OTUs/ASVs is significantly different between R and NR to the several domains. Statistical significance was considered for p-values below 0.05.

##### Analyses of inflammatory markers

To assess changes of each inflammatory marker, the ratio between each timepoint and the baseline (M1/M0 and M3/M0) was estimated. Wilcoxon Signed-Ranks Test (GraphPad Prism 8) with post-hoc false discovery rate (FDR) correction from multiple comparisons was applied to compare ratios at M1 and M3 to baseline in all experimental groups. Mann-Whitney U-test was used to assess the differences in the median ratio of each inflammatory marker per time-point (M1 vs M1 and M3 vs M3, respectively), among the experimental groups. Z-test for variance (*PairedData* package [42] from *R* software [16]) were executed to assess differences in the variance of ratios of each inflammatory marker per time-point (M1 vs M1 and M3 vs M3, respectively), among the experimental groups. Statistical significance was considered for *p-values* below 0.05.

##### Repeated Measures Correlations

Repeated measures correlations were performed to assess the longitudinal co-variation of the frequency of genera/ASVs and inflammatory markers during PR, in the groups of R and NR to exercise capacity, dyspnoea and impact of the disease. Correlations were computed with Rmcorr function from *rmcorr* package [43] (R software [16]). Prior to correlation analyses, estimated values of inflammatory markers were transformed with log_10_ transformation, OTUs list was filtered to consider only OTUs that were present in at least 20% of the samples. Furthermore, the filtered dataset was transformed with arcsine square root transformation. Patients with only two longitudinal measurements, M0 and M3, were also included in the analyses. Statistical significance was considered for p-values below 0.05, however results were only accepted as valid if the assumptions of the *rmcorr* model were validated (namely, model residuals normally distributed and centered in zero). Valid correlations, according to previous criteria, were then plotted in correlation network plots with *igraph* package ([44] from R software [16], with node diameter proportional to the number of correlations and the edges’ width proportional to the correlation strength.

The same analysis (including timepoints M0, M1 and M3) with a subset of bacterial genera and ASVs was carried out to assess bacterial interactions that could be related with PR responsiveness. Since compositional data is intrinsically correlated the subgroup of bacterial genera/ASVs included the major hubs found in the correlation between bacteria and inflammatory markers (*Lautropia*, *Rothia*, *Gemellaceae* and *Kingella*) and six other oral taxa previously associated with severity [45] (*Prevotella*, *P. melaninogenica*, *Streptococcus*, *Streptococcus* sp., *Haemophilus* and *Porphyromonas*).

### Supplementary Tables

**Supplementary Table 1.** Number of saliva samples collected per group in each timepoint over the 5-month period.

| Time-points | Intervention<br>(n=38) | Control<br>(n=38) |
| --- | --- | --- |
| M0 | 38 | 38 |
| M1 | 37 | 38 |
| M2 | 38 | 38 |
| M3 | 37 | 38 |
| M4 | 22 | 38 |
| M5 | 18 | 38 |
| Total | 190 | 228 |

**Supplementary Table 2.**
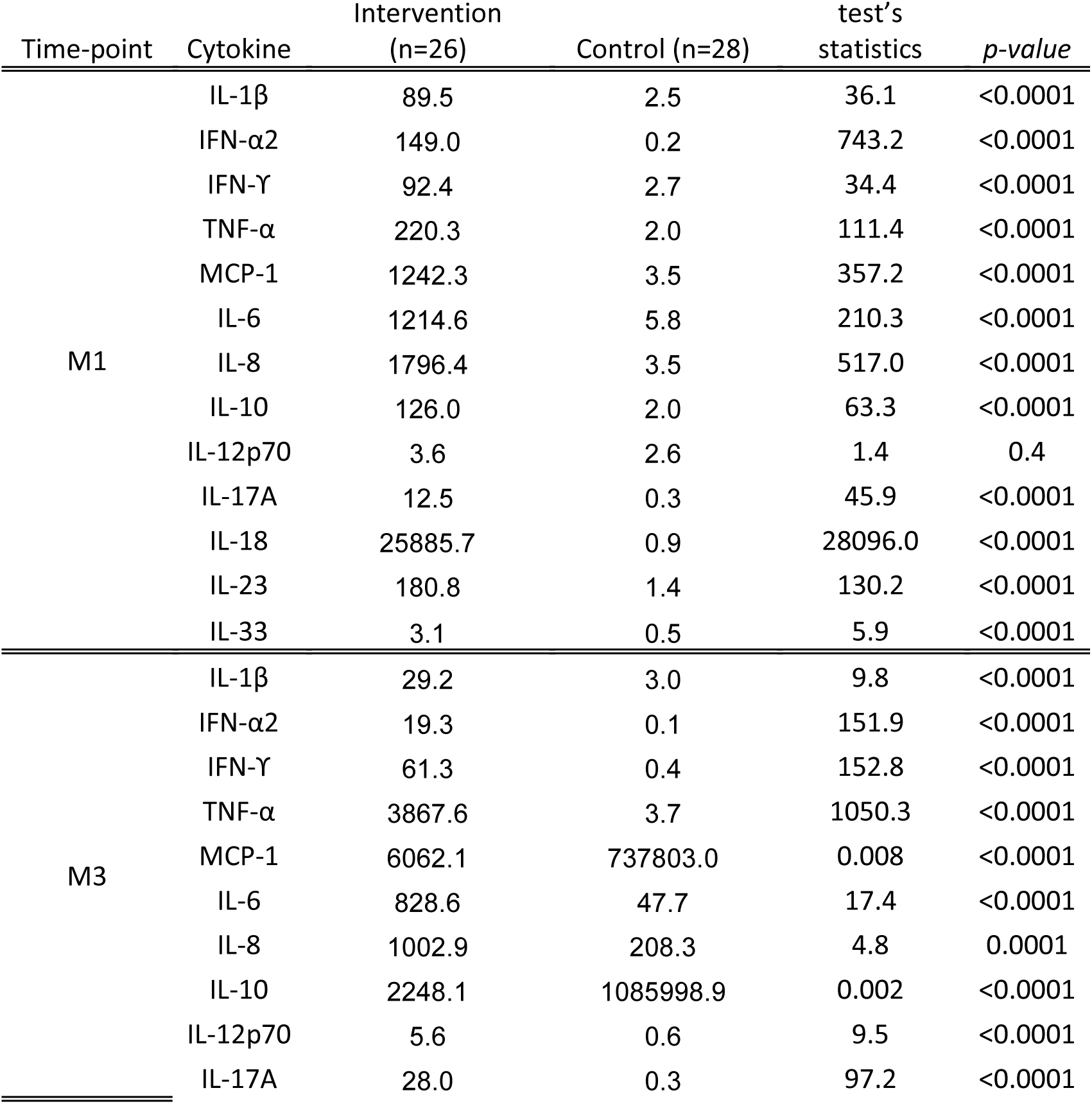
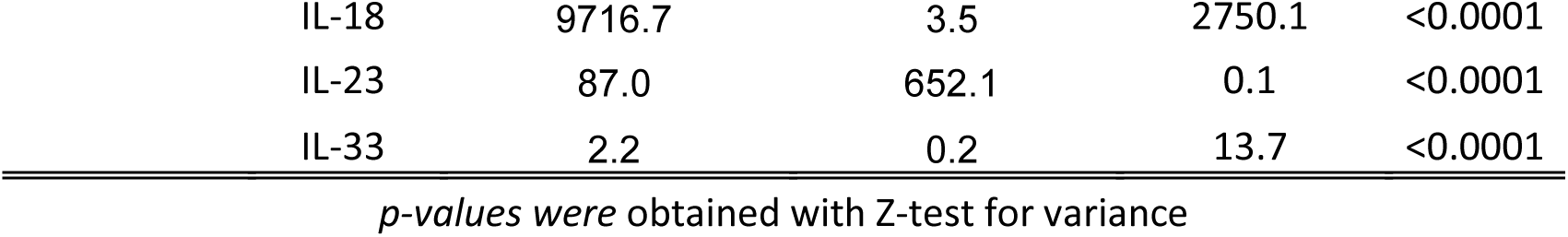
Variance of the global rate of change estimated for each cytokine at M1 and M3 in the intervention and control groups. Z-test for variance was performed to assess significant differences between the groups.

**Supplementary Table 3.** Summary of the effects of pulmonary rehabilitation in people with chronic obstructive pulmonary disease (n=38)

| Clinical Parameters | mean Pre<br>(n=38) | SD | mean Post<br>(n=38) | SD | p-value | Mean<br>Difference | Cohen's D |
| --- | --- | --- | --- | --- | --- | --- | --- |
| BMI | 26.03 | 4.3 | 25.8 | 4.1 | 0.07 | -0.28 | 0.31 |
| mBorg | 0.79 | 1.2 | 1.1 | 1.7 | 0.5 | 0.28 | 0.18 |
| CAT | 17.08 | 8.03 | 13.5 | 7.4 | 0.001 | -3.6 | 0.46 |
| 6MWT: walked distance in m | 389 | 132.2 | 434.4 | 134.2 | 0.0006 | 45.4 | 0.34 |

BMI: Body Mass Index; mBorg: modified Borg Scale of Dyspnoea; CAT: COPD assessment test; 6MWT: six-minute walk test

**Supplementary Table 4.**
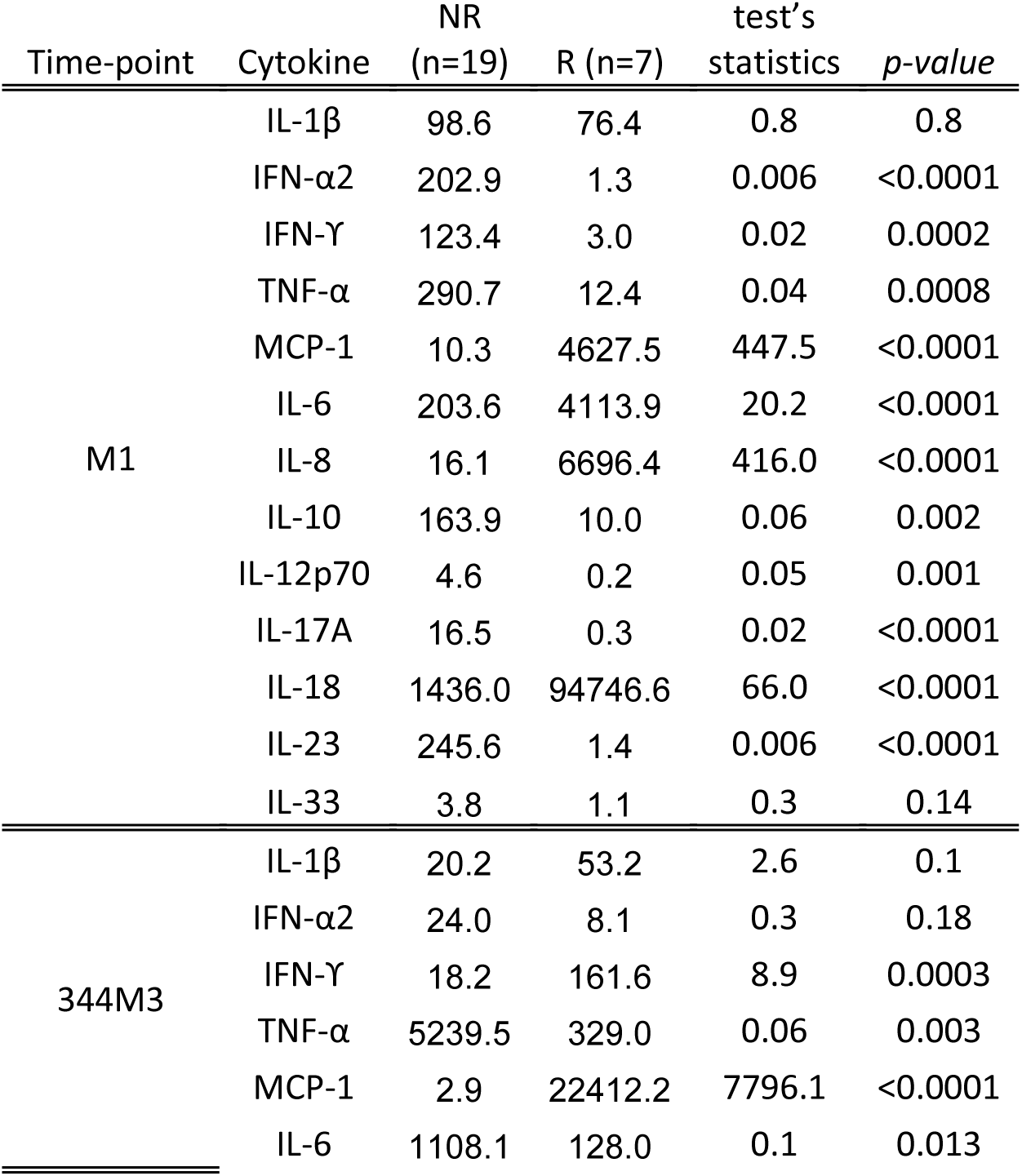
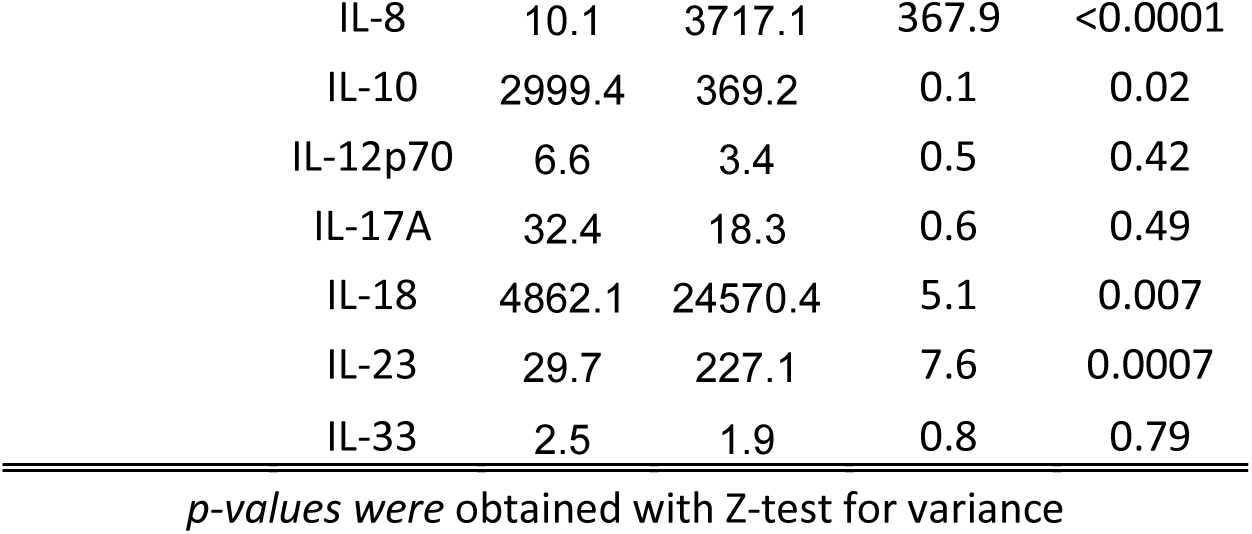
Variance of the global rate of change estimated for each cytokine at M1 and M3 in R and NR to dyspnoea (mBorg). Z-test for variance was performed to assess significant differences between R and NR.

| Time-point | Cytokine | NR<br>(n=19) | R (n=7) | test's<br>statistics | p-value |
| --- | --- | --- | --- | --- | --- |
| M1 | IL-1β | 98.6 | 76.4 | 0.8 | 0.8 |
| | IFN-α2 | 202.9 | 1.3 | 0.006 | <0.0001 |
| | IFN-γ | 123.4 | 3.0 | 0.02 | 0.0002 |
| | TNF-α | 290.7 | 12.4 | 0.04 | 0.0008 |
|  | MCP-1 | 10.3 | 4627.5 | 447.5 | <0.0001 |
|  | IL-6 | 203.6 | 4113.9 | 20.2 | <0.0001 |
|  | IL-8 | 16.1 | 6696.4 | 416.0 | <0.0001 |
|  | IL-10 | 163.9 | 10.0 | 0.06 | 0.002 |
|  | IL-12p70 | 4.6 | 0.2 | 0.05 | 0.001 |
|  | IL-17A | 16.5 | 0.3 | 0.02 | <0.0001 |
|  | IL-18 | 1436.0 | 94746.6 | 66.0 | <0.0001 |
|  | IL-23 | 245.6 | 1.4 | 0.006 | <0.0001 |
|  | IL-33 | 3.8 | 1.1 | 0.3 | 0.14 |
| 344M3 | IL-1β | 20.2 | 53.2 | 2.6 | 0.1 |
| | IFN-α2 | 24.0 | 8.1 | 0.3 | 0.18 |
| | IFN-γ | 18.2 | 161.6 | 8.9 | 0.0003 |
| | TNF-α | 5239.5 | 329.0 | 0.06 | 0.003 |
|  | MCP-1 | 2.9 | 22412.2 | 7796.1 | <0.0001 |
|  | IL-6 | 1108.1 | 128.0 | 0.1 | 0.013 |

|  |  |  |  |  |
| --- | --- | --- | --- | --- |
| IL-8 | 10.1 | 3717.1 | 367.9 | <0.0001 |
| IL-10 | 2999.4 | 369.2 | 0.1 | 0.02 |
| IL-12p70 | 6.6 | 3.4 | 0.5 | 0.42 |
| IL-17A | 32.4 | 18.3 | 0.6 | 0.49 |
| IL-18 | 4862.1 | 24570.4 | 5.1 | 0.007 |
| IL-23 | 29.7 | 227.1 | 7.6 | 0.0007 |
| IL-33 | 2.5 | 1.9 | 0.8 | 0.79 |

*p-values were obtained with Z-test for variance*

**Supplementary Table 5.**
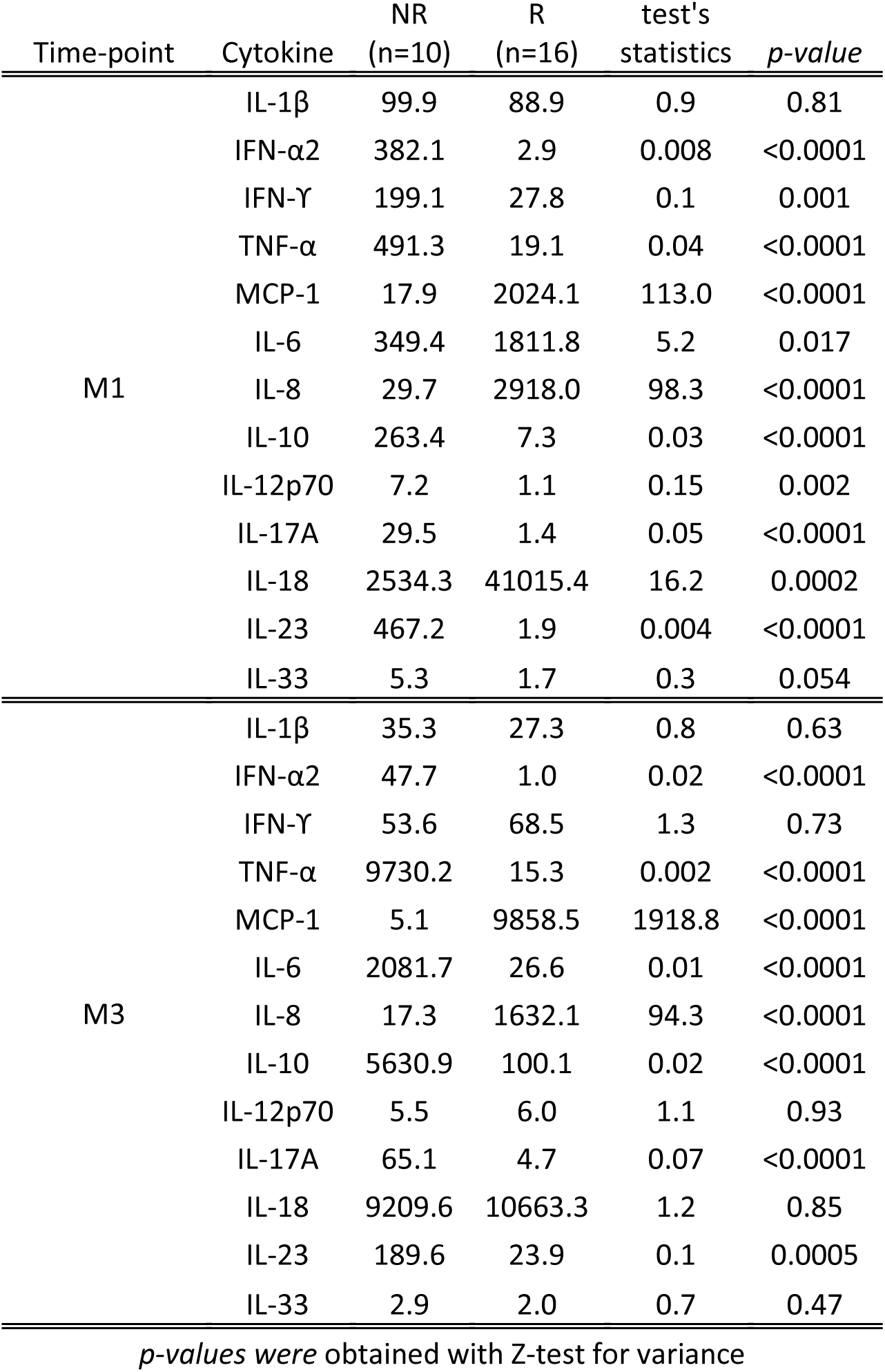
Variance of the global rate of change estimated for each cytokine at M1 and M3 in R and NR to exercise capacity (6MWT). Z-test for variance was performed to assess significant differences between R and NR.

**Supplementary Table 6.** Variance of the global rate of change estimated for each cytokine at M1 and M3 in R and NR to the impact of disease (CAT). Z-test for variance was performed to assess significant differences between R and NR.

| Time-point | Cytokine | NR<br>(n=12) | R<br>(n=14) | test's<br>statistics | *p-value* |
| --- | --- | --- | --- | --- | --- |
| M1 | IL-1β | 47.3 | 131.1 | 2.8 | 0.099 |
| | IFN-α2 | 1.0 | 276.3 | 287.0 | <0.0001 |
| | IFN-γ | 3.0 | 168.7 | 56.2 | <0.0001 |
| | TNF-α | 19.0 | 385.9 | 20.3 | <0.0001 |
|  | MCP-1 | 2692.6 | 14.0 | 0.005 | <0.0001 |
|  | IL-6 | 2401.8 | 232.5 | 0.1 | 0.0002 |
|  | IL-8 | 3883.9 | 22.1 | 0.006 | <0.0001 |
|  | IL-10 | 40.3 | 105.9 | 5.1 | 0.01 |
|  | IL-12p70 | 0.9 | 6.1 | 6.4 | 0.004 |
|  | IL-17A | 0.5 | 22.5 | 45.2 | <0.0001 |
|  | IL-18 | 54805.0 | 1964.3 | 0.04 | <0.0001 |
|  | IL-23 | 2.1 | 337.0 | 164.2 | <0.0001 |
|  | IL-33 | 1.9 | 4.4 | 2.4 | 0.16 |
| M3 | IL-1β | 33.0 | 27.7 | 0.8 | 0.76 |
| | IFN-α2 | 5.6 | 32.0 | 5.7 | 0.007 |
| | IFN-γ | 31.5 | 90.9 | 2.9 | 0.09 |
| | TNF-α | 227.0 | 7097.8 | 31.3 | <0.0001 |
|  | MCP-1 | 13133.7 | 3.5 | 0.0003 | <0.0001 |
|  | IL-6 | 90.2 | 1473.0 | 16.3 | <0.0001 |
|  | IL-8 | 2174.3 | 13.2 | 0.006 | <0.0001 |
|  | IL-10 | 160.9 | 4086.2 | 25.4 | <0.0001 |
|  | IL-12p70 | 5.1 | 6.4 | 1.3 | 0.71 |
|  | IL-17A | 9.1 | 45.5 | 5.0 | 0.01 |
|  | IL-18 | 14073.7 | 6639.2 | 0.5 | 0.2 |
|  | IL-23 | 124.2 | 61.6 | 0.5 | 0.23 |
|  | IL-33 | 2.8 | 1.9 | 0.7 | 0.48 |

*p-values* were obtained with Z-test for variance

### Supplementary Figures

**Supplementary Figure 1.**
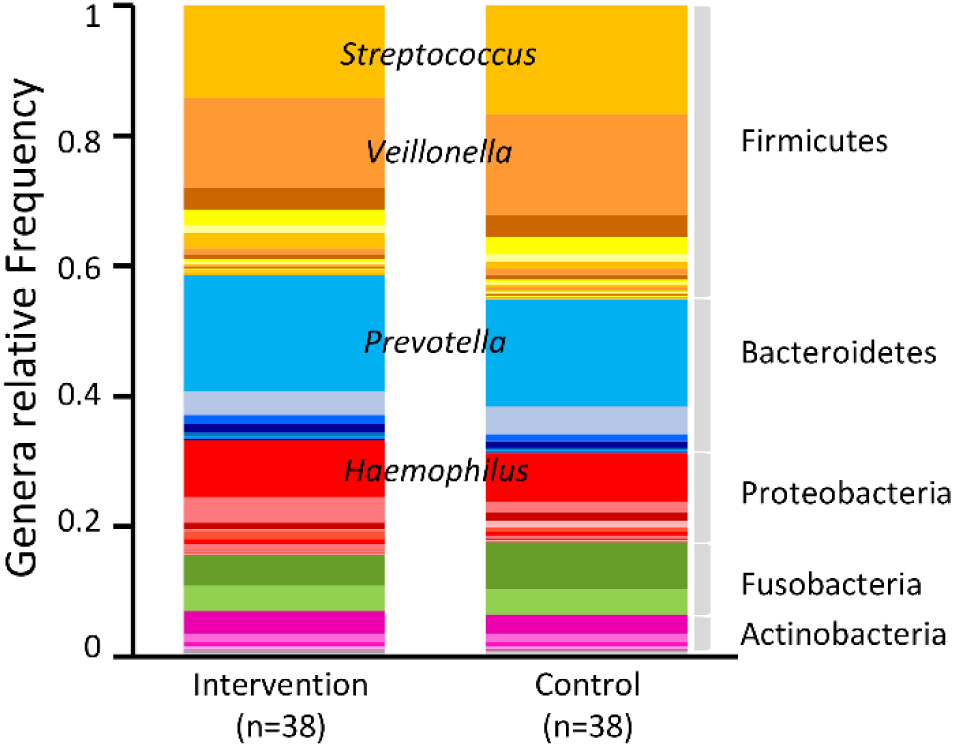
Mean frequency of phyla and genera of bacteria present in intervention and control groups at baseline (M0).

**Supplementary Figure 2.**
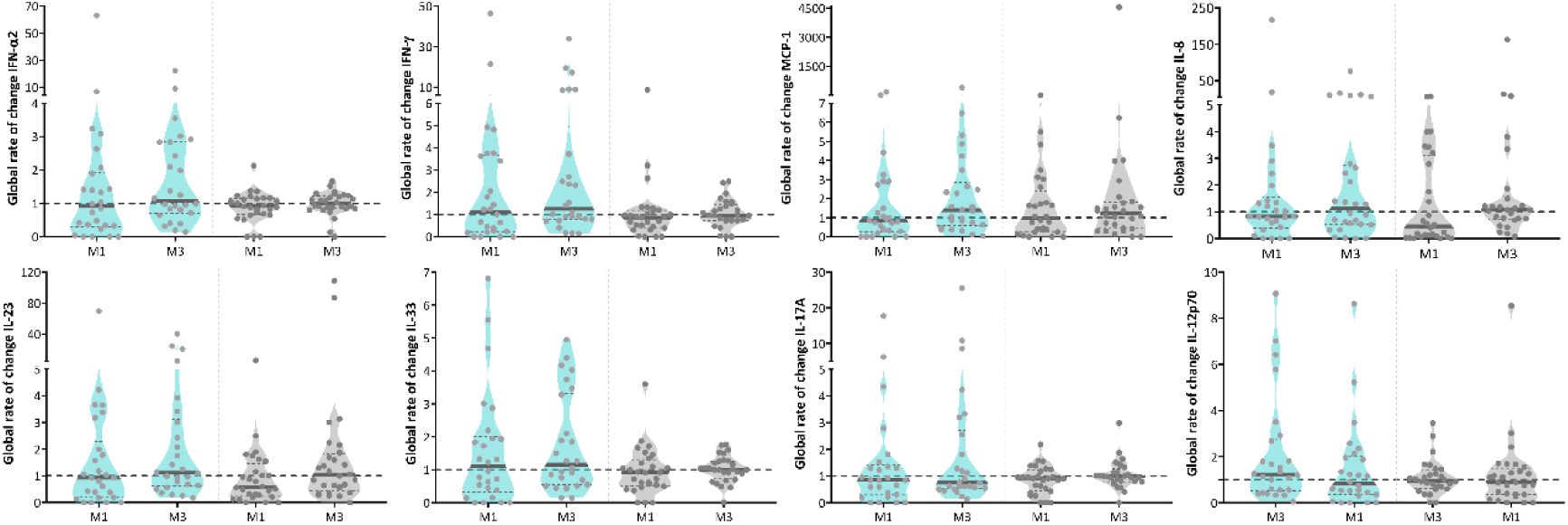
Violin plots representing the global rate of change of IFN-α2, IFN-ϒ, MCP-1, IL-8, IL-23, IL-33, IL-17A, IL-18 and IL-12p70 in saliva from patients submitted to the 12- week pulmonary rehabilitation programme (blue) and controls (grey). Global rate of change represents the ratio between cytokine values measured at baseline and M1 or M3 (*i.e.* M1/M0 and M3/M0). Differences between M1 and the baseline and between M3 and the baseline were assessed by Wilcoxon signed-rank, in each group (intervention (blue) and control (grey)). Differences between groups at M1 and M3 were assessed by Mann-Whitney U-test. According to these criteria no significant shifts were observed for the cytokines represented in the figure.

**Supplementary Figure 3.**
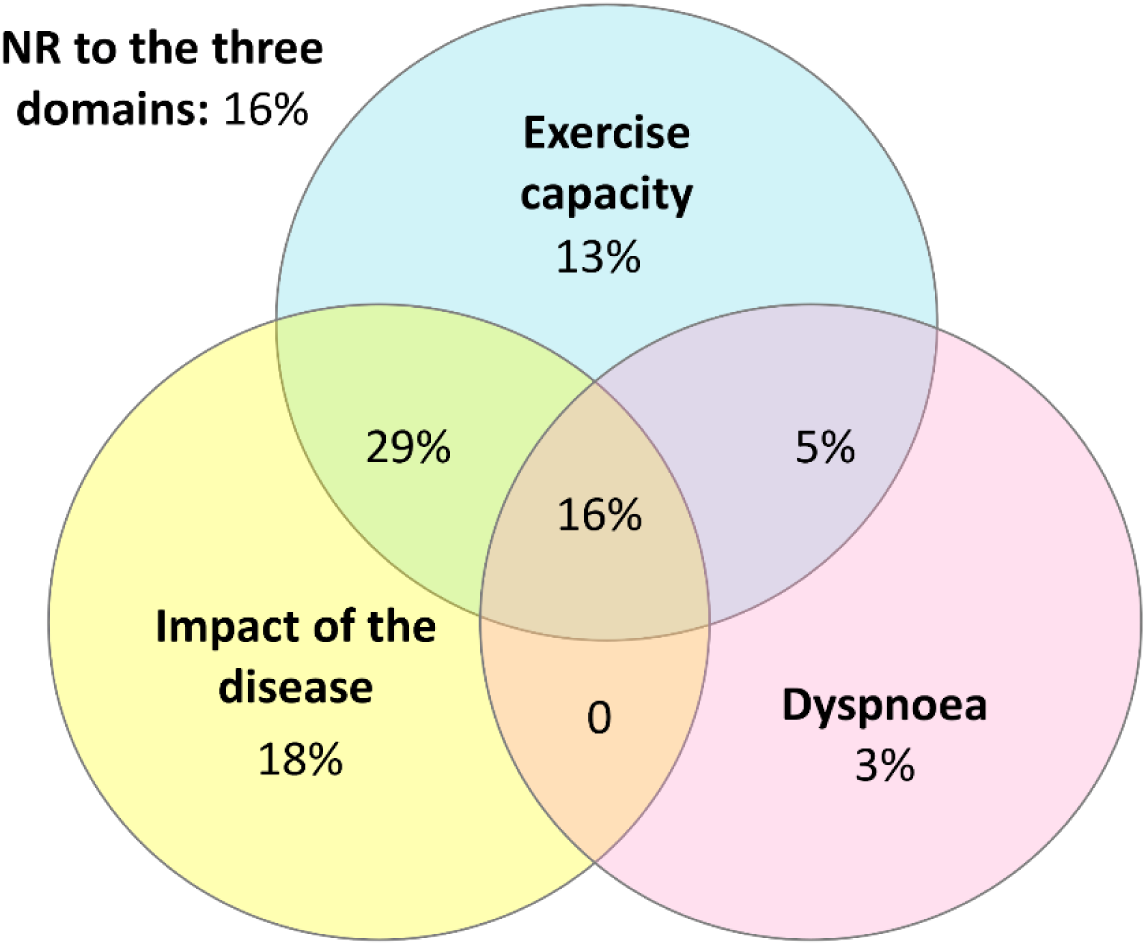
Venn diagram showing the percentage of overlap between patients’ (n=38) responsiveness to each domain: dyspnoea, exercise capacity and impact of the disease.

**Supplementary Figure 4.**
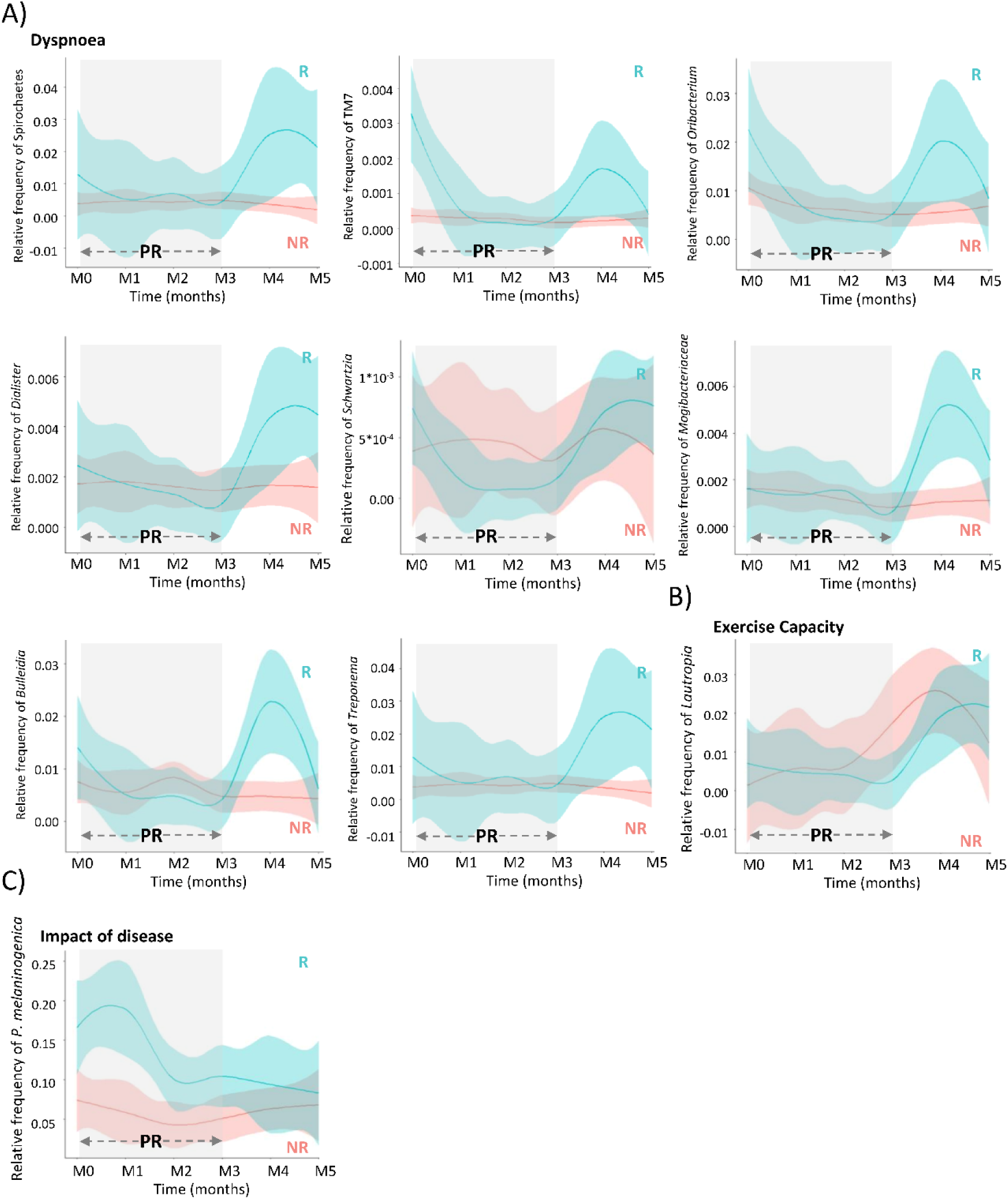
Relative frequencies over time of taxa presenting significantly different dynamics between responders and non-responders. Linear mixed-effects models were applied to arcsine square root transformed frequencies of ASVs/OTUs to determine the taxa presenting differential dynamics between R and NR to A) dyspnoea during exercise, B) exercise capacity and C) impact of the disease. Grey rectangles include all the time-points where patients were under pulmonary rehabilitation. Blue and red loess lines were fitted over the data points representing the relative frequency of the correspondent taxon in responders and non-responders (M0=immediately prior to PR; M1, M2, M3= 1,2,3 months after initiating PR; M4, M5=1 and 2 months after terminating PR). The blue and red areas represent the 0.95 confidence intervals.

**Supplementary Figure 5.**
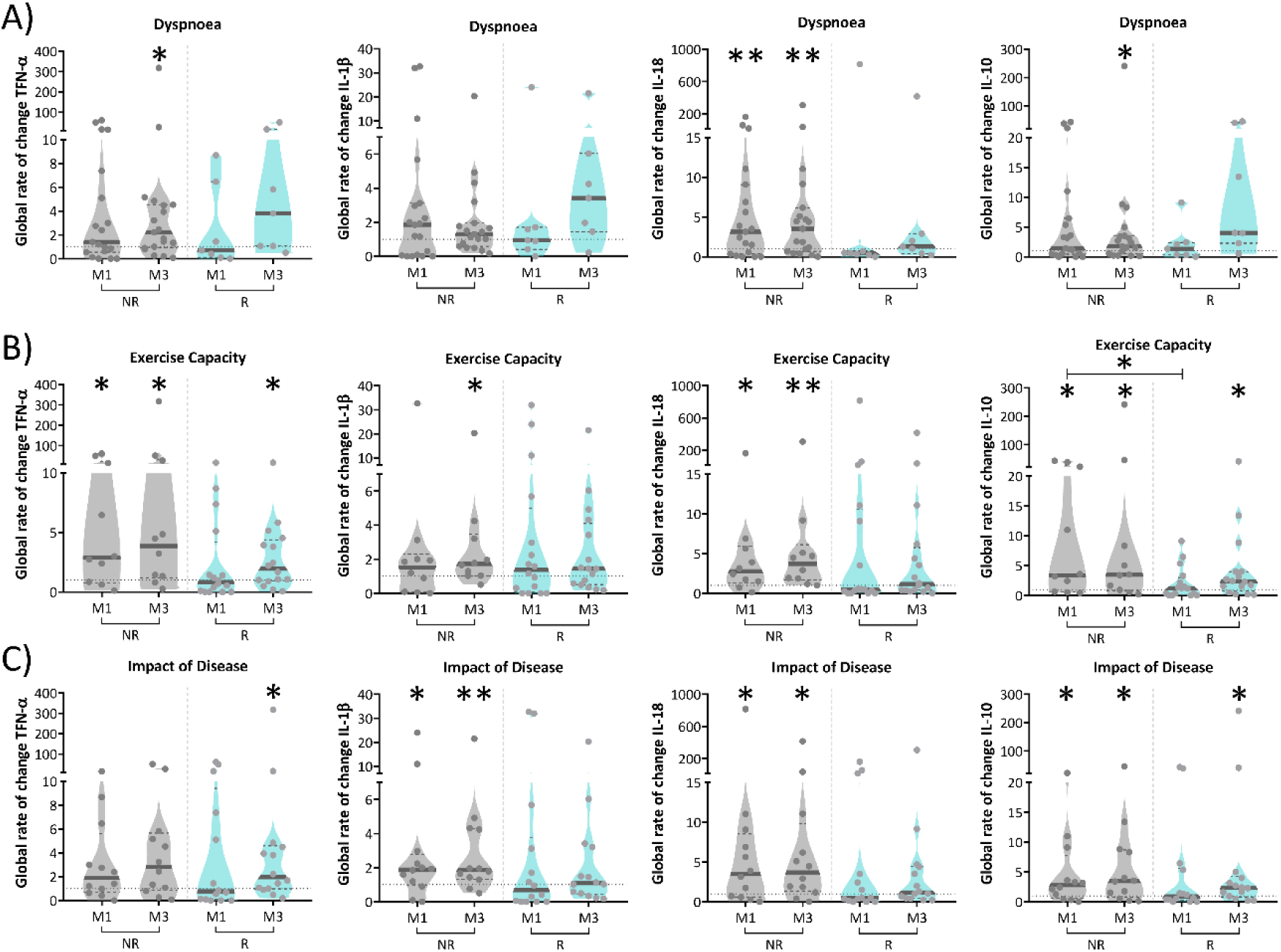
Responsiveness to pulmonary rehabilitation (PR) was associated with specific alterations in the inflammatory markers. Global rate of change represents the ratio between cytokine values measured at baseline and M1 or M3 (*i.e*. M1/M0 and M3/M0). Differences between M1 and the baseline and between M3 and the baseline were assessed by Wilcoxon signed-rank, in each group (intervention (blue) and control (grey)). Differences between groups (R and NR) at M1 and M3 were assessed by Mann-Whitney U-test. *p<0.05, **p<0.01, **p<0.001. A) dyspnoea during exercise, B) exercise capacity and C) impact of the disease.

**Supplementary Figure 6.**
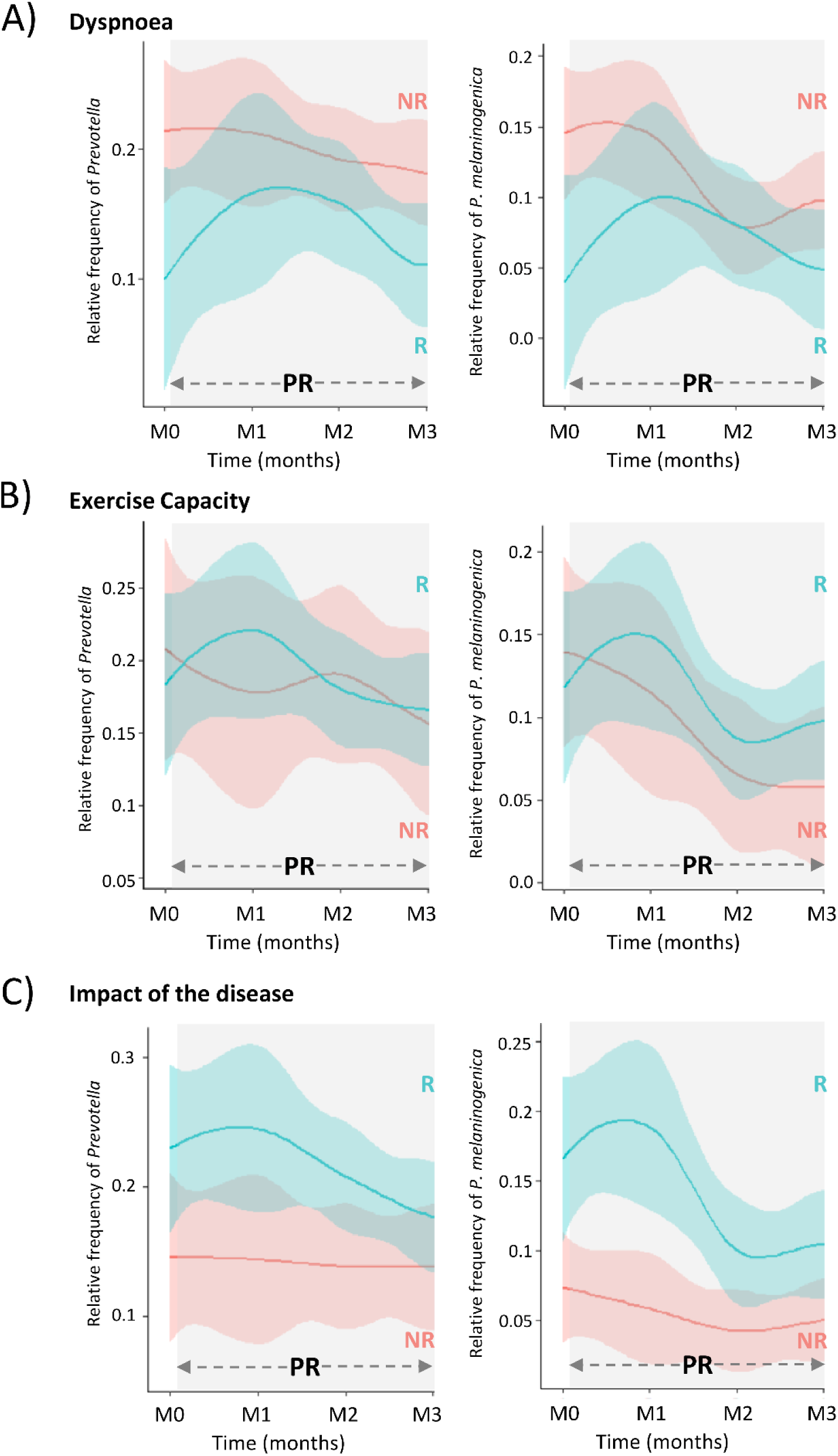
Frequency of *Prevotella* and *P. melaninogenica* over time in responders and non-responders to A) dyspnoea, B) exercise capacity and C) impact of the disease. The grey rectangles include all the time-points where patients were under pulmonary rehabilitation. Blue and red loess lines were fitted over the data points representing the relative frequency of the correspondent taxon in responders and non-responders (M0=immediately prior to PR; M1, M2, M3= 1,2,3 months after initiating PR). The blue and red areas represent the 0.95 confidence intervals.

### Code for data analyses

#### Qiime2 v20.8

The scripts provided were constructed based on the complete dataset, appropriate selection of samples was performed for analyses considering specific time-points and groups of patients with the command: “*Filter samples”*

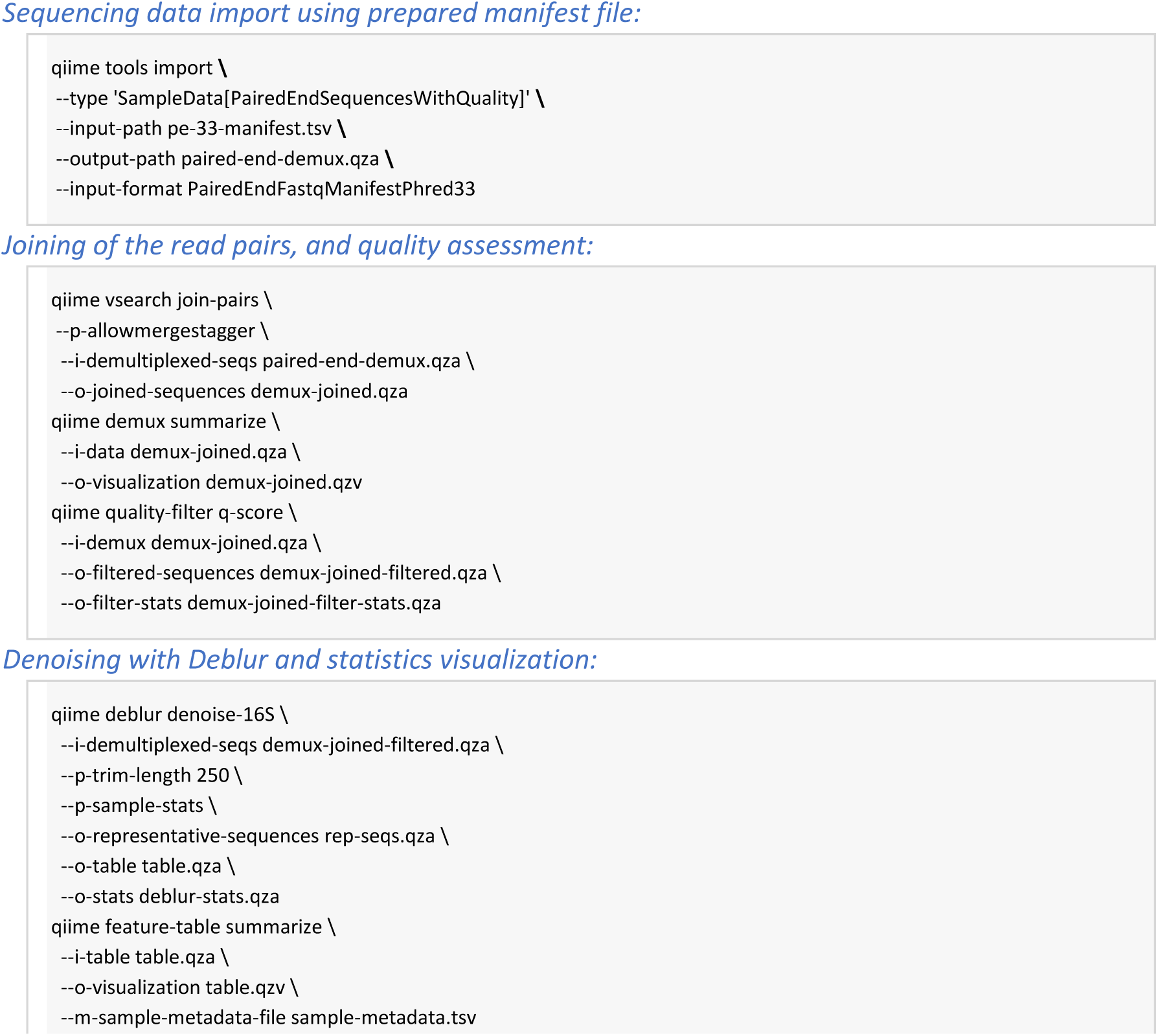
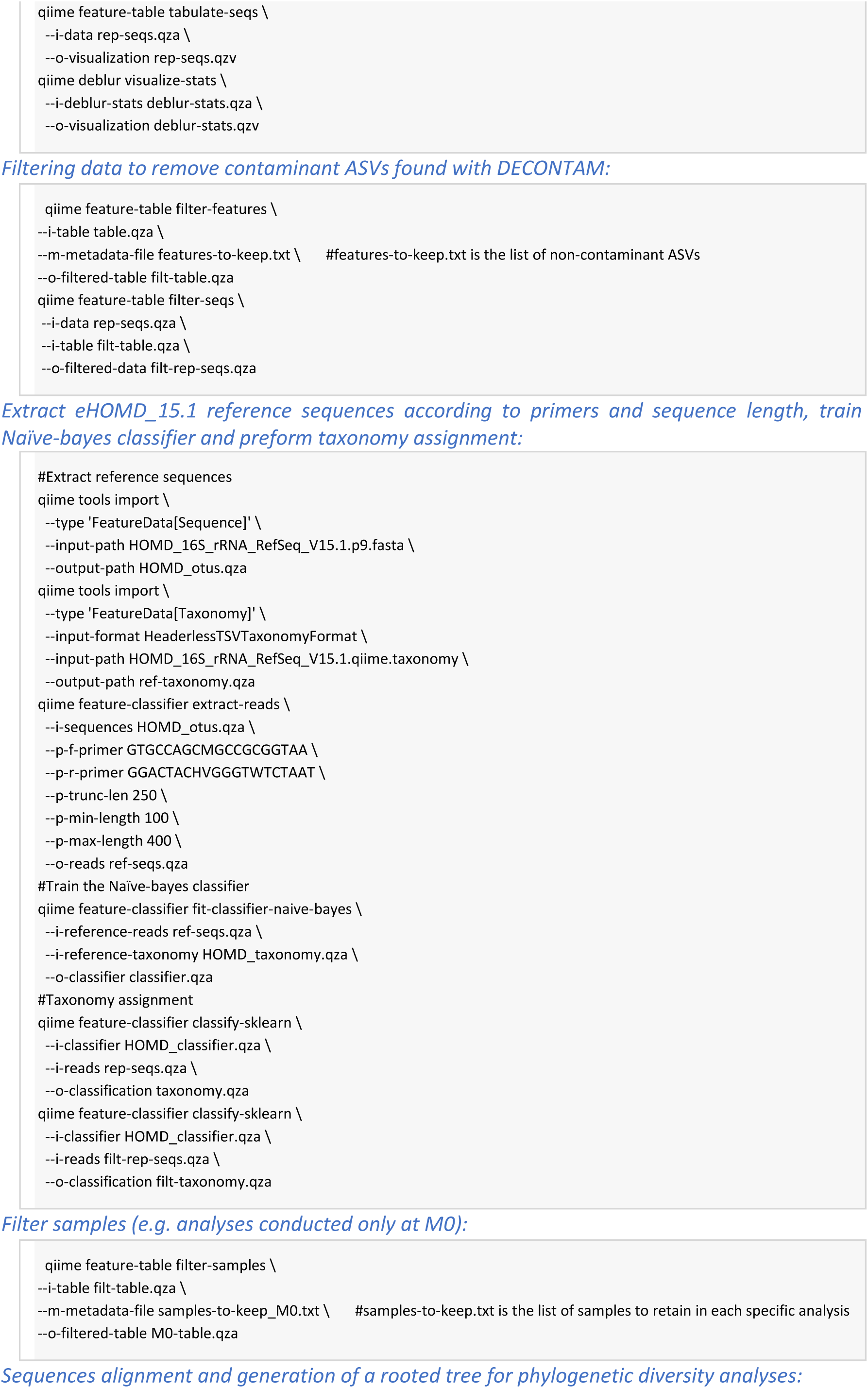
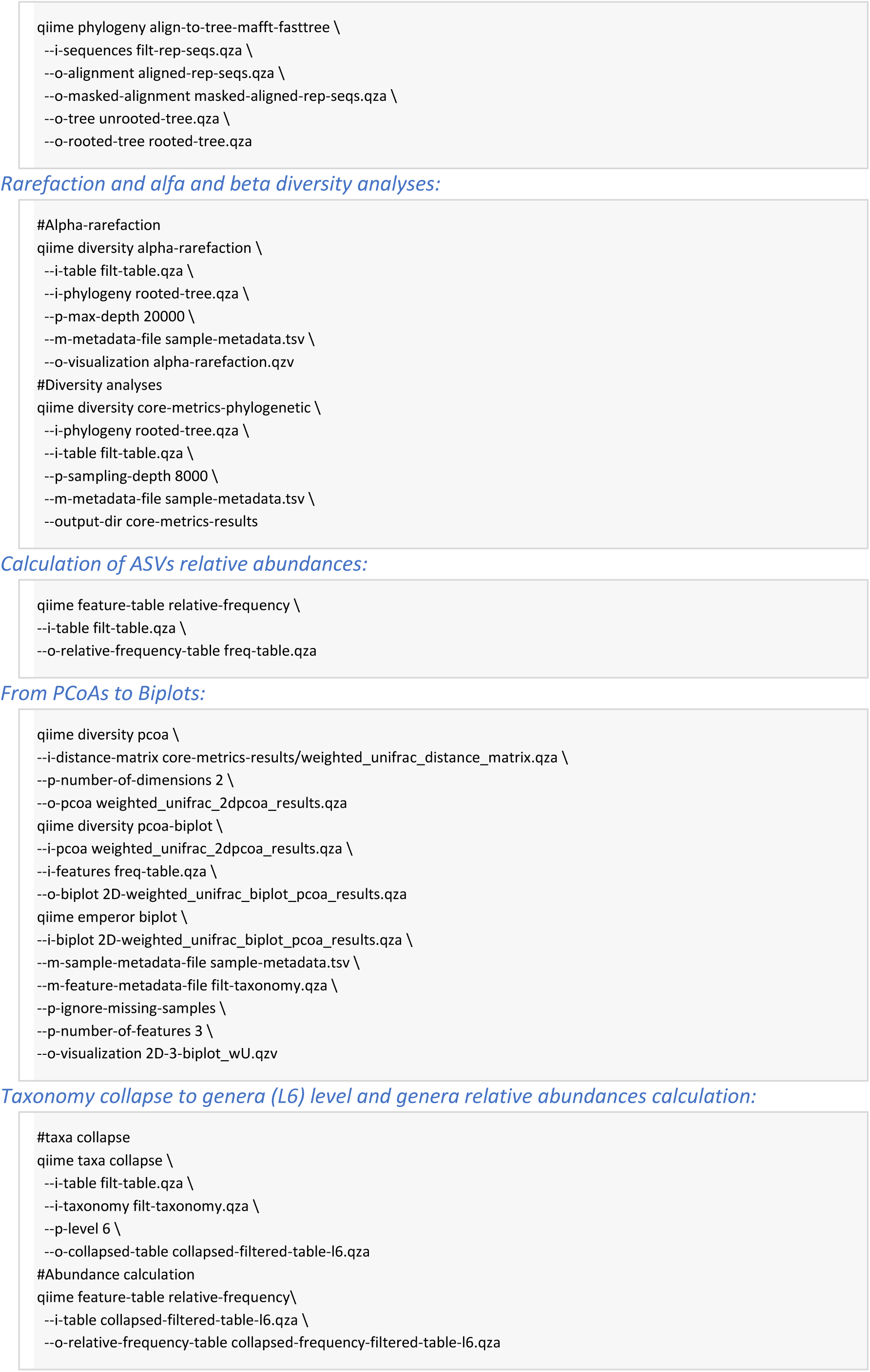
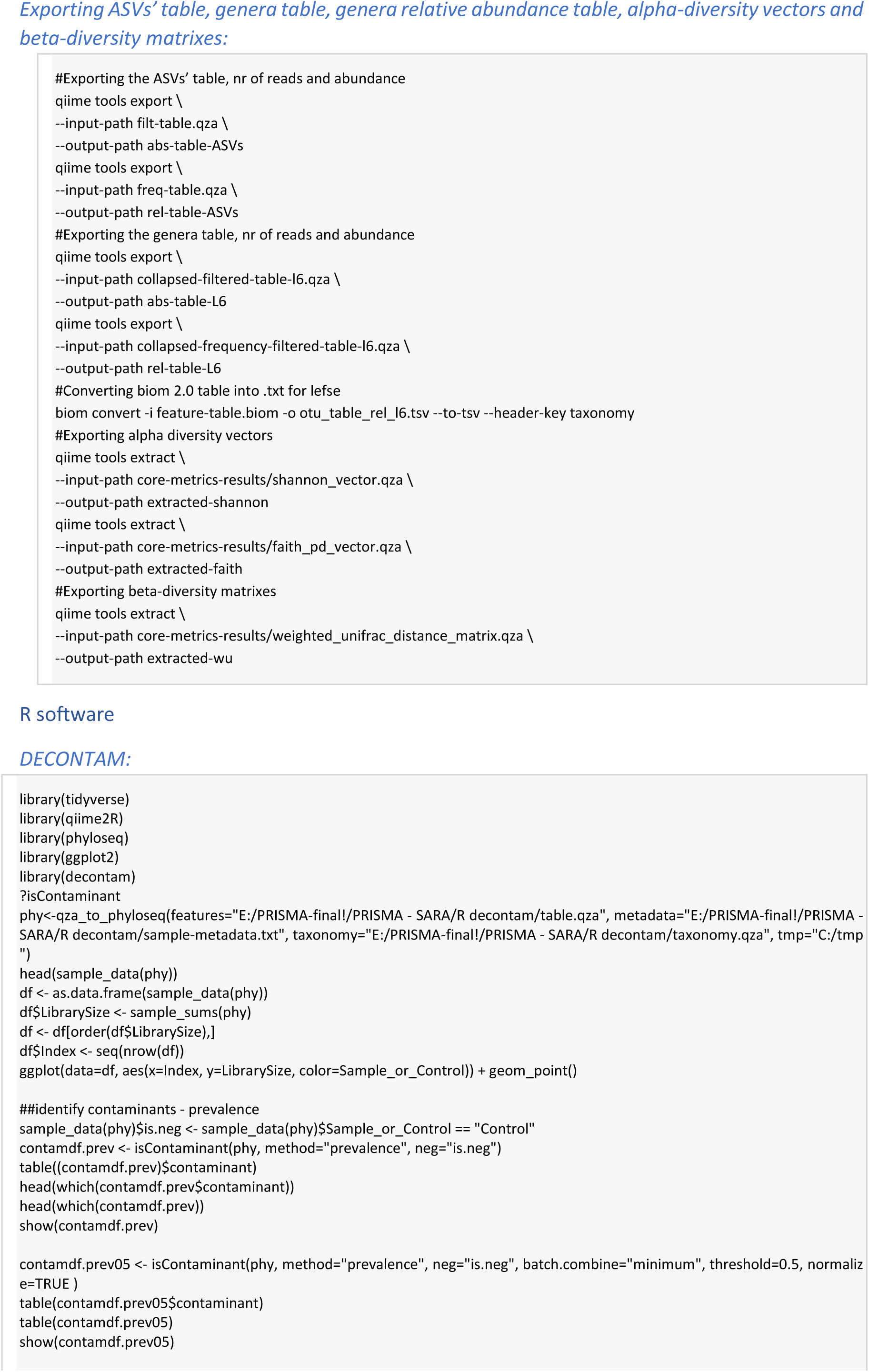
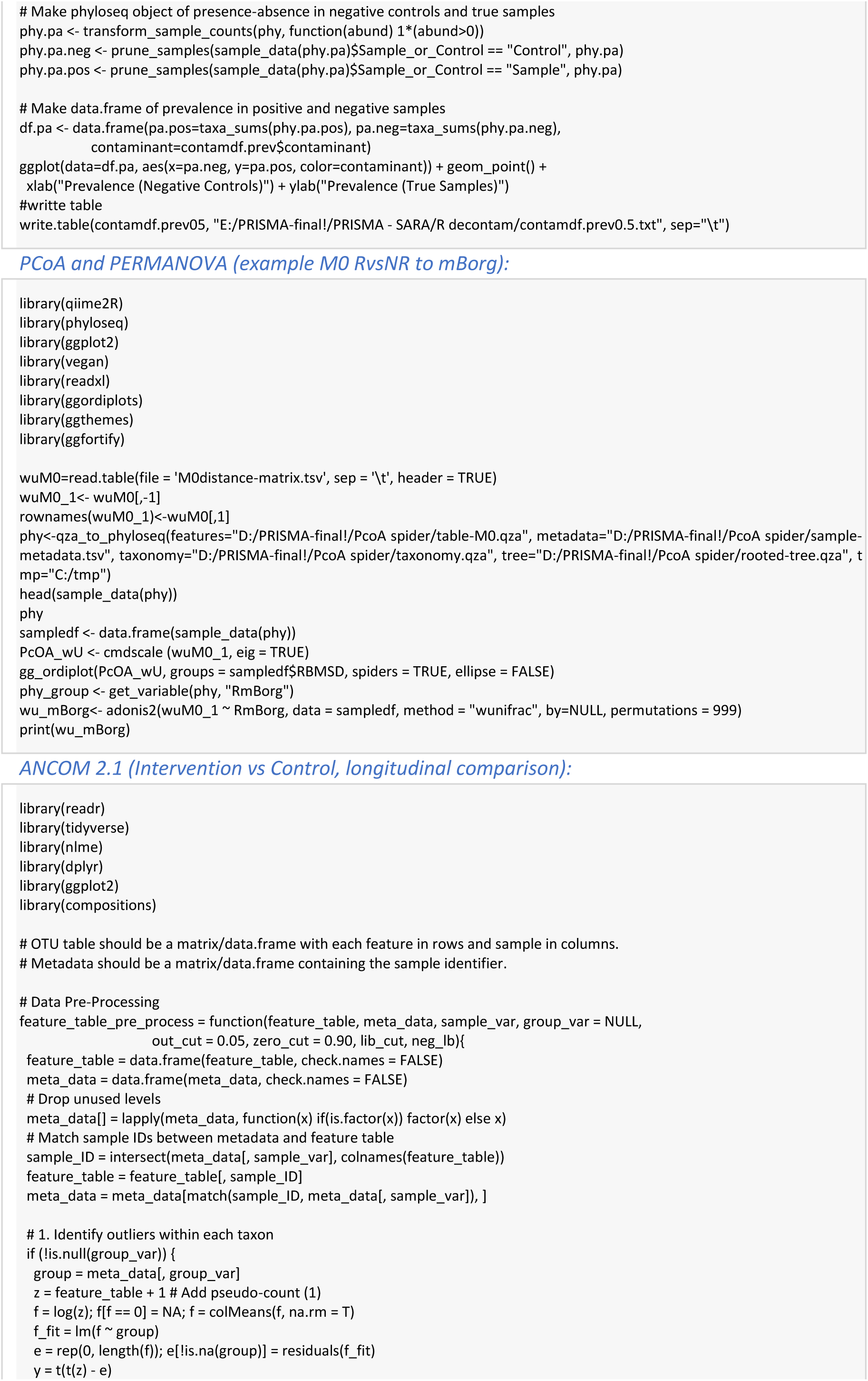
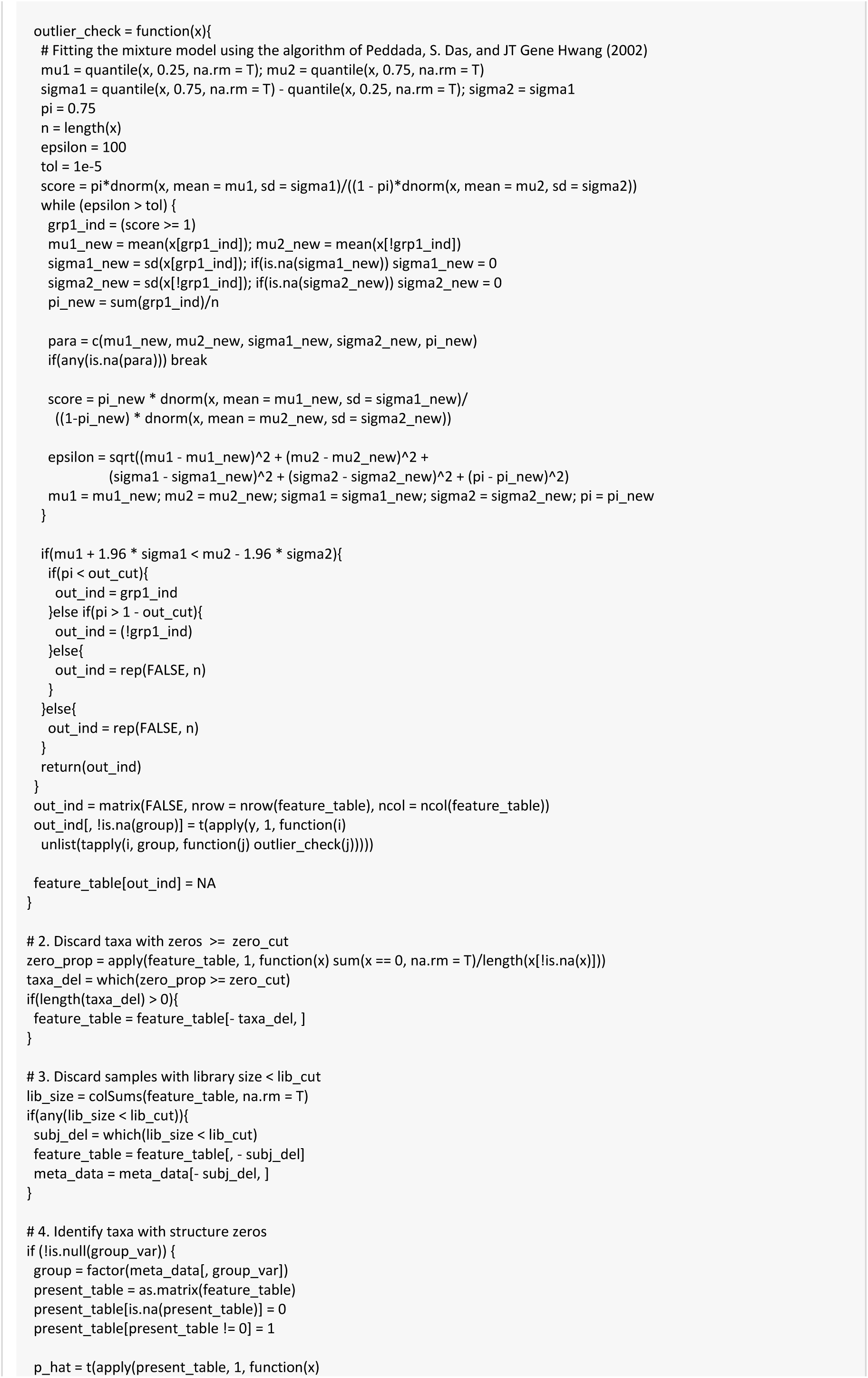
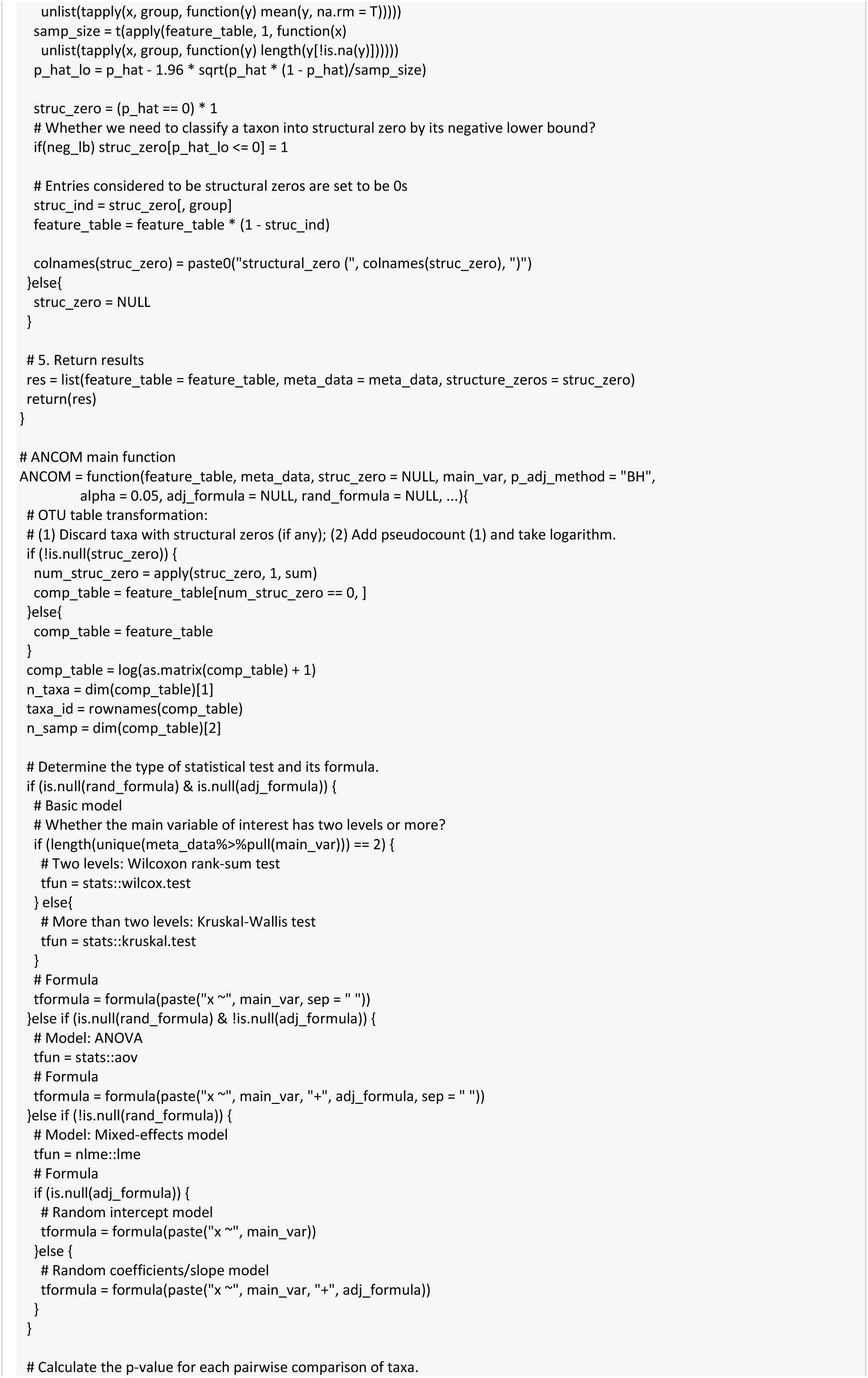
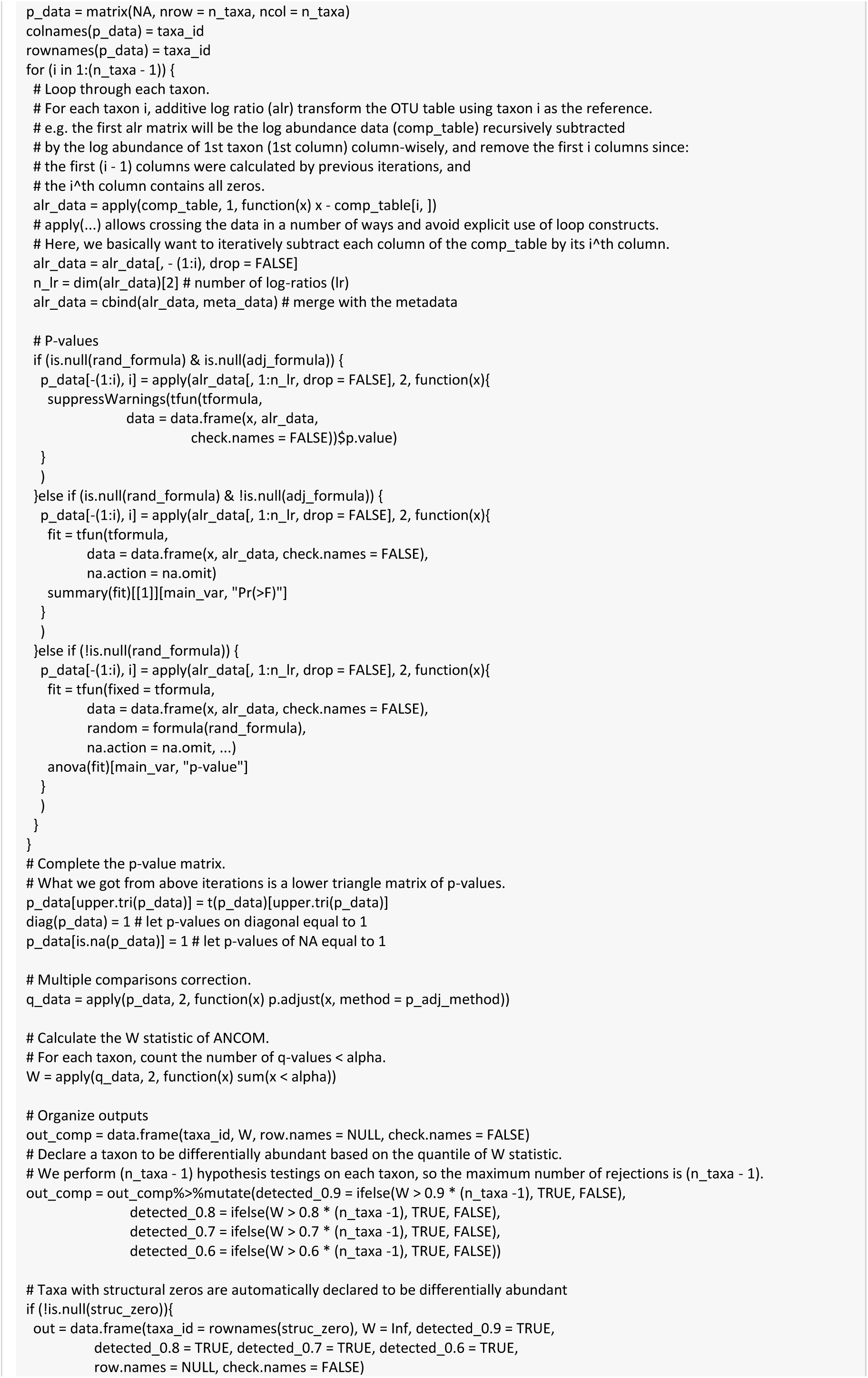
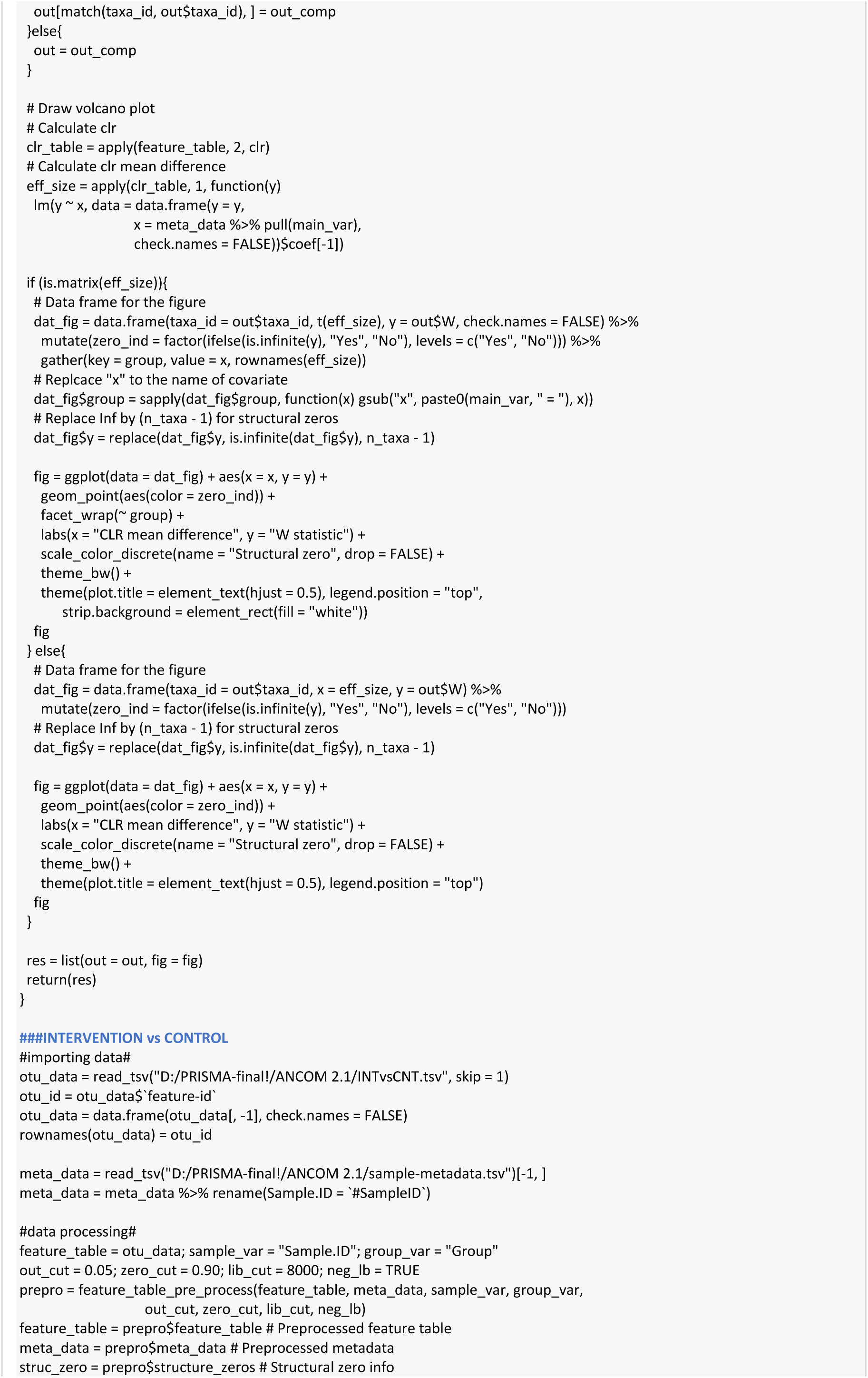
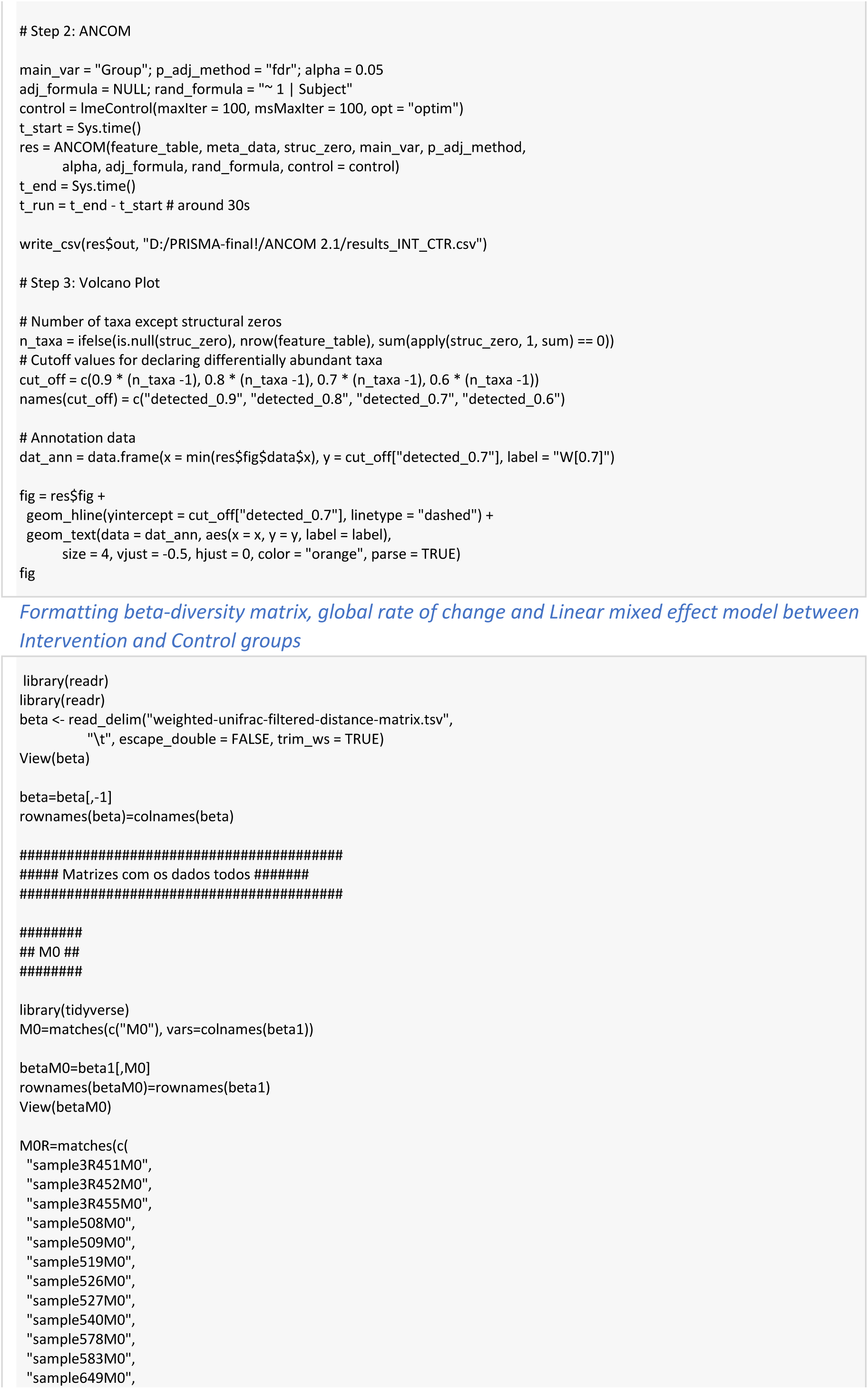
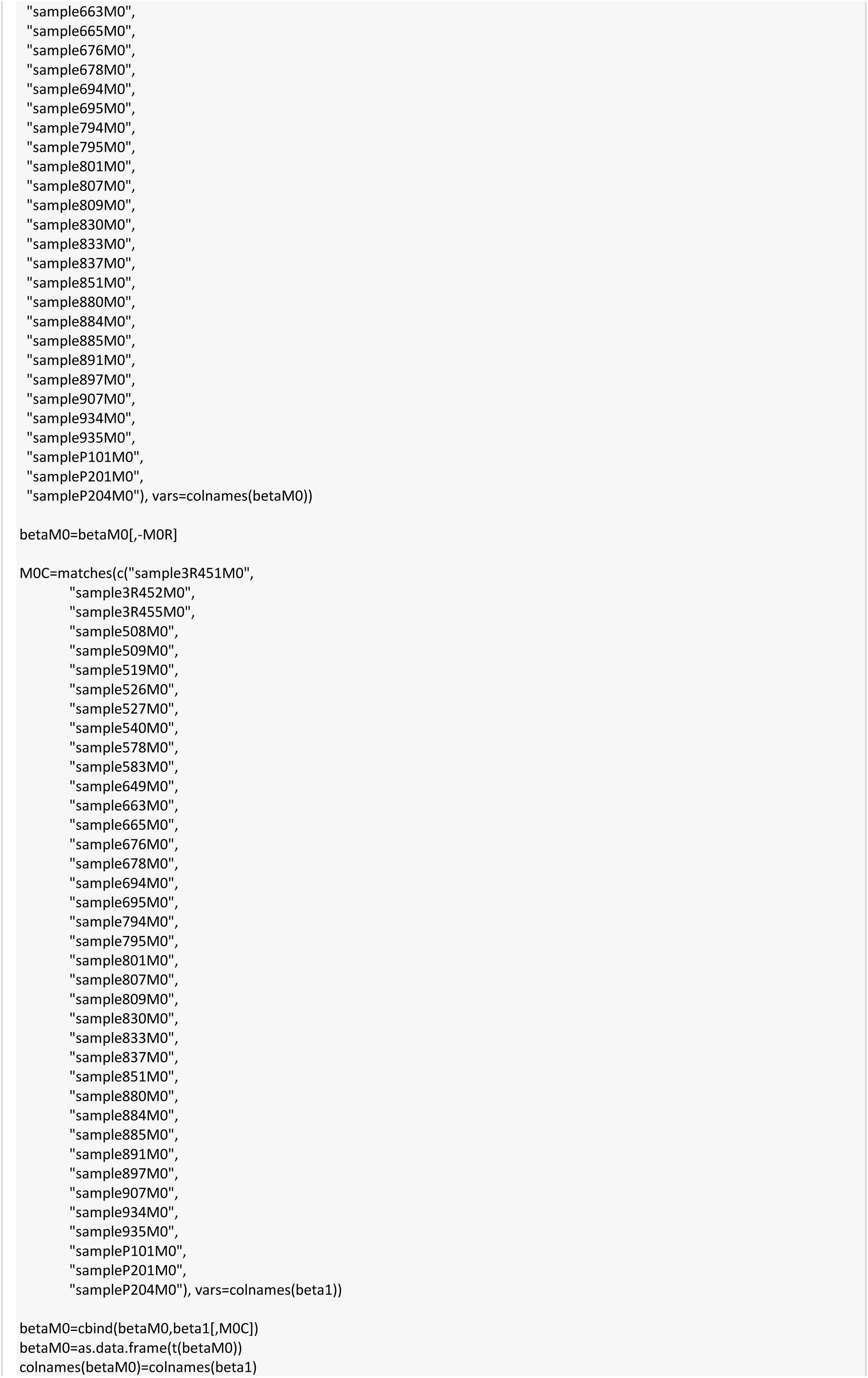
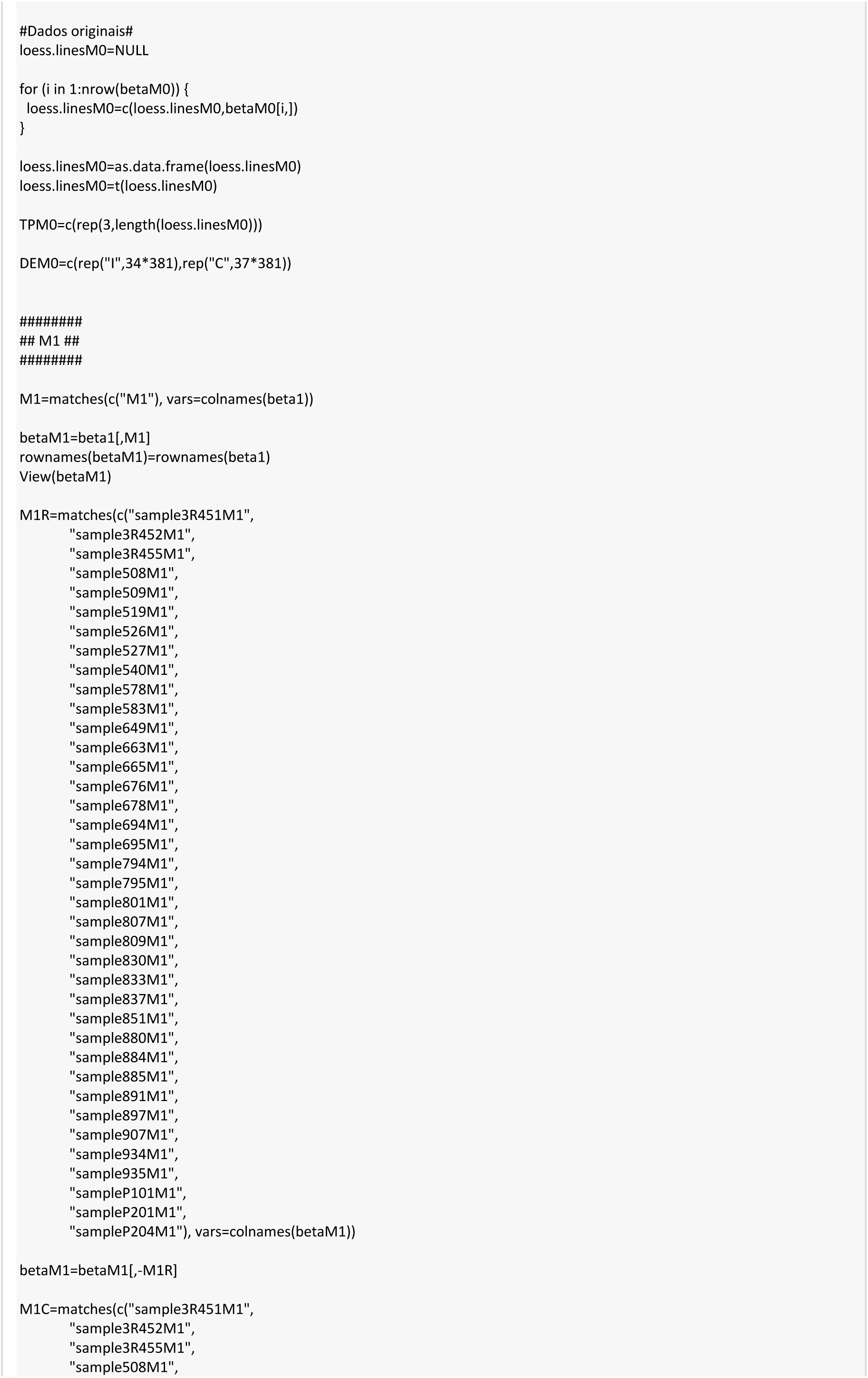
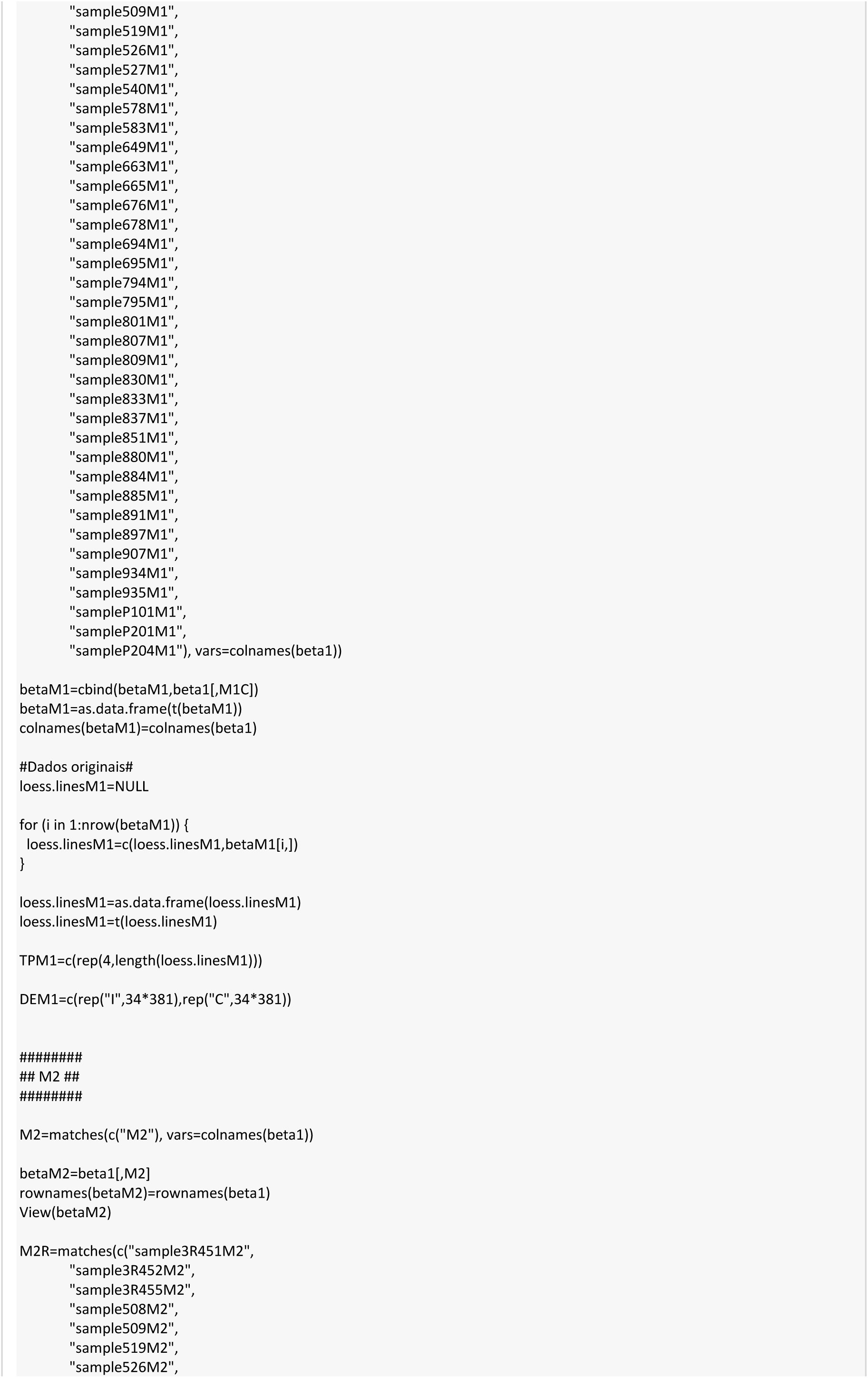
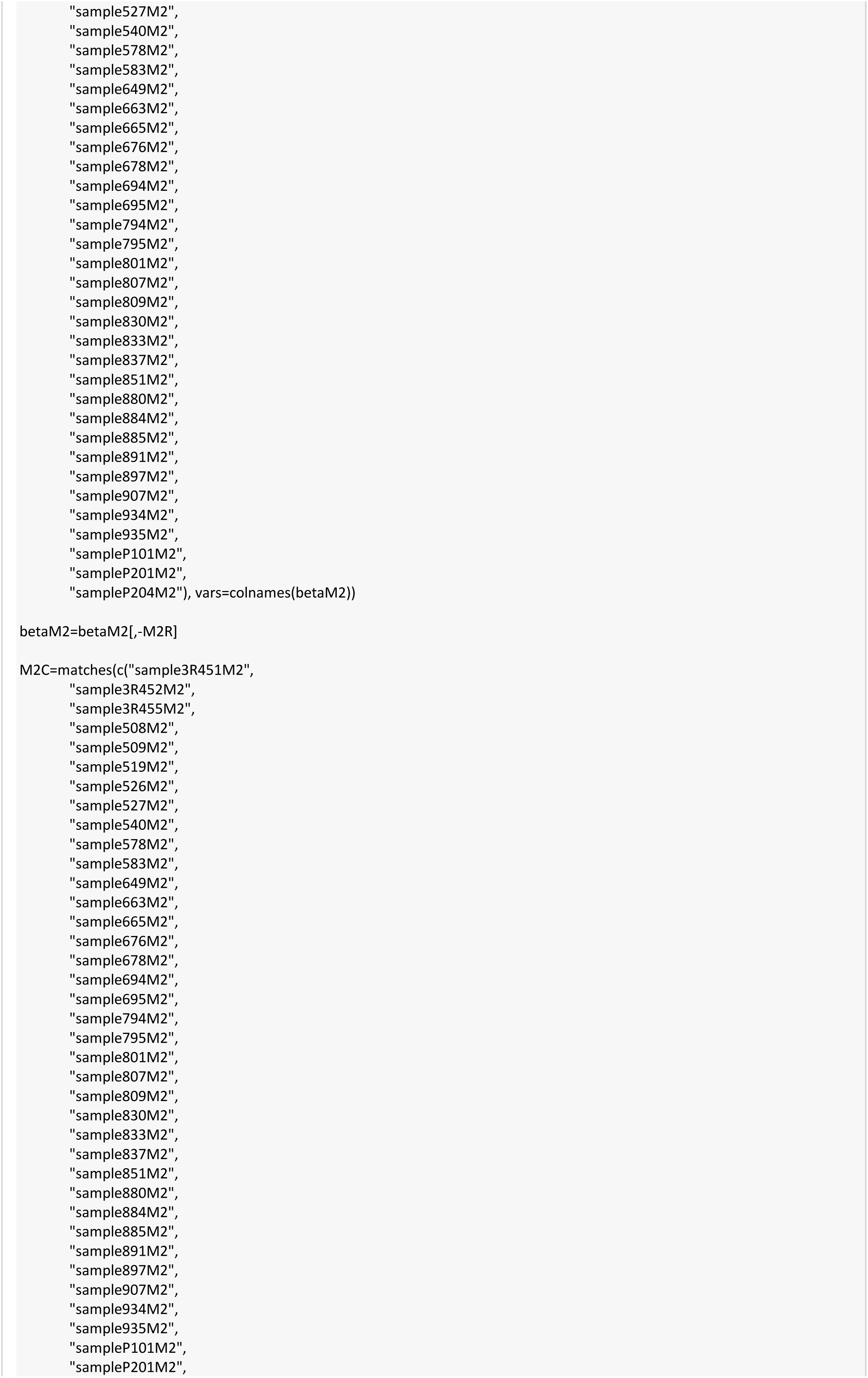
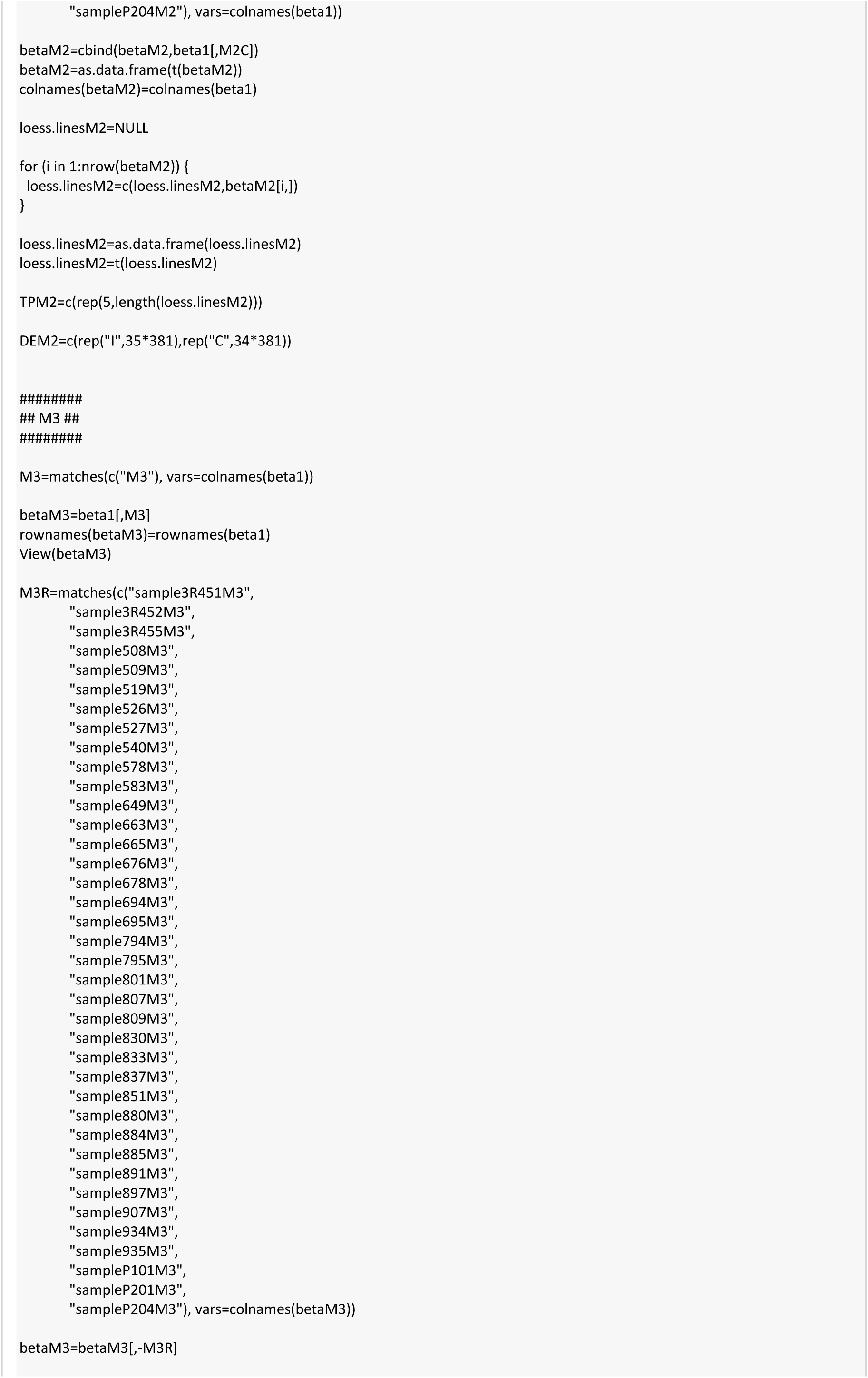
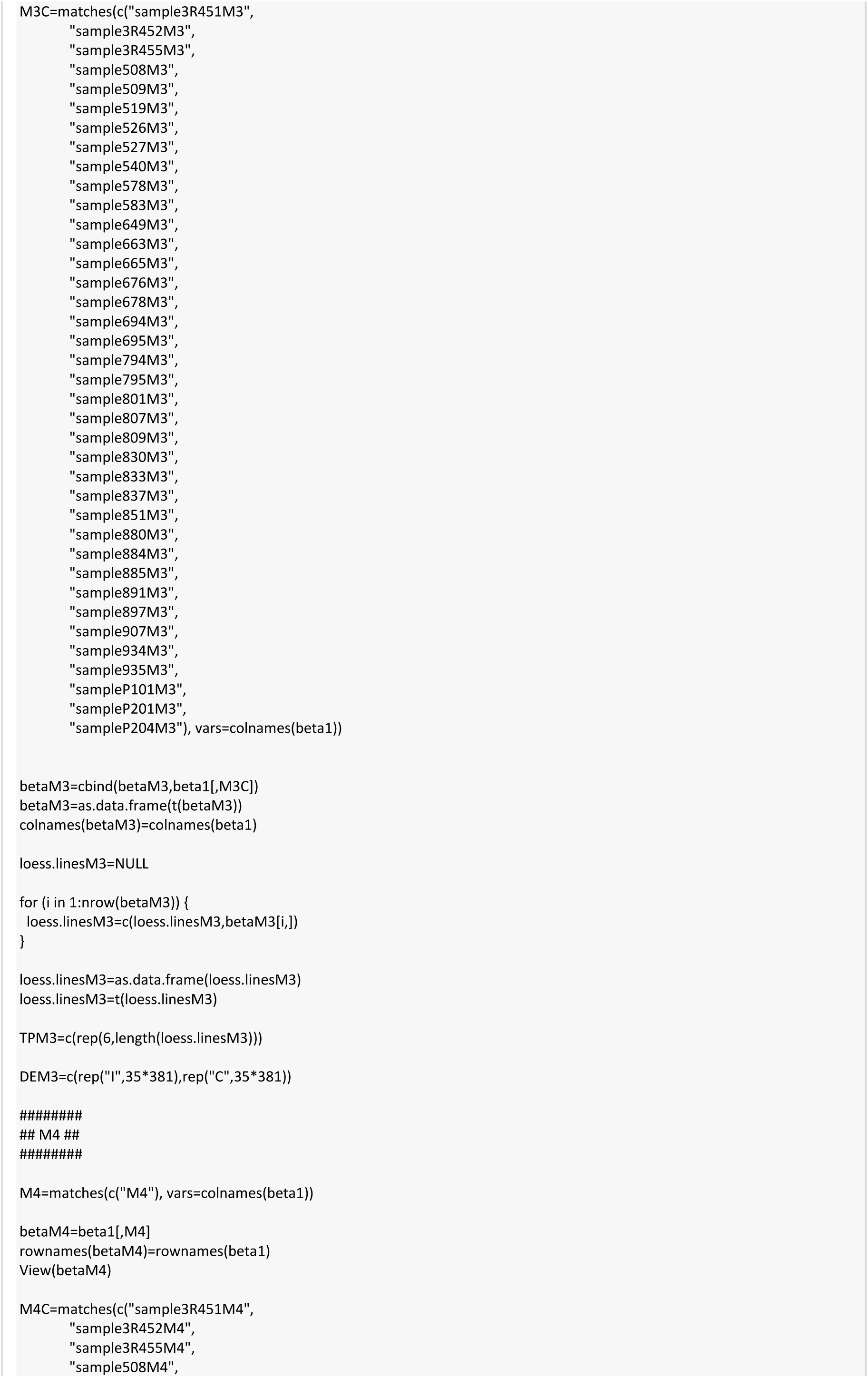
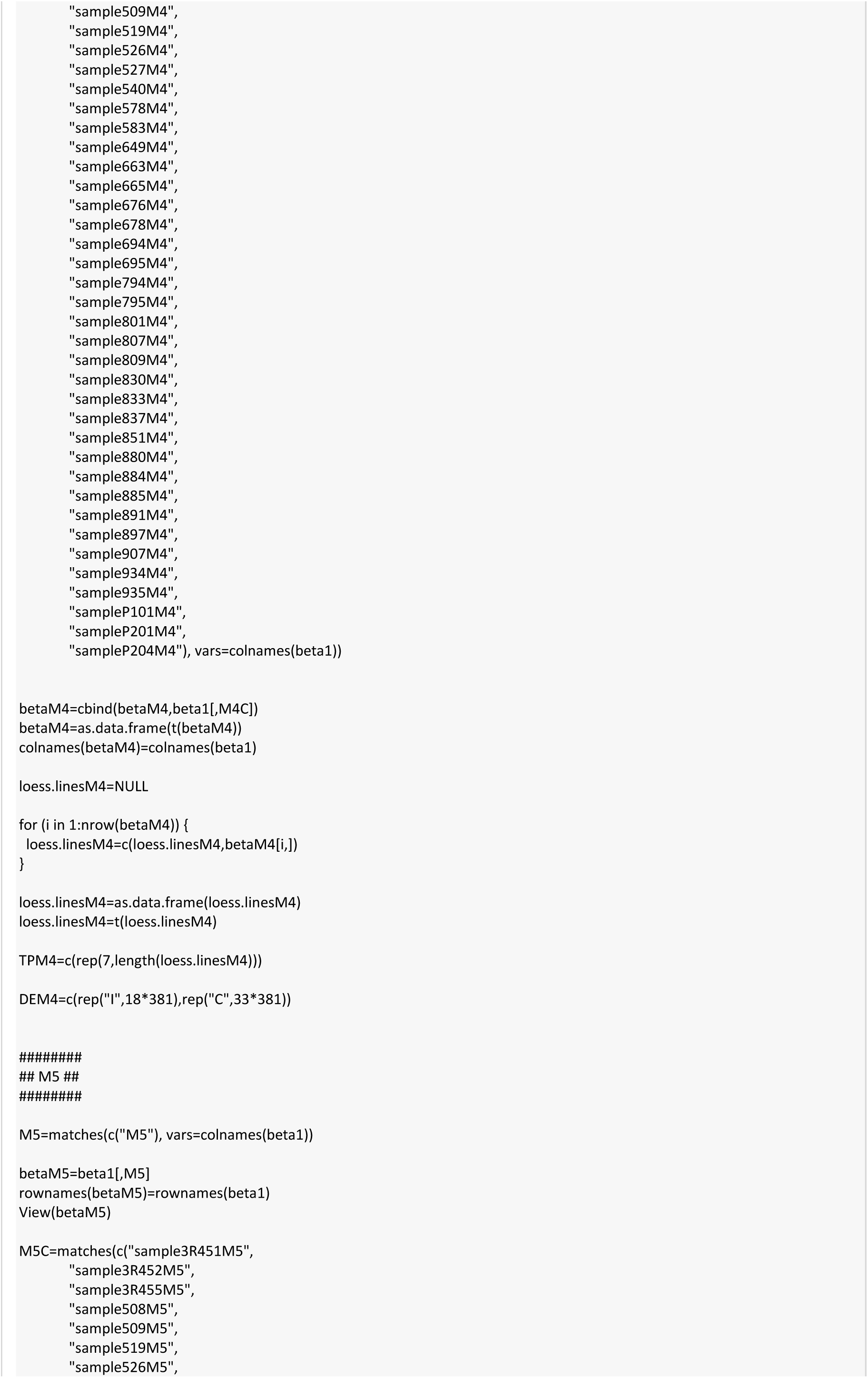
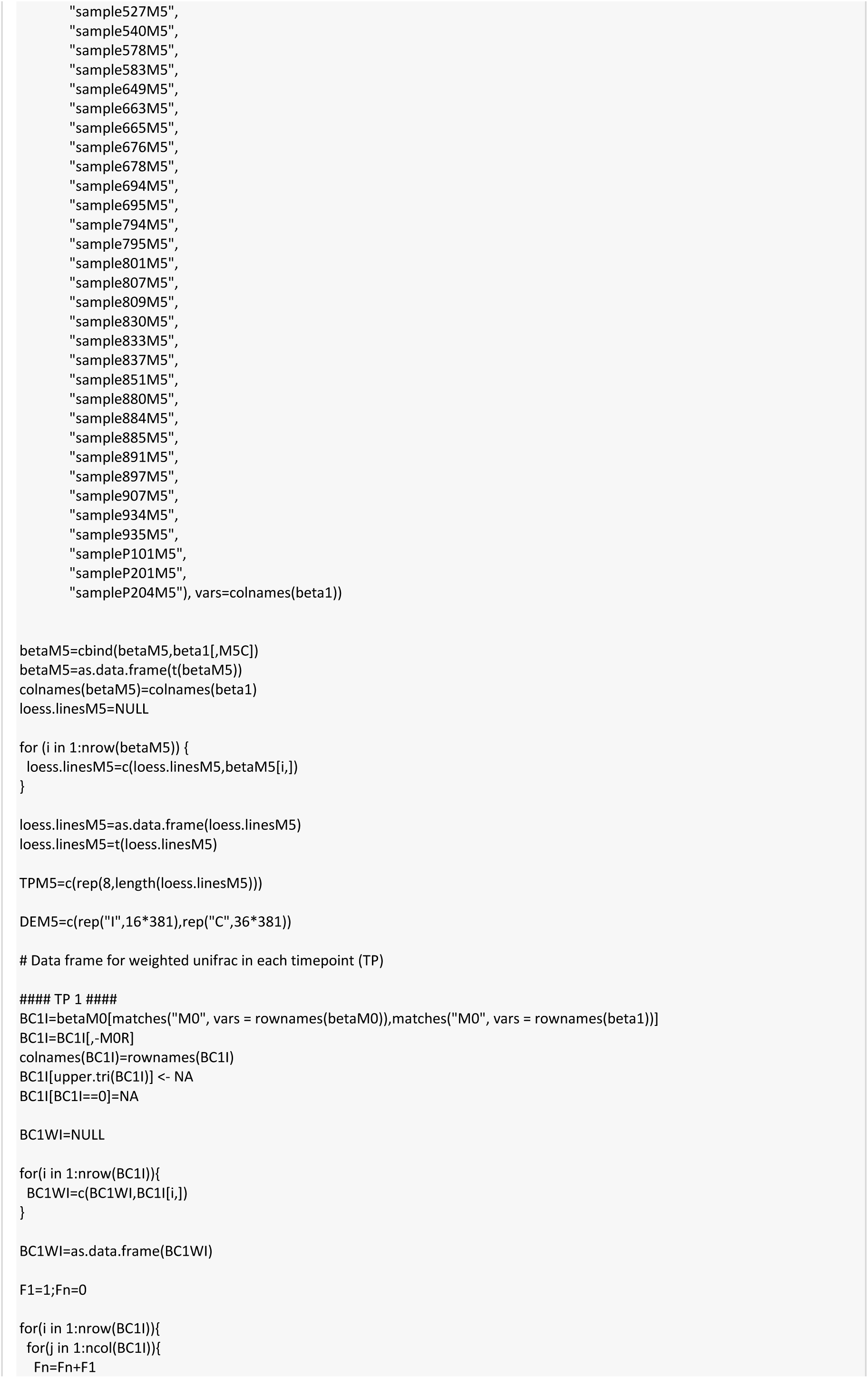
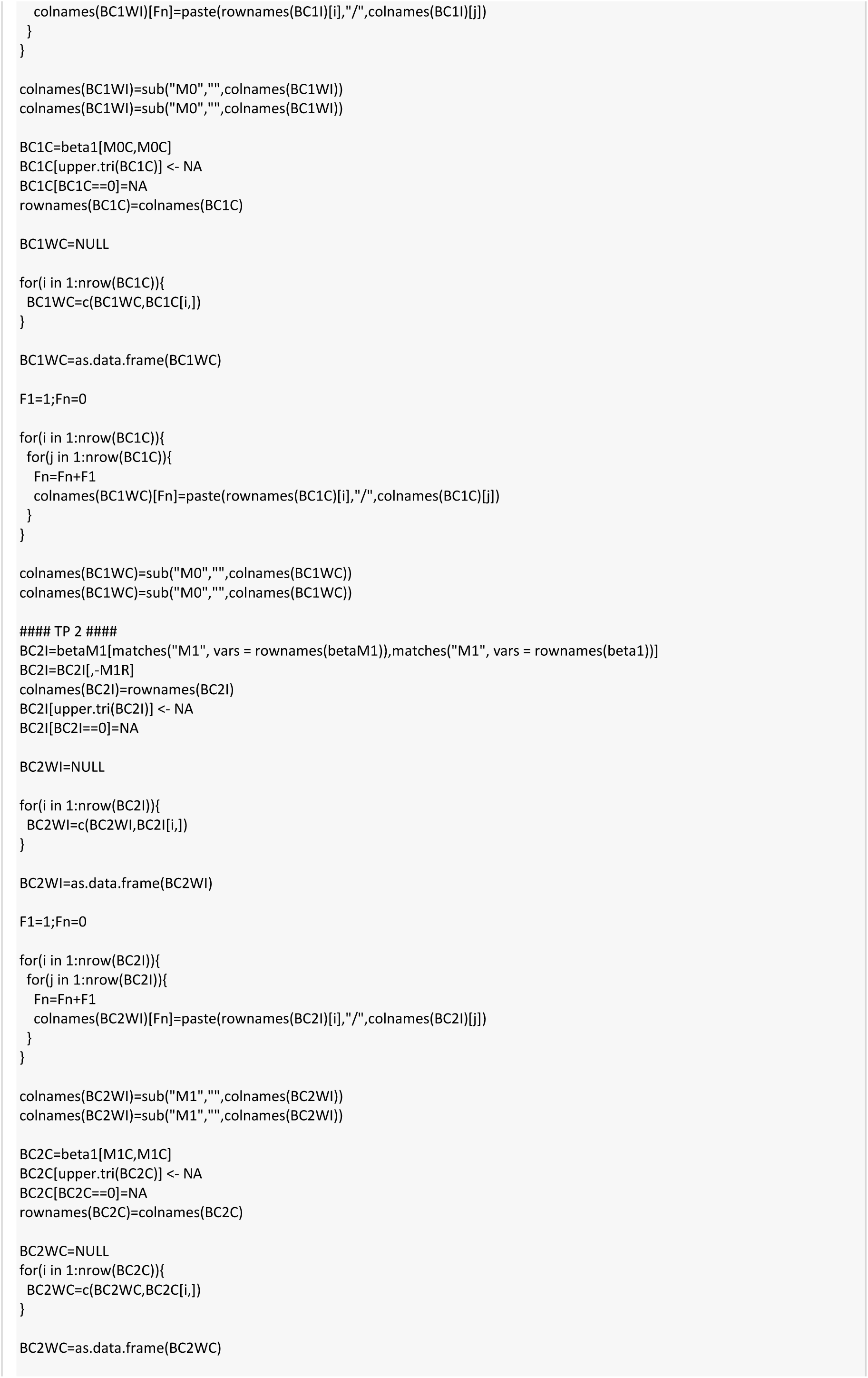
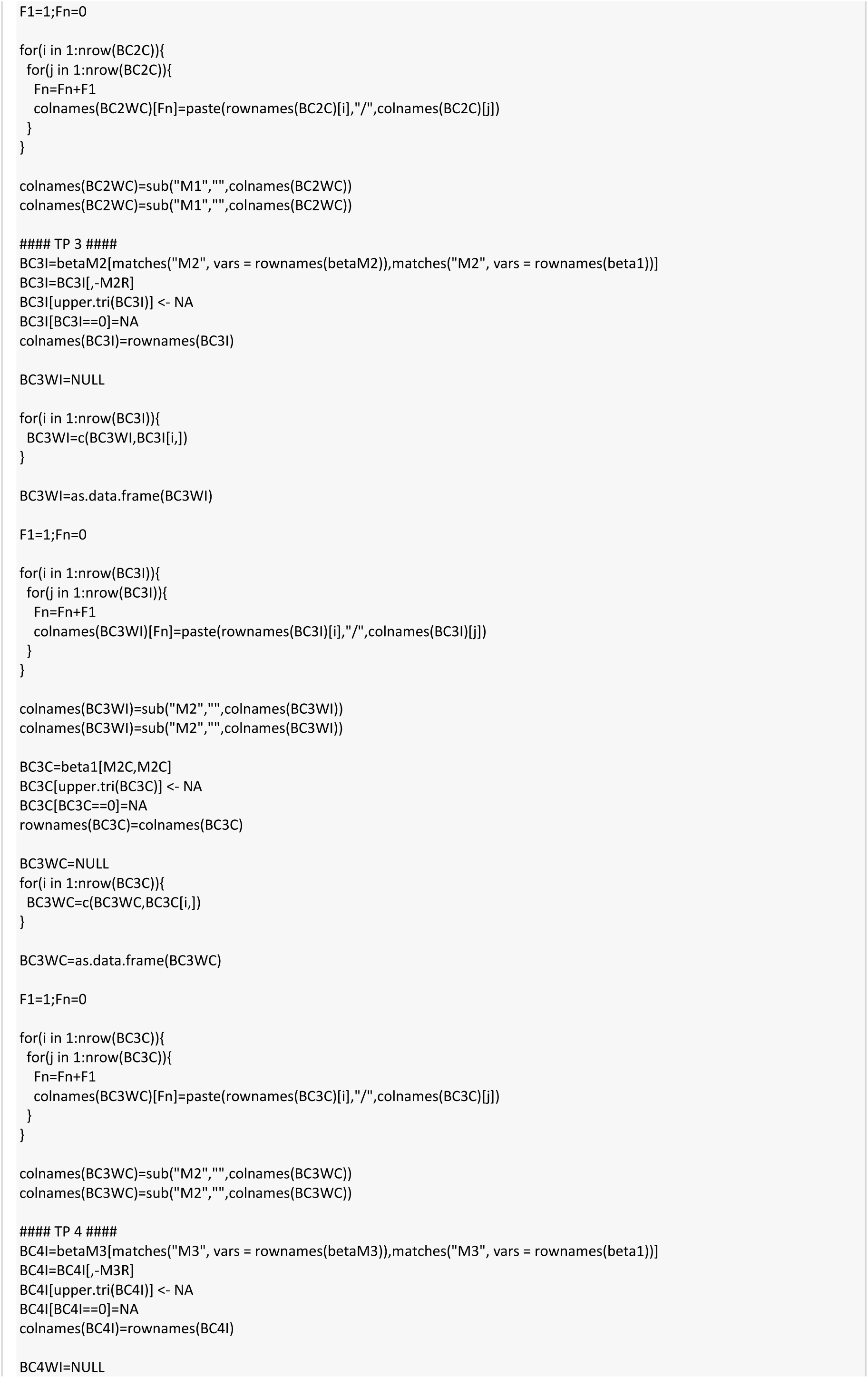
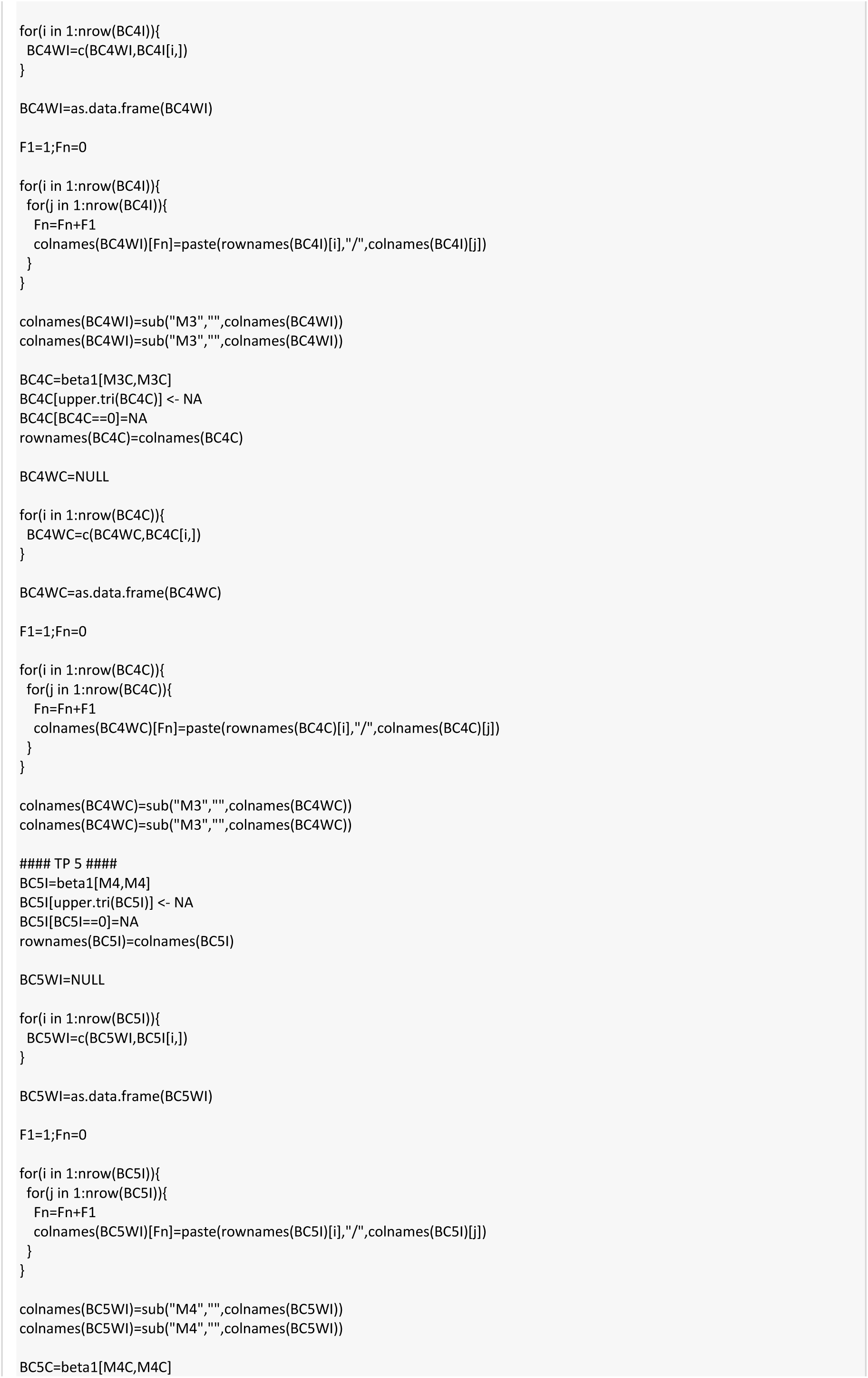
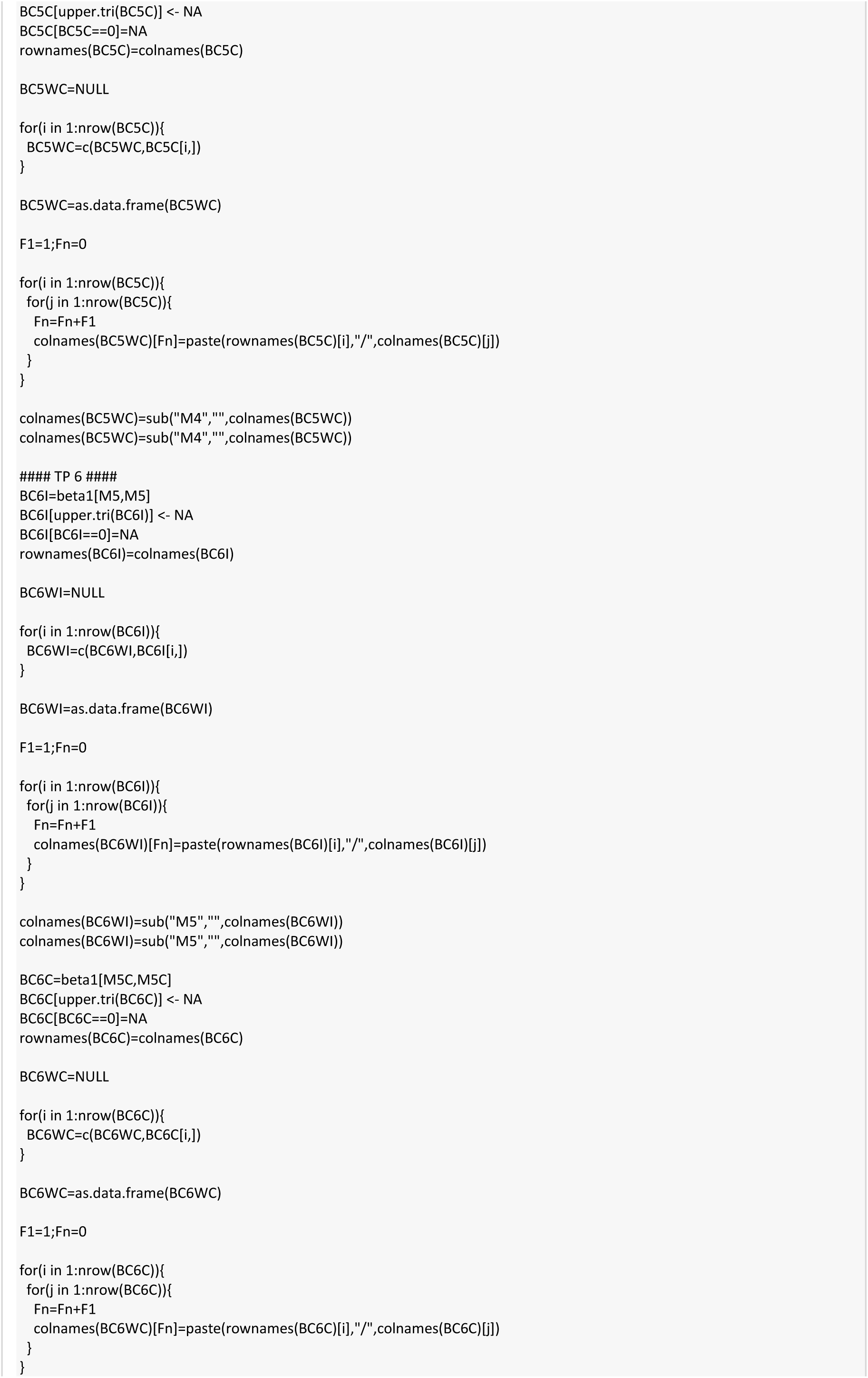
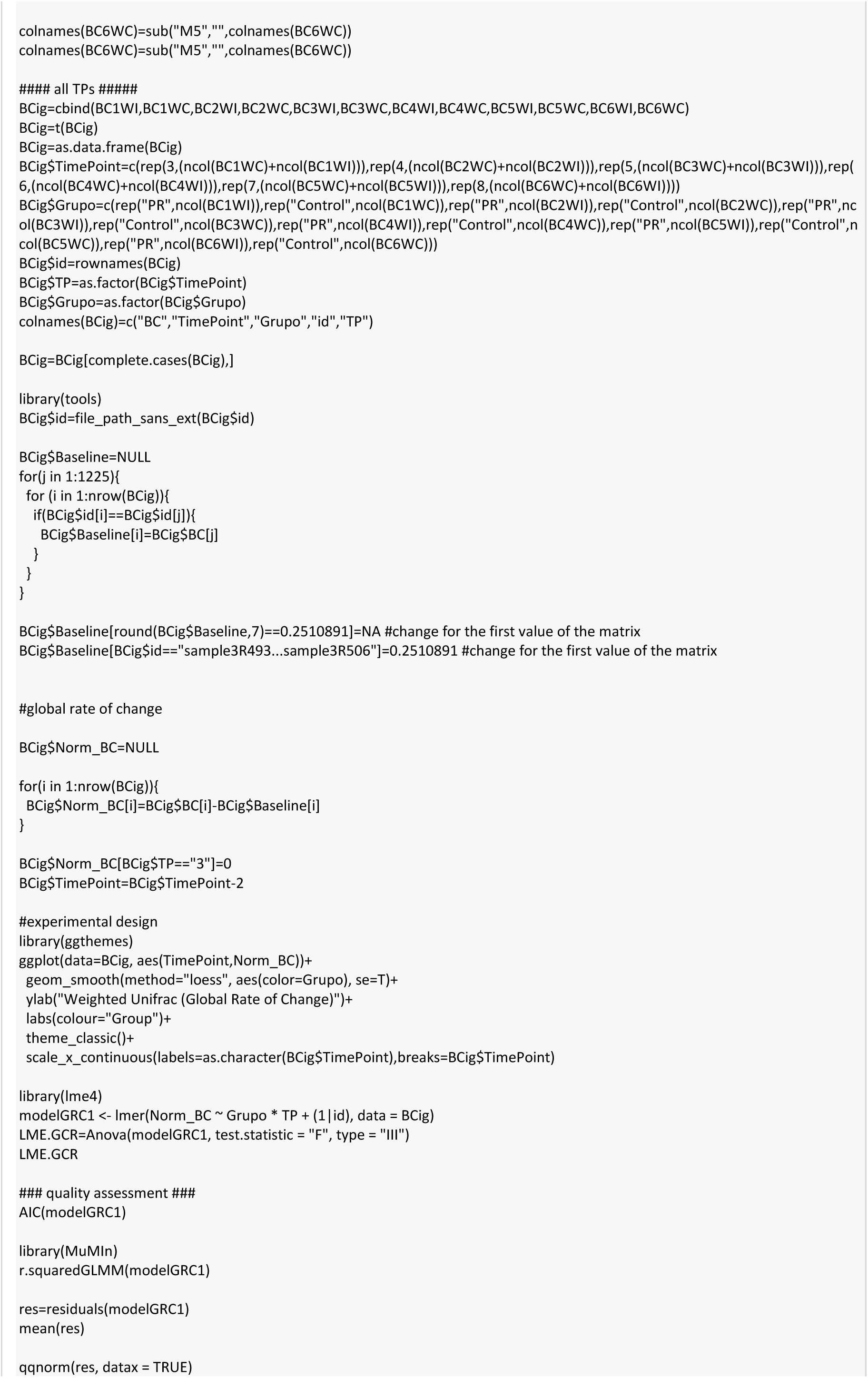
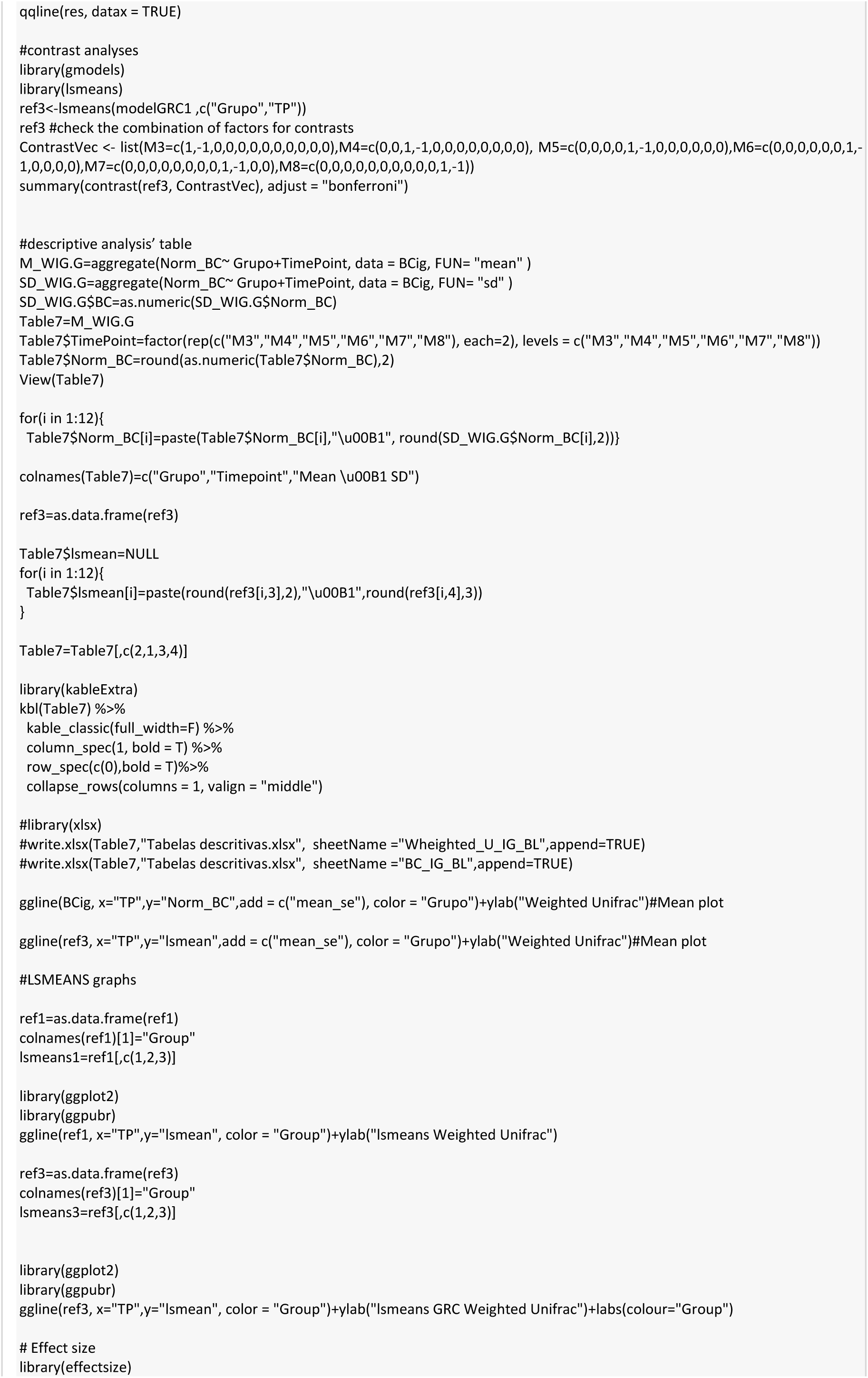
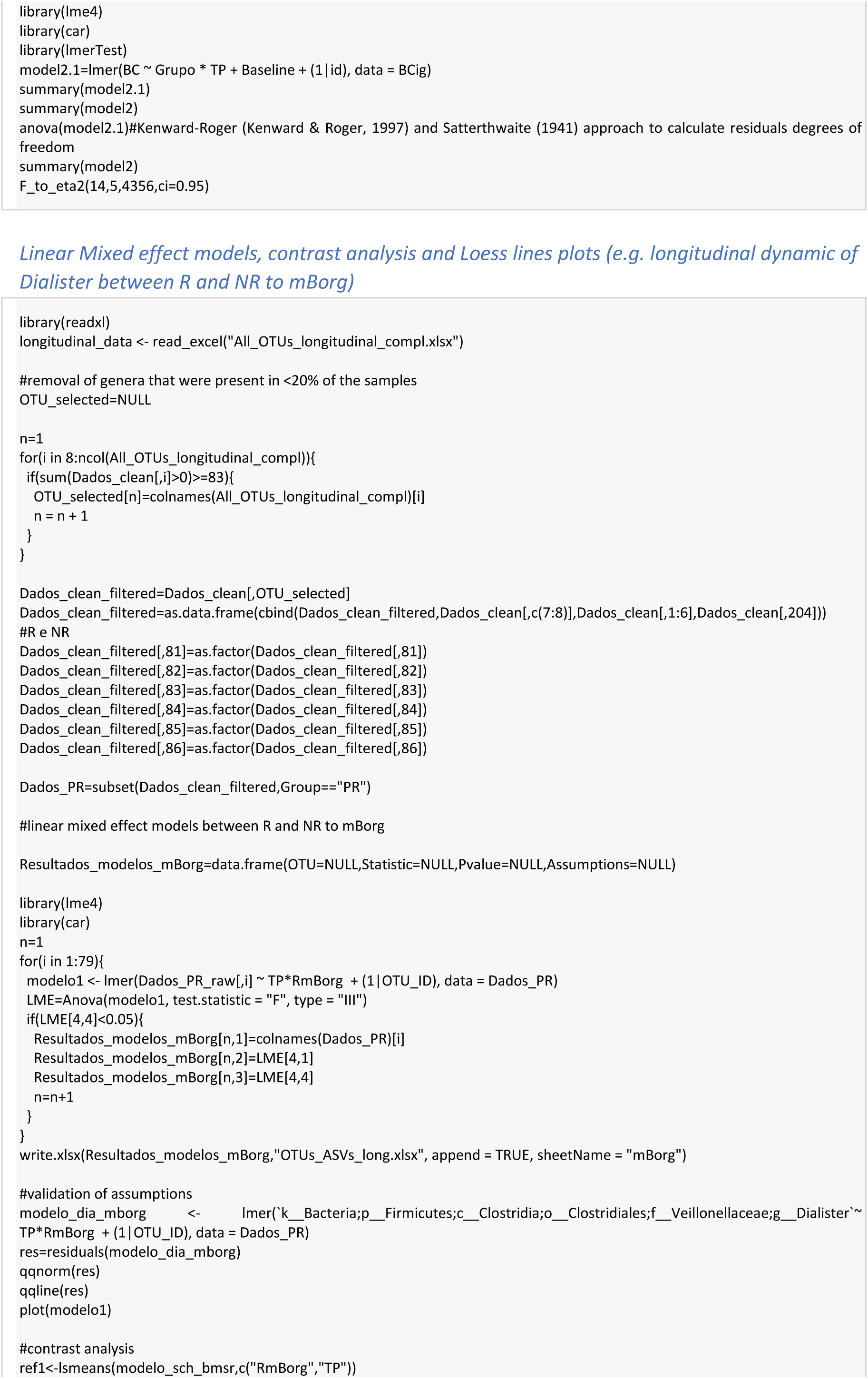
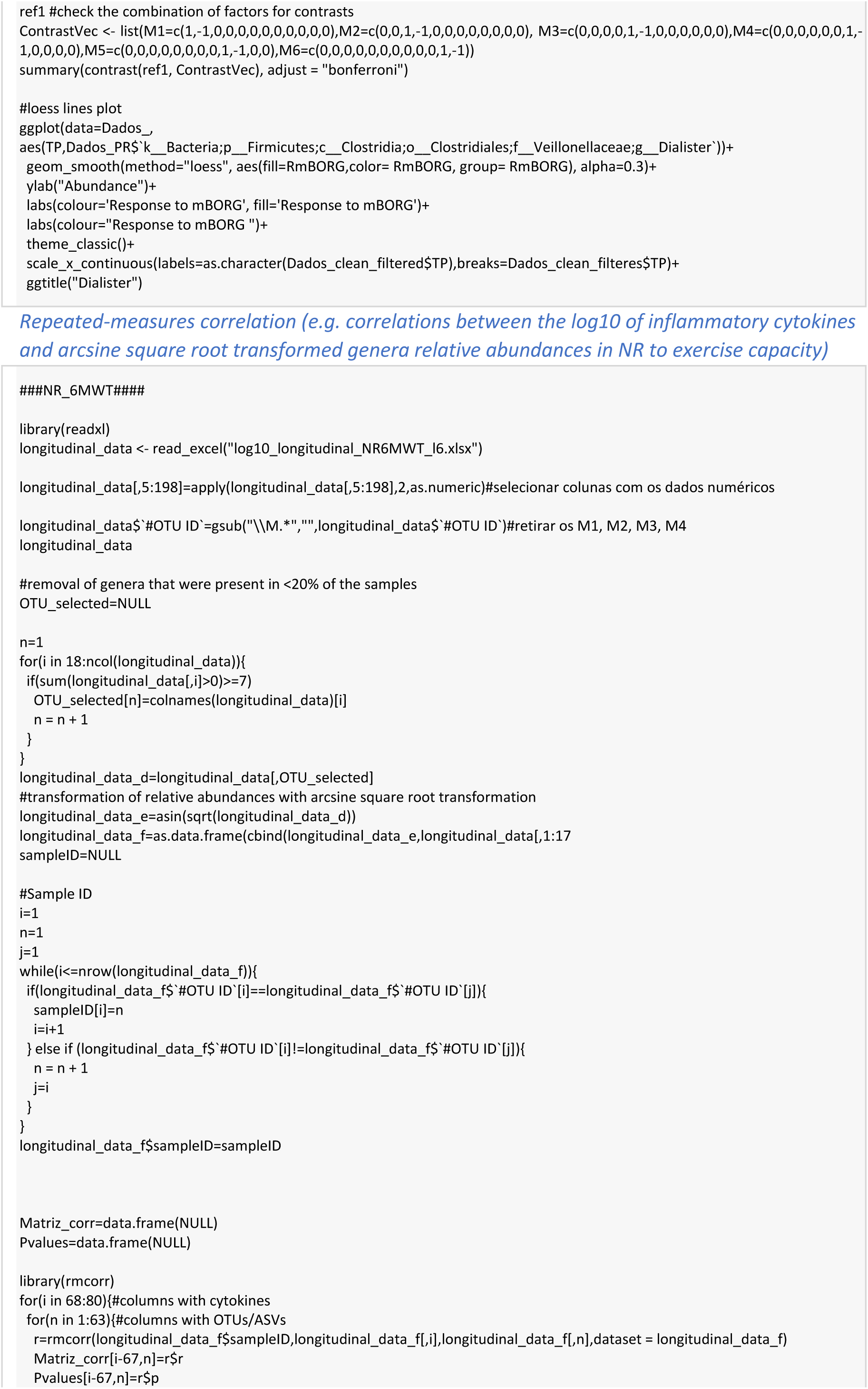
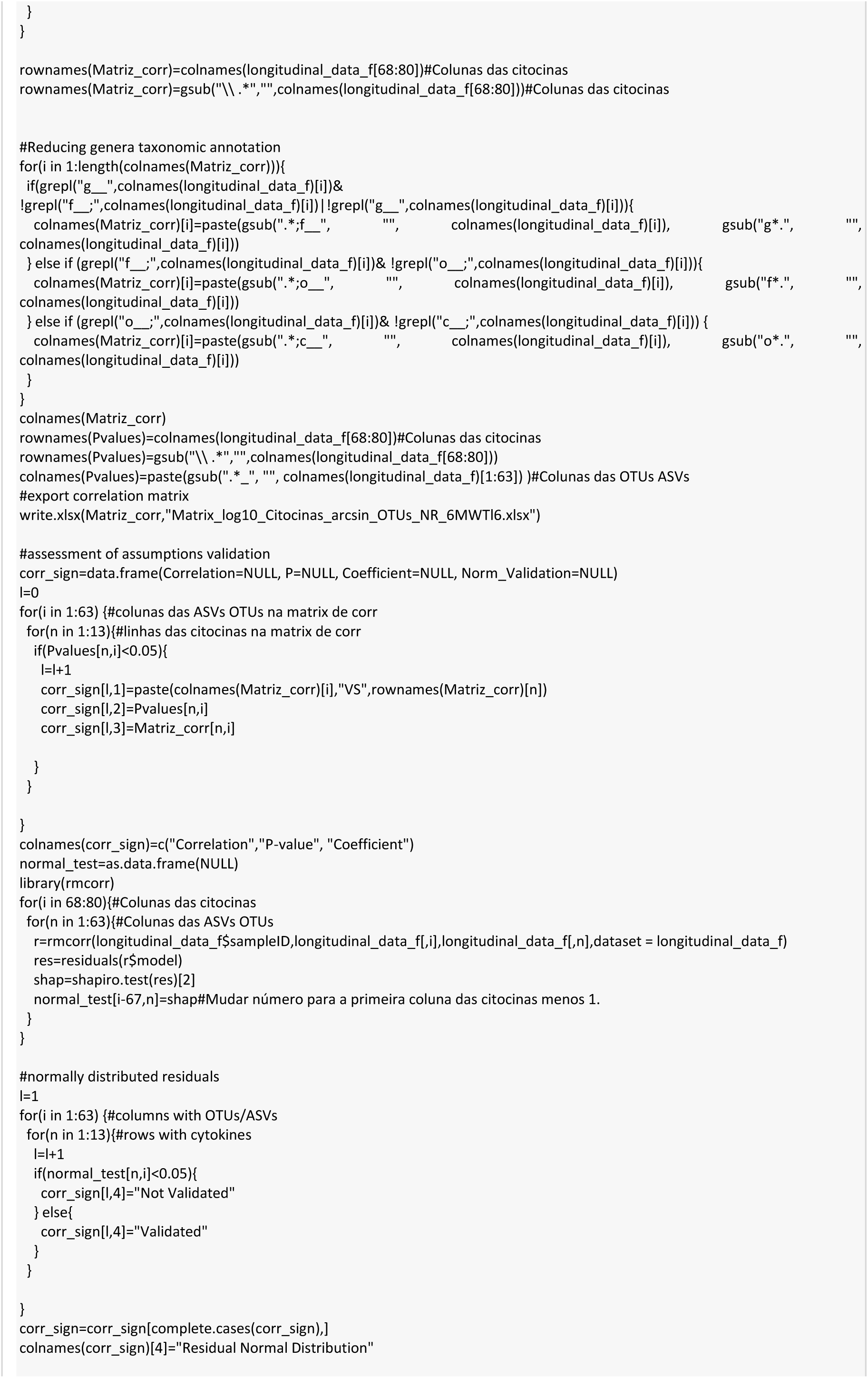
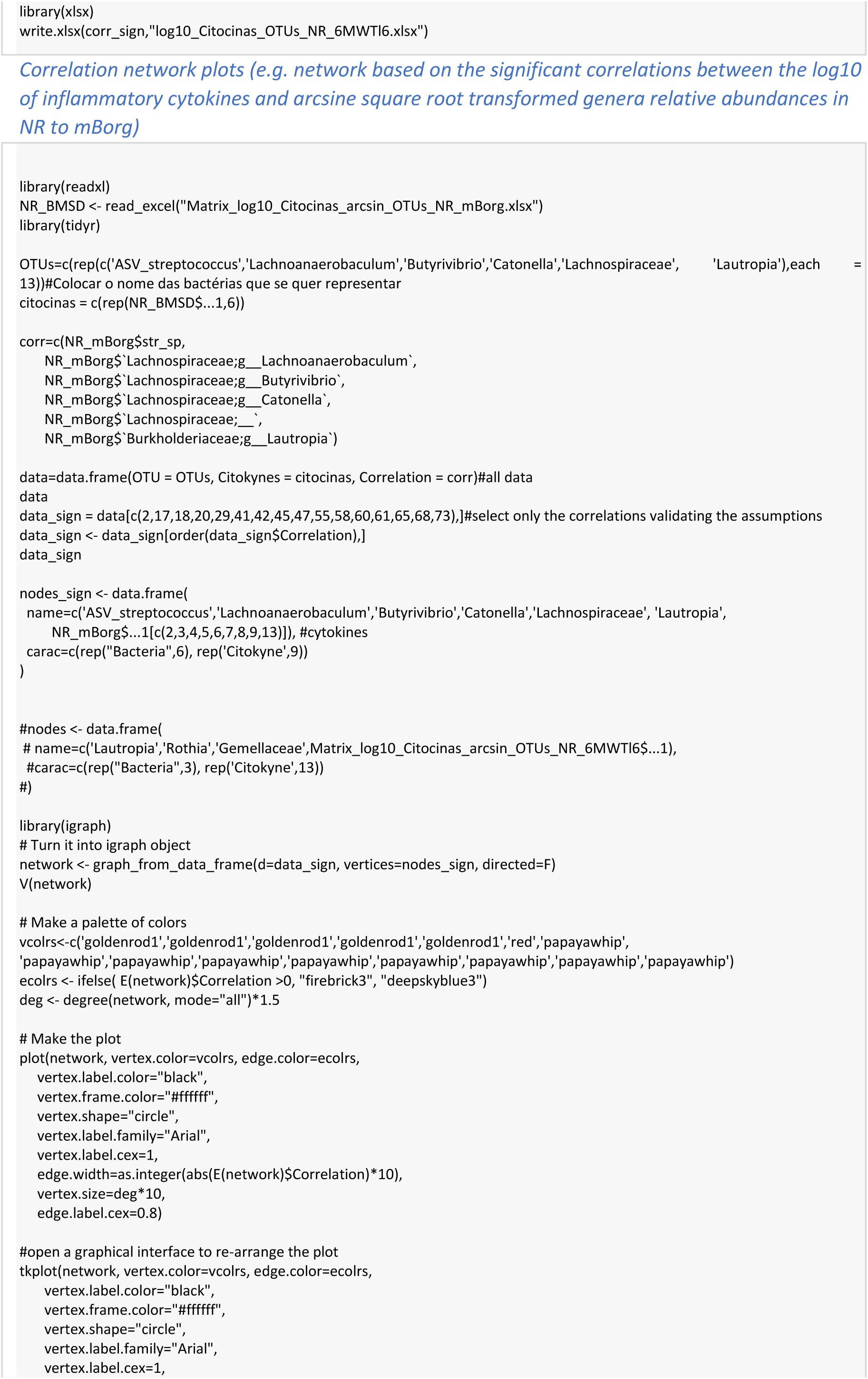
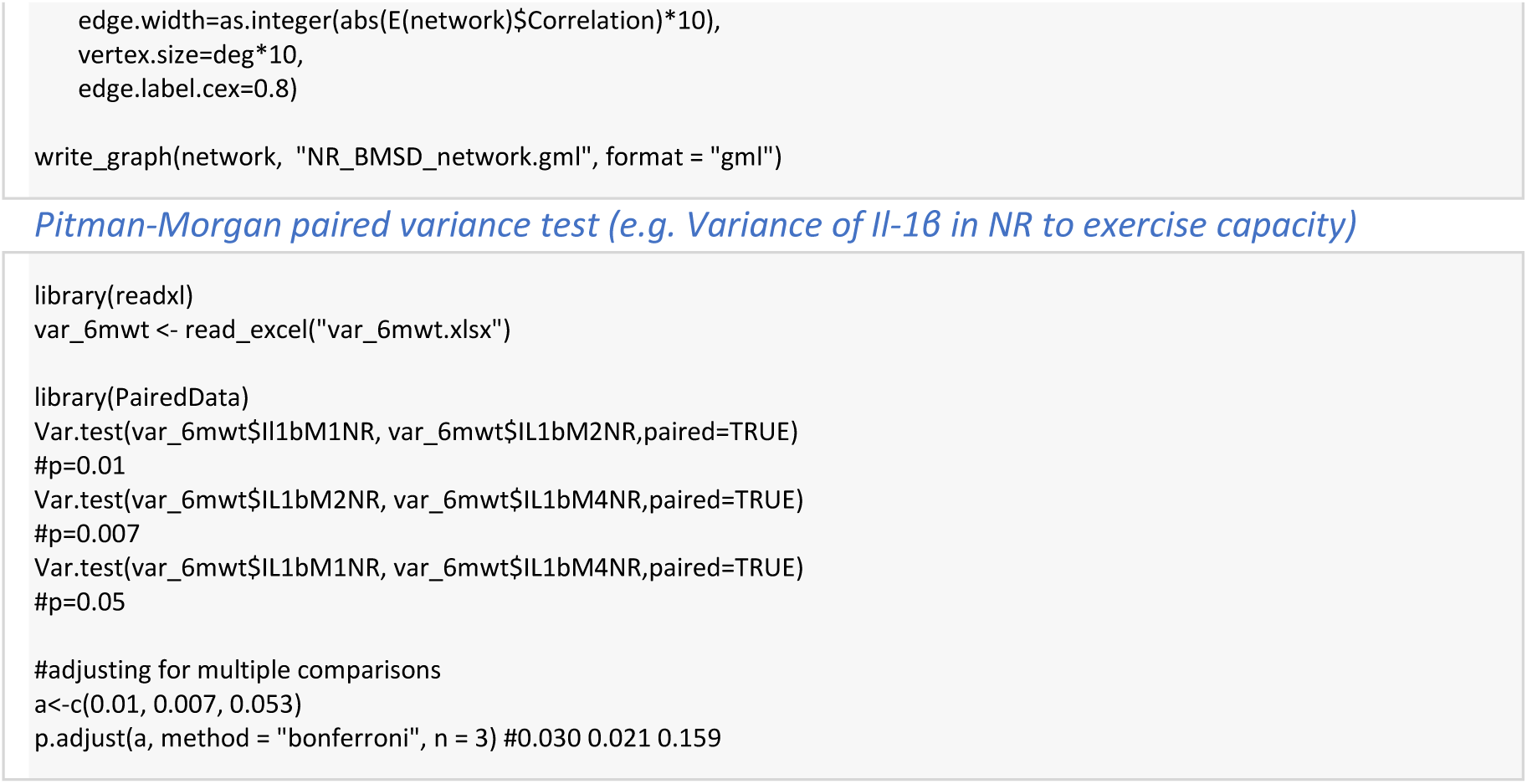

## STROBE Statement—Checklist of items that should be included in reports of cohort studies

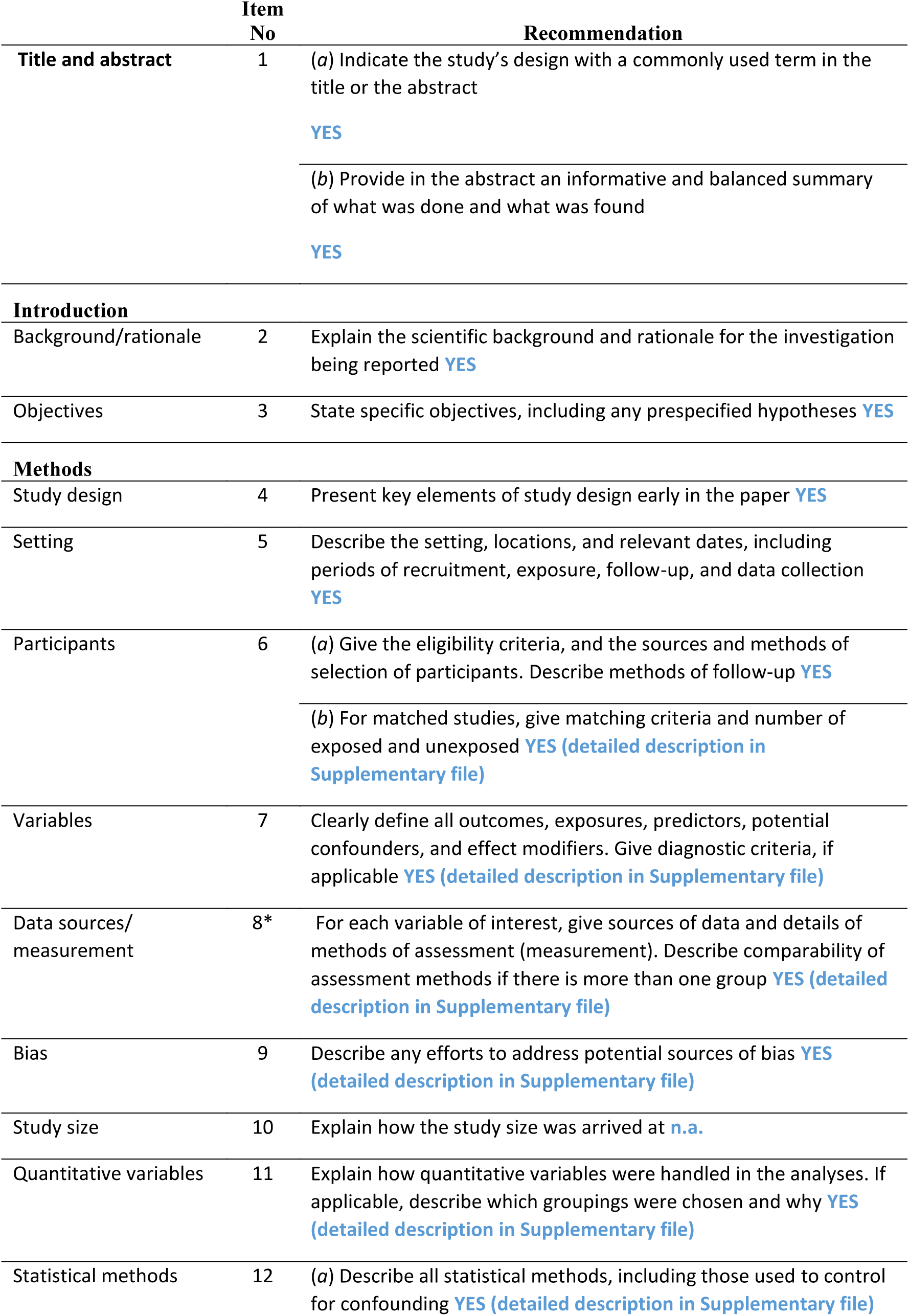
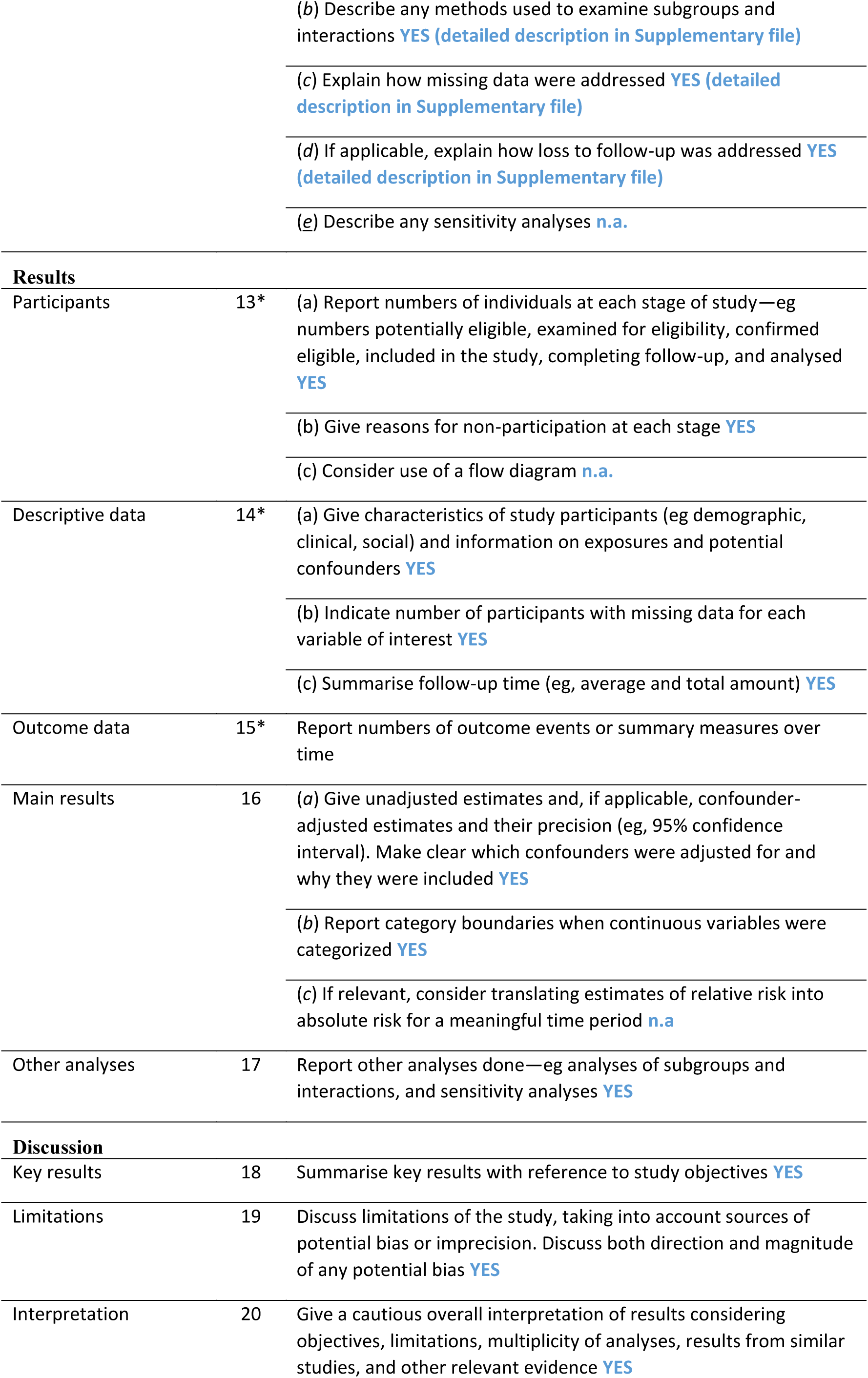
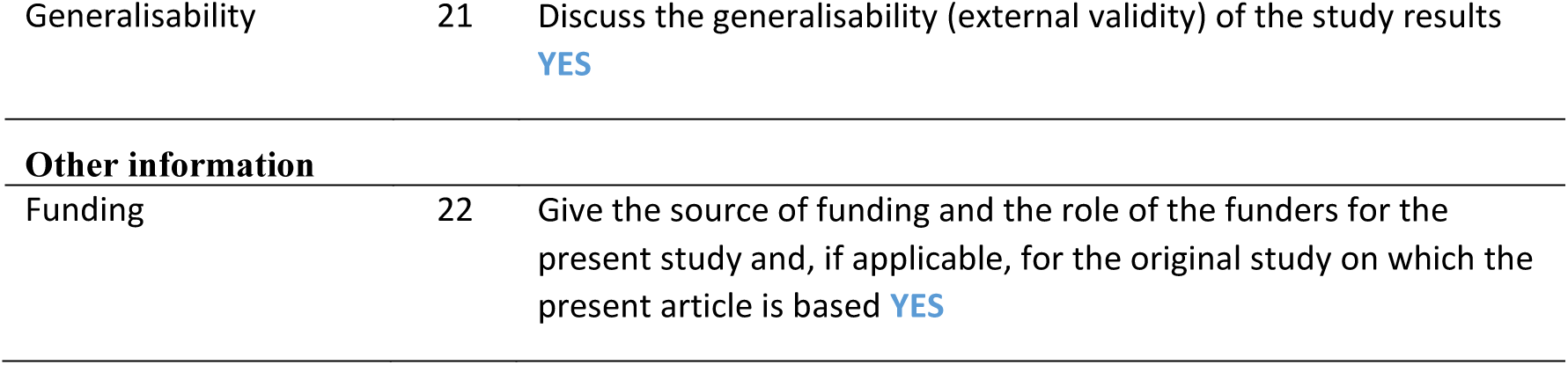

|  | Item No | Recommendation |
| --- | --- | --- |
| Title and abstract | 1 | (a) Indicate the study's design with a commonly used term in the title or the abstract<br>YES |
|  |  | (b) Provide in the abstract an informative and balanced summary of what was done and what was found<br>YES |
| Introduction |  |  |
| Background/rationale | 2 | Explain the scientific background and rationale for the investigation being reported YES |
| Objectives | 3 | State specific objectives, including any prespecified hypotheses YES |
| Methods |  |  |
| Study design | 4 | Present key elements of study design early in the paper YES |
| Setting | 5 | Describe the setting, locations, and relevant dates, including periods of recruitment, exposure, follow-up, and data collection YES |
| Participants | 6 | (a) Give the eligibility criteria, and the sources and methods of selection of participants. Describe methods of follow-up YES |
|  |  | (b) For matched studies, give matching criteria and number of exposed and unexposed YES (detailed description in Supplementary file) |
| Variables | 7 | Clearly define all outcomes, exposures, predictors, potential confounders, and effect modifiers. Give diagnostic criteria, if applicable YES (detailed description in Supplementary file) |
| Data sources/measurement | 8* | For each variable of interest, give sources of data and details of methods of assessment (measurement). Describe comparability of assessment methods if there is more than one group YES (detailed description in Supplementary file) |
| Bias | 9 | Describe any efforts to address potential sources of bias YES (detailed description in Supplementary file) |
| Study size | 10 | Explain how the study size was arrived at n.a. |
| Quantitative variables | 11 | Explain how quantitative variables were handled in the analyses. If applicable, describe which groupings were chosen and why YES (detailed description in Supplementary file) |
| Statistical methods | 12 | (a) Describe all statistical methods, including those used to control for confounding YES (detailed description in Supplementary file) |
|  |  | (b) Describe any methods used to examine subgroups and interactions **YES (detailed description in Supplementary file)** |
|  |  | (c) Explain how missing data were addressed **YES (detailed description in Supplementary file)** |
|  |  | (d) If applicable, explain how loss to follow-up was addressed **YES (detailed description in Supplementary file)** |
|  |  | (e) Describe any sensitivity analyses **n.a.** |
| **Results** |  |  |
| Participants | 13* | (a) Report numbers of individuals at each stage of study—eg numbers potentially eligible, examined for eligibility, confirmed eligible, included in the study, completing follow-up, and analysed **YES** |
|  |  | (b) Give reasons for non-participation at each stage **YES** |
|  |  | (c) Consider use of a flow diagram **n.a.** |
| Descriptive data | 14* | (a) Give characteristics of study participants (eg demographic, clinical, social) and information on exposures and potential confounders **YES** |
|  |  | (b) Indicate number of participants with missing data for each variable of interest **YES** |
|  |  | (c) Summarise follow-up time (eg, average and total amount) **YES** |
| Outcome data | 15* | Report numbers of outcome events or summary measures over time |
| Main results | 16 | (a) Give unadjusted estimates and, if applicable, confounder-adjusted estimates and their precision (eg, 95% confidence interval). Make clear which confounders were adjusted for and why they were included **YES** |
|  |  | (b) Report category boundaries when continuous variables were categorized **YES** |
|  |  | (c) If relevant, consider translating estimates of relative risk into absolute risk for a meaningful time period **n.a** |
| Other analyses | 17 | Report other analyses done—eg analyses of subgroups and interactions, and sensitivity analyses **YES** |
| **Discussion** |  |  |
| Key results | 18 | Summarise key results with reference to study objectives **YES** |
| Limitations | 19 | Discuss limitations of the study, taking into account sources of potential bias or imprecision. Discuss both direction and magnitude of any potential bias **YES** |
| Interpretation | 20 | Give a cautious overall interpretation of results considering objectives, limitations, multiplicity of analyses, results from similar studies, and other relevant evidence **YES** |
| Generalisability | 21 | Discuss the generalisability (external validity) of the study results<br>**YES** |
| **Other information** |  |  |
| Funding | 22 | Give the source of funding and the role of the funders for the present study and, if applicable, for the original study on which the present article is based **YES** |

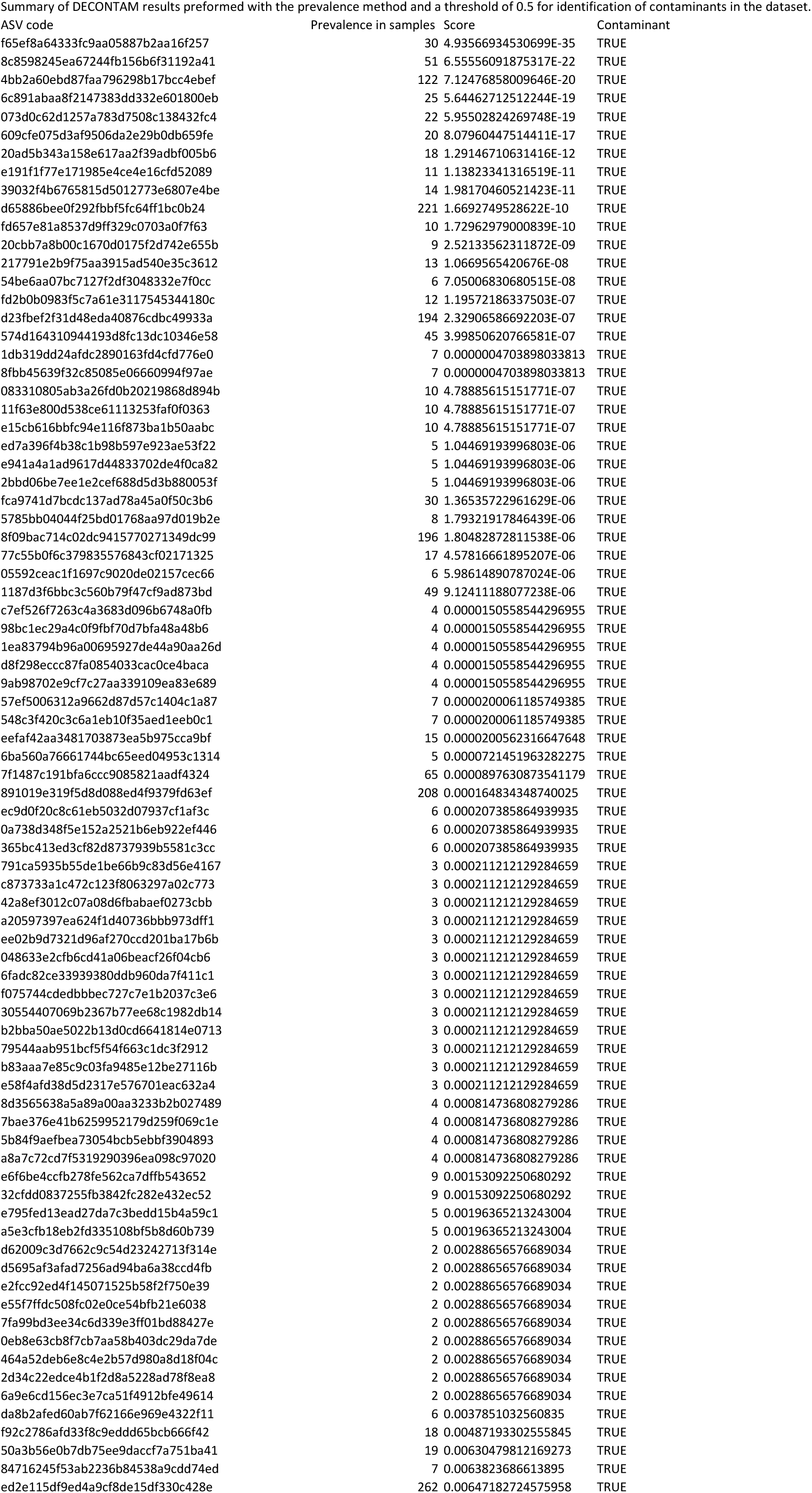
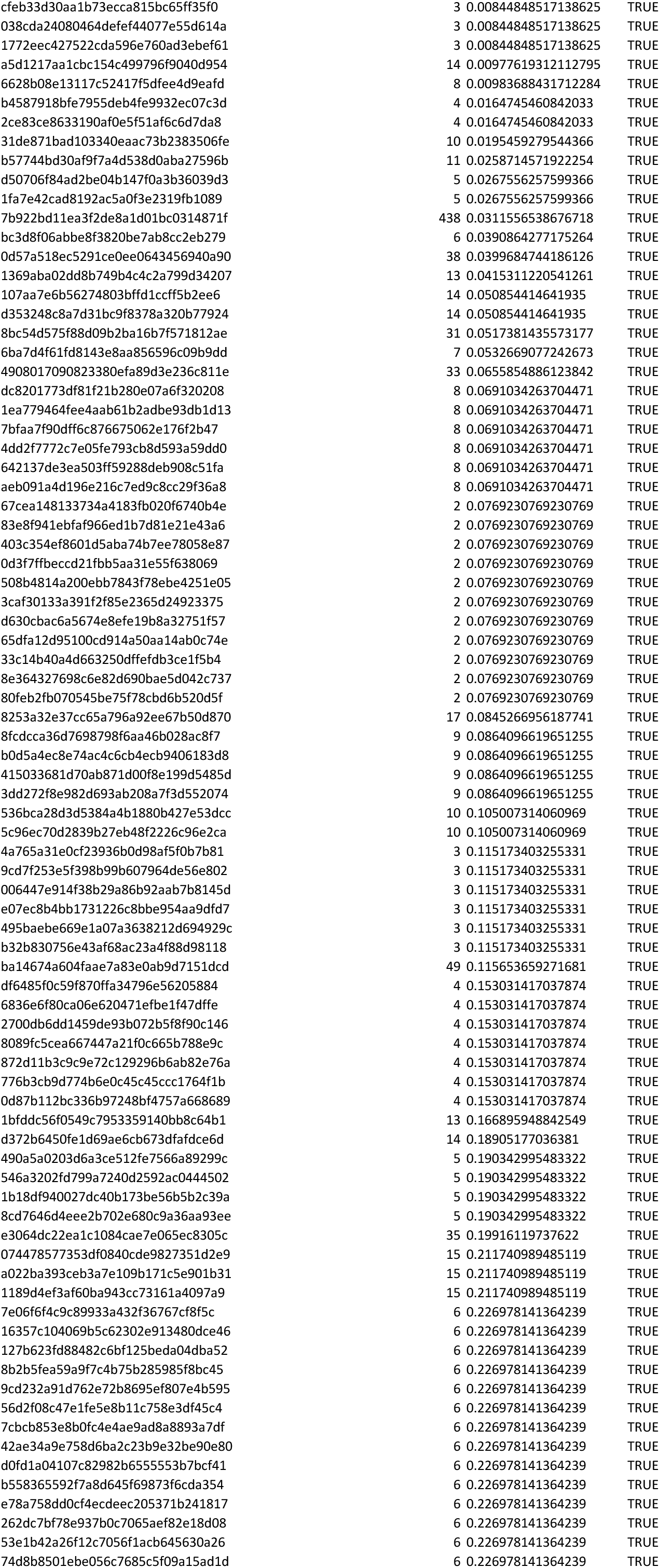
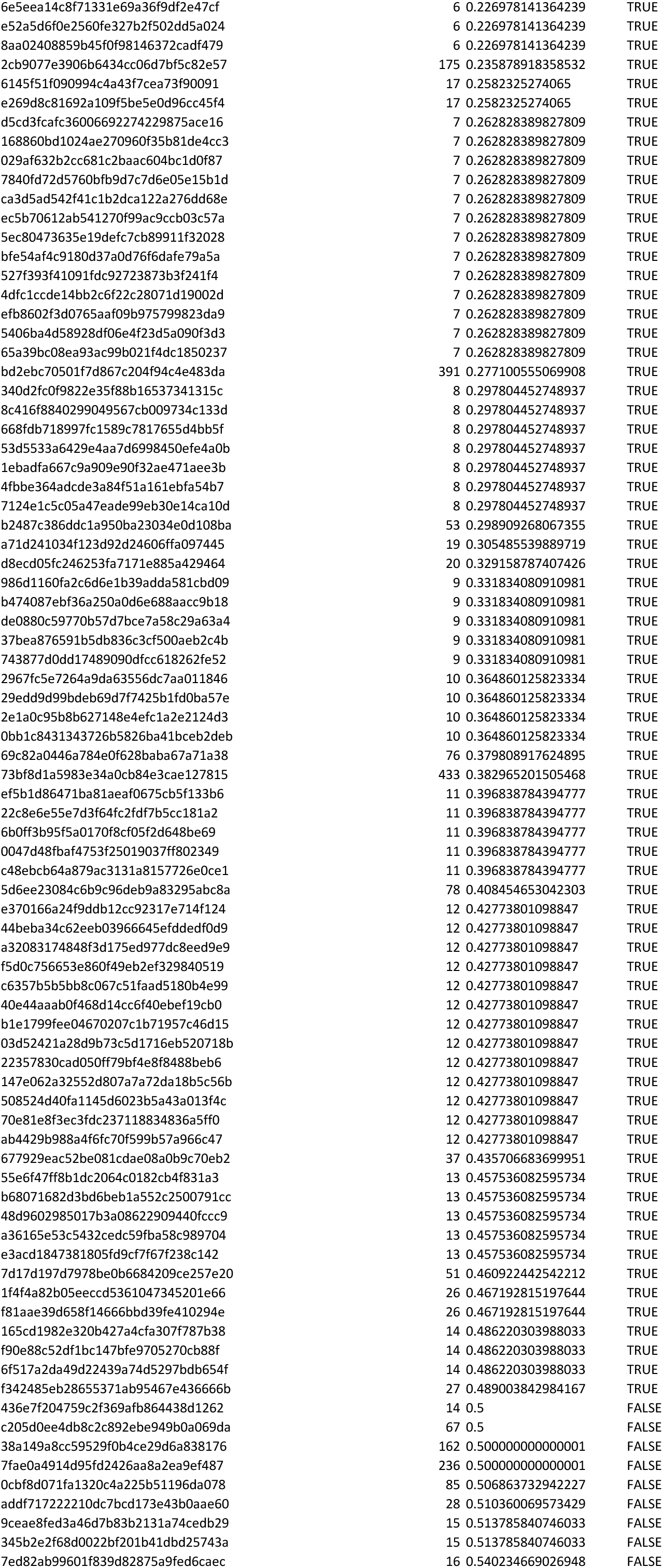
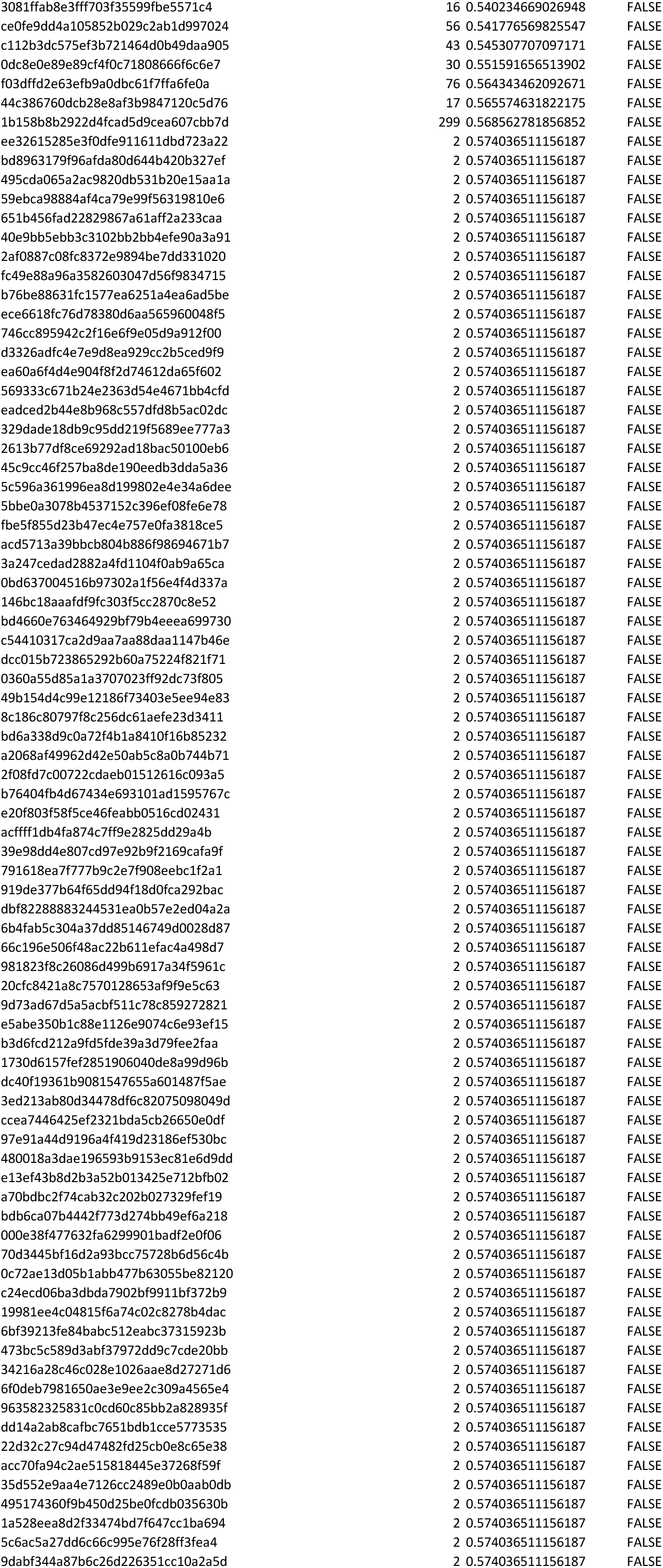
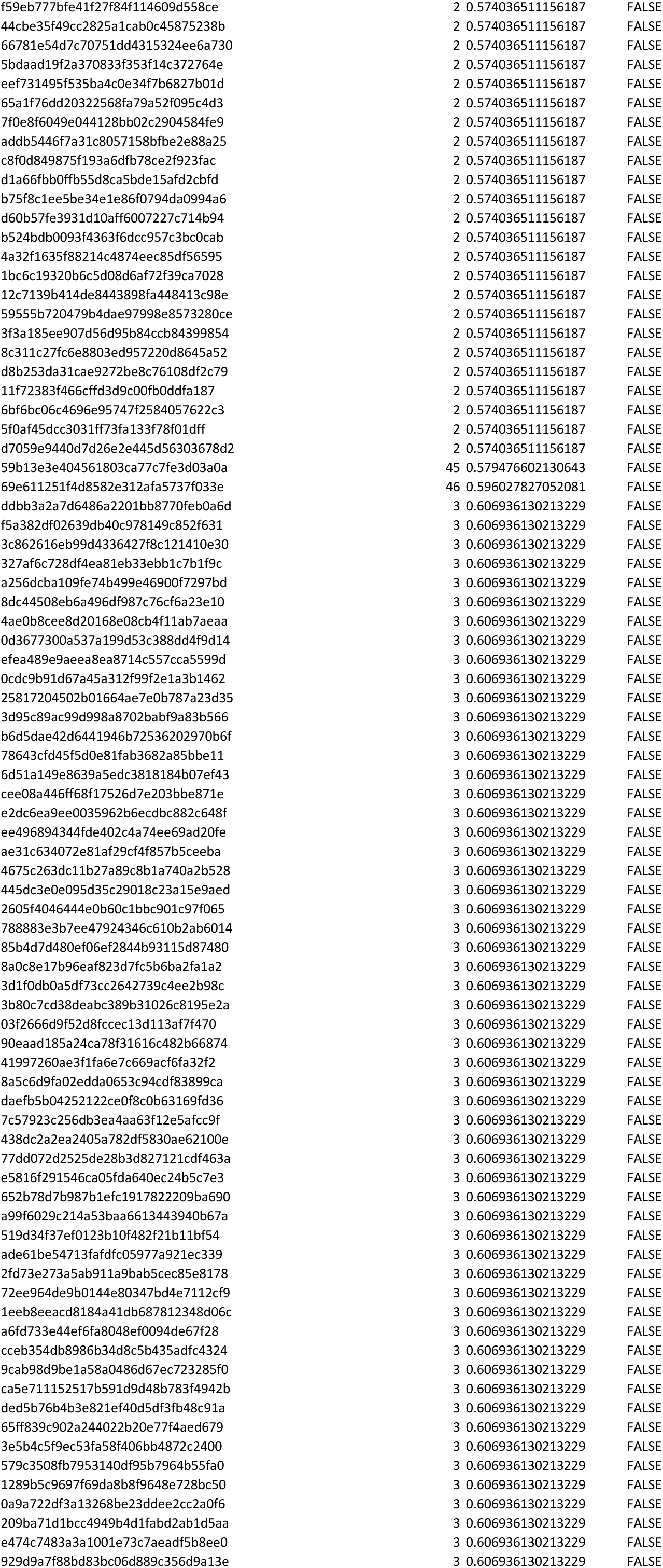
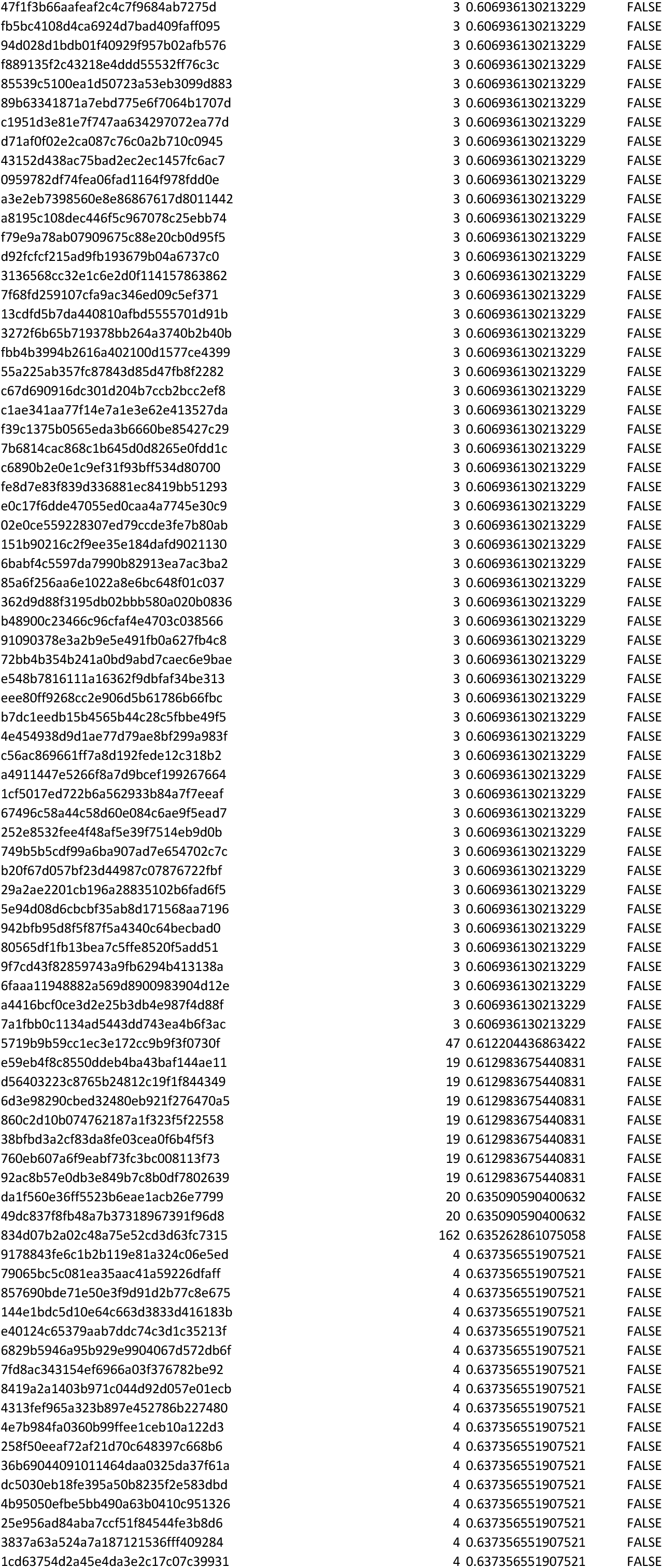
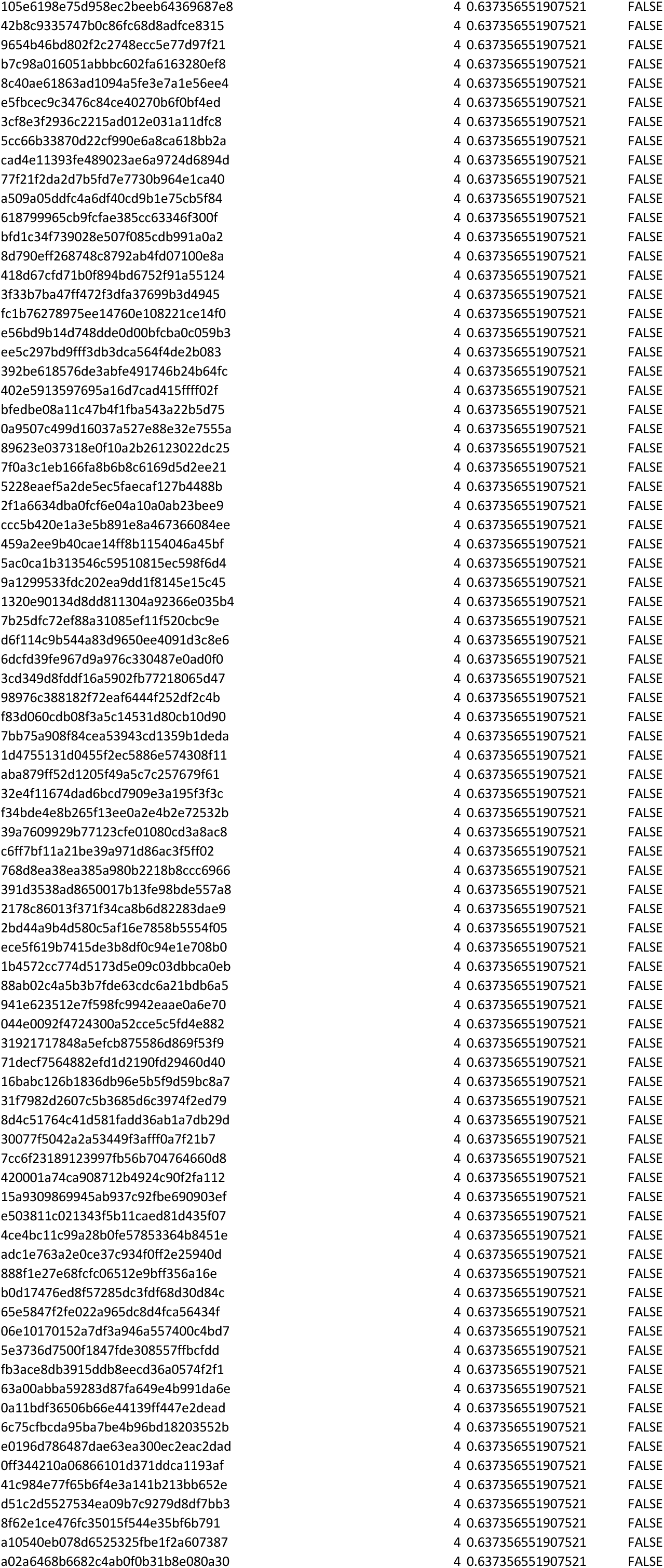
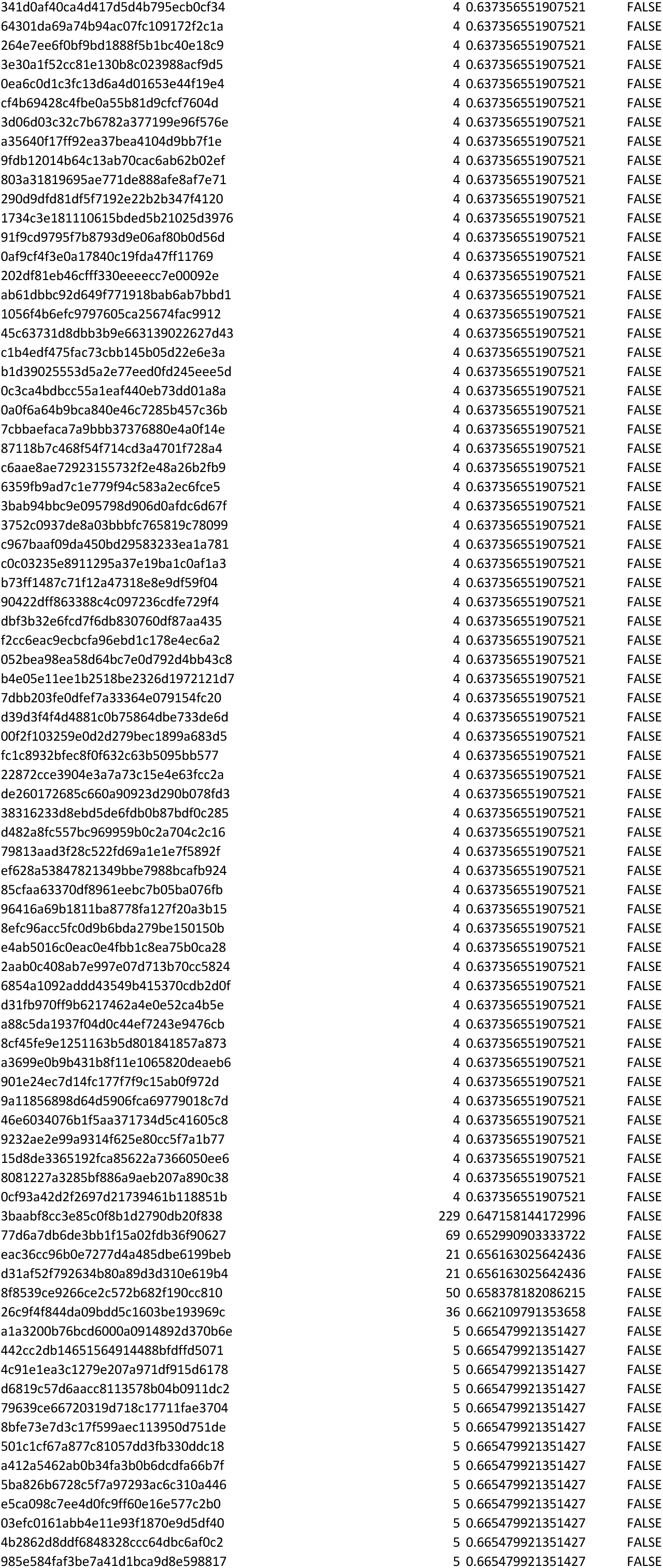
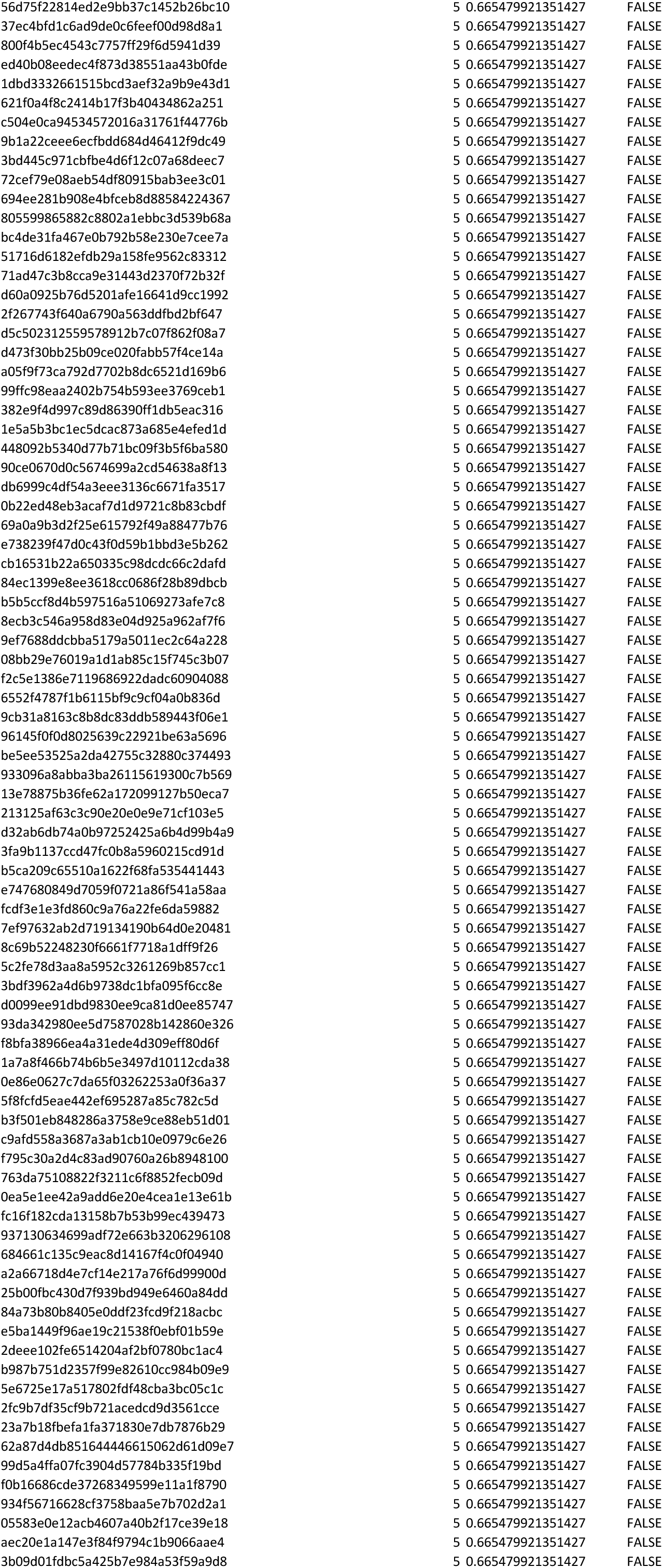
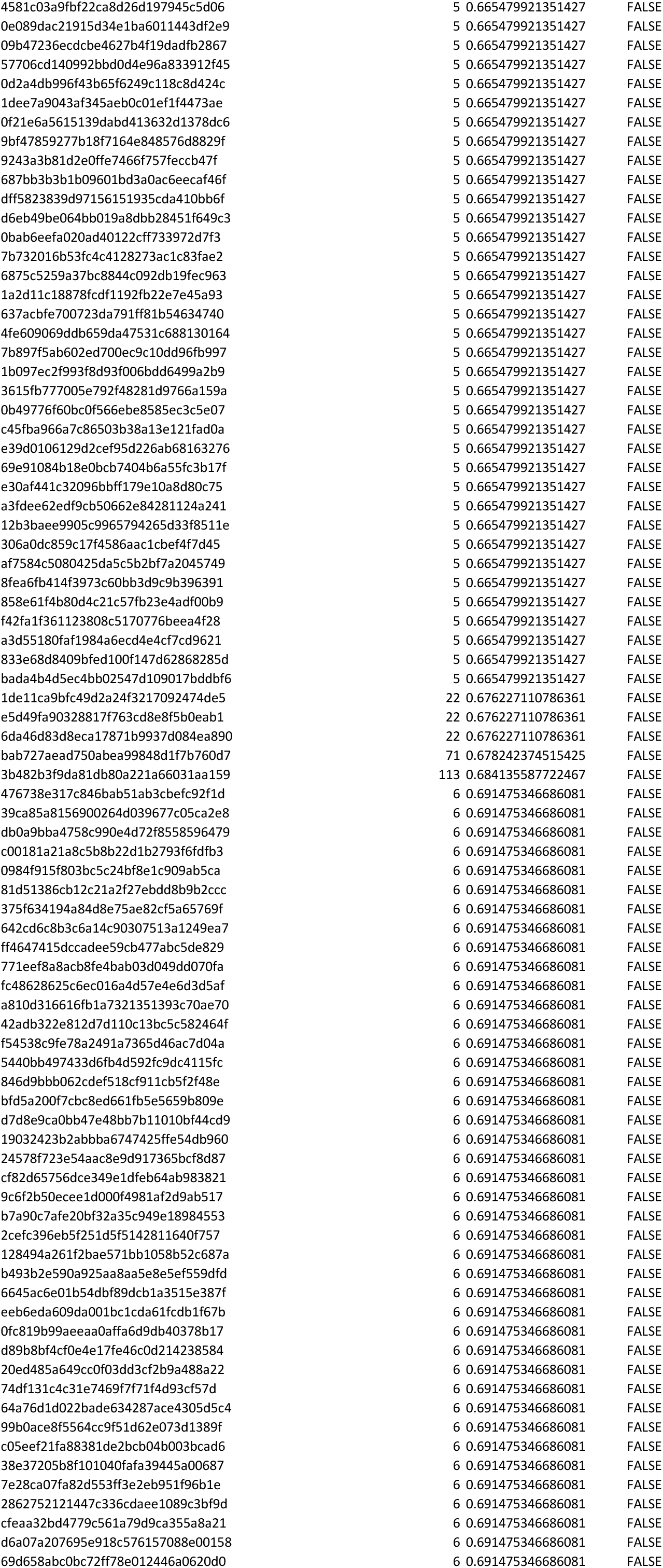
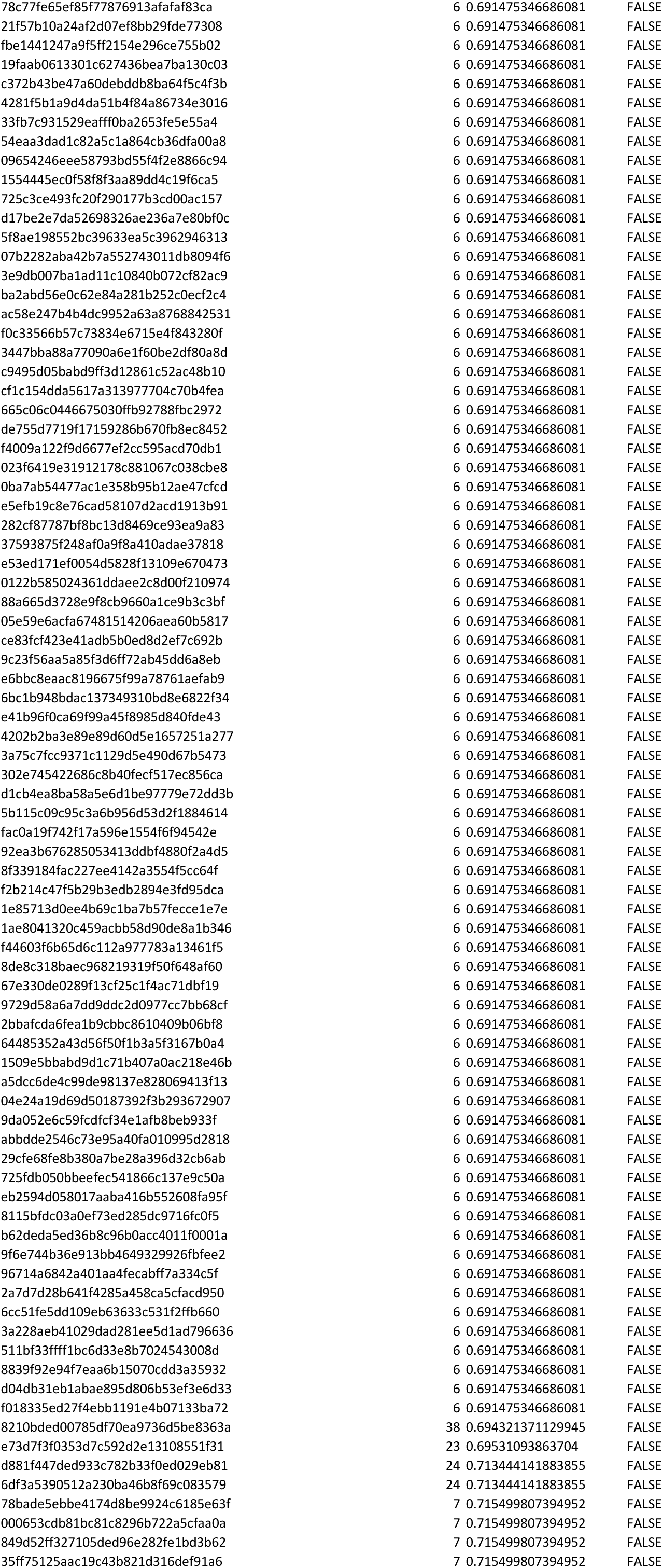
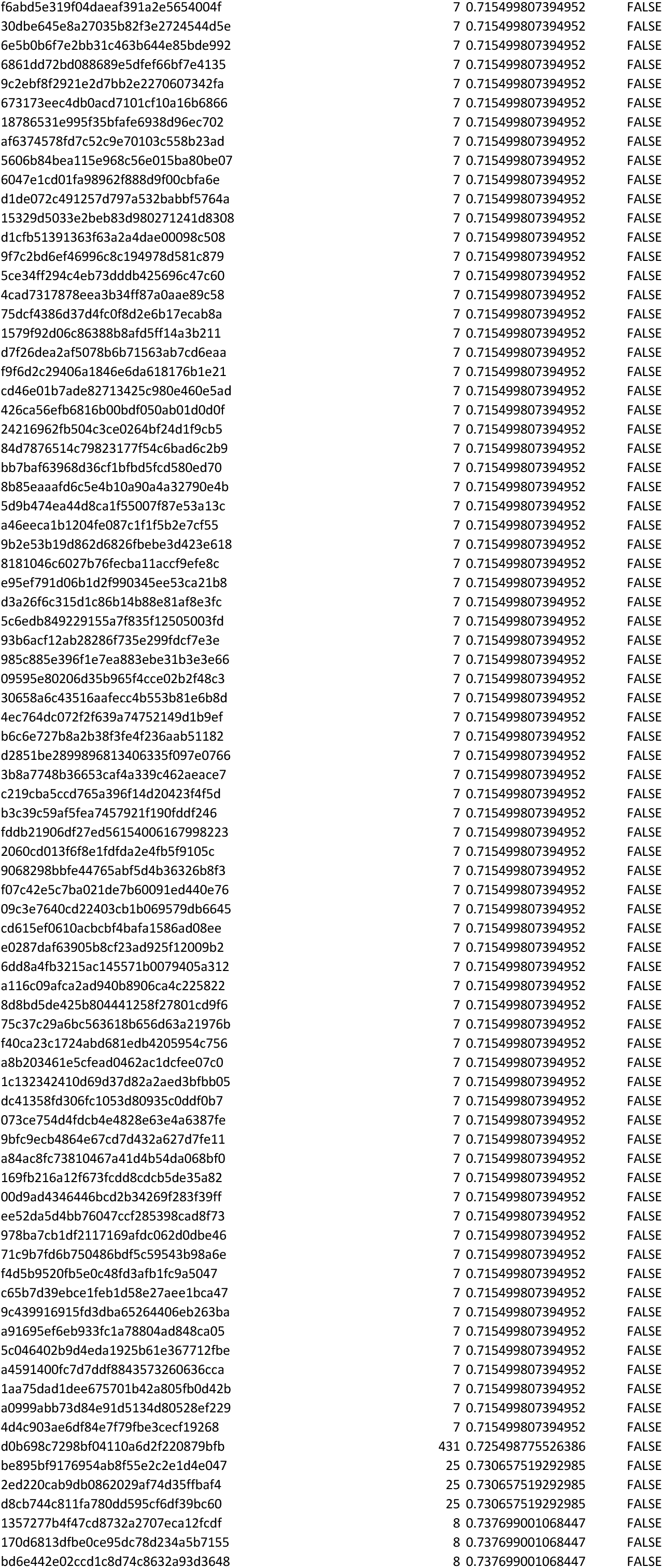
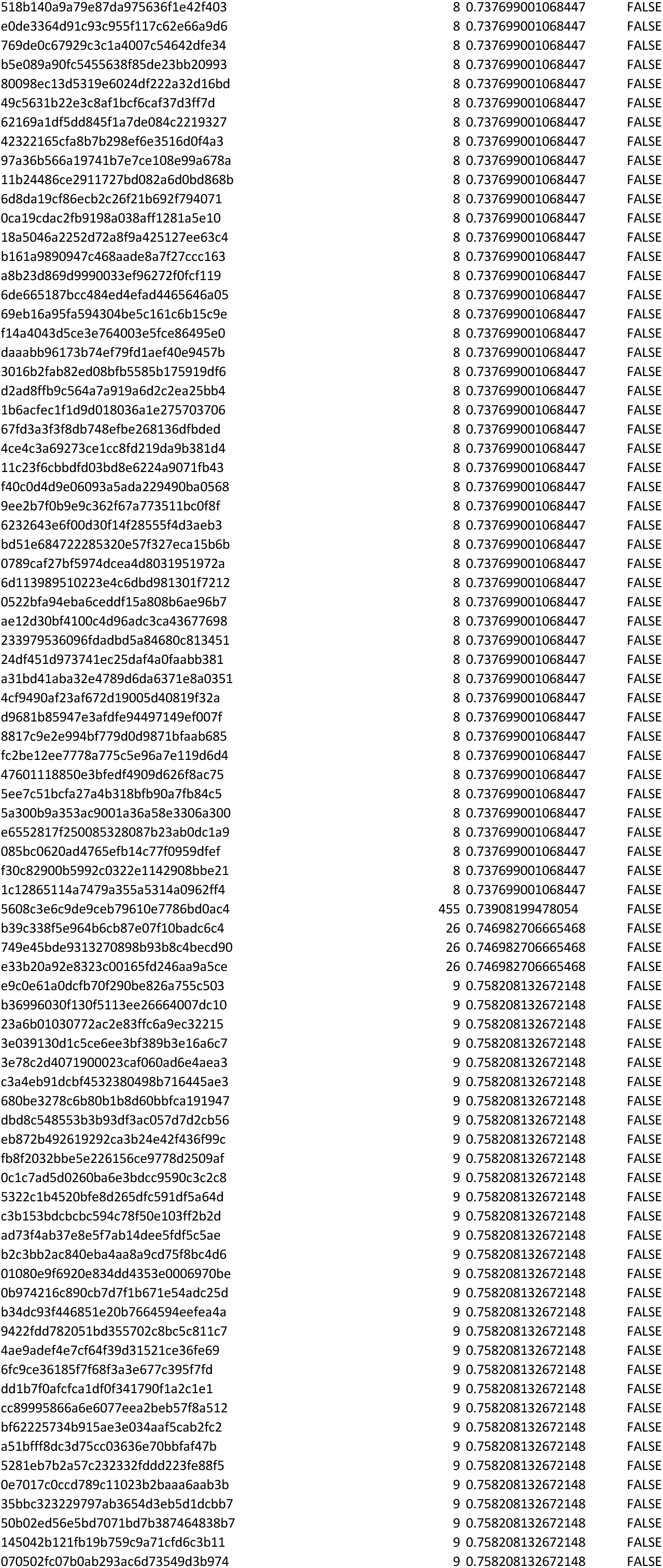
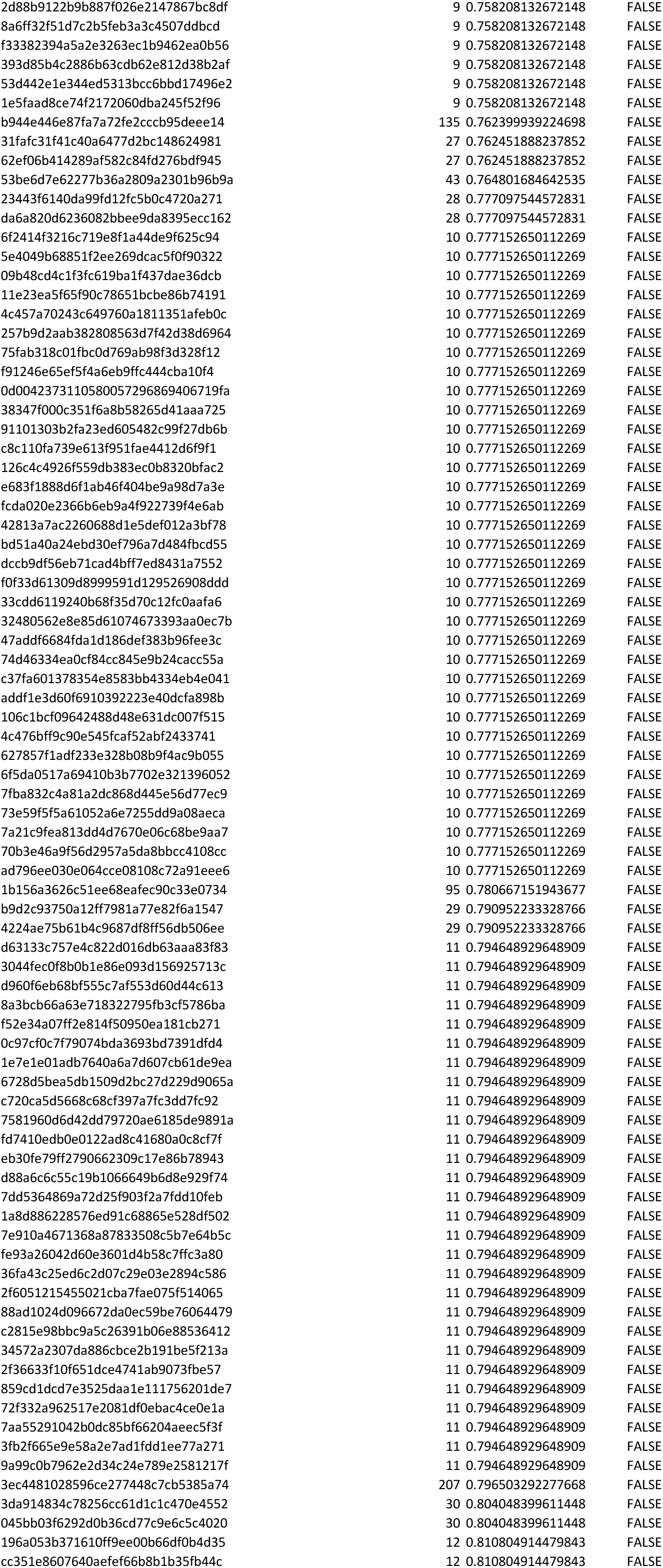
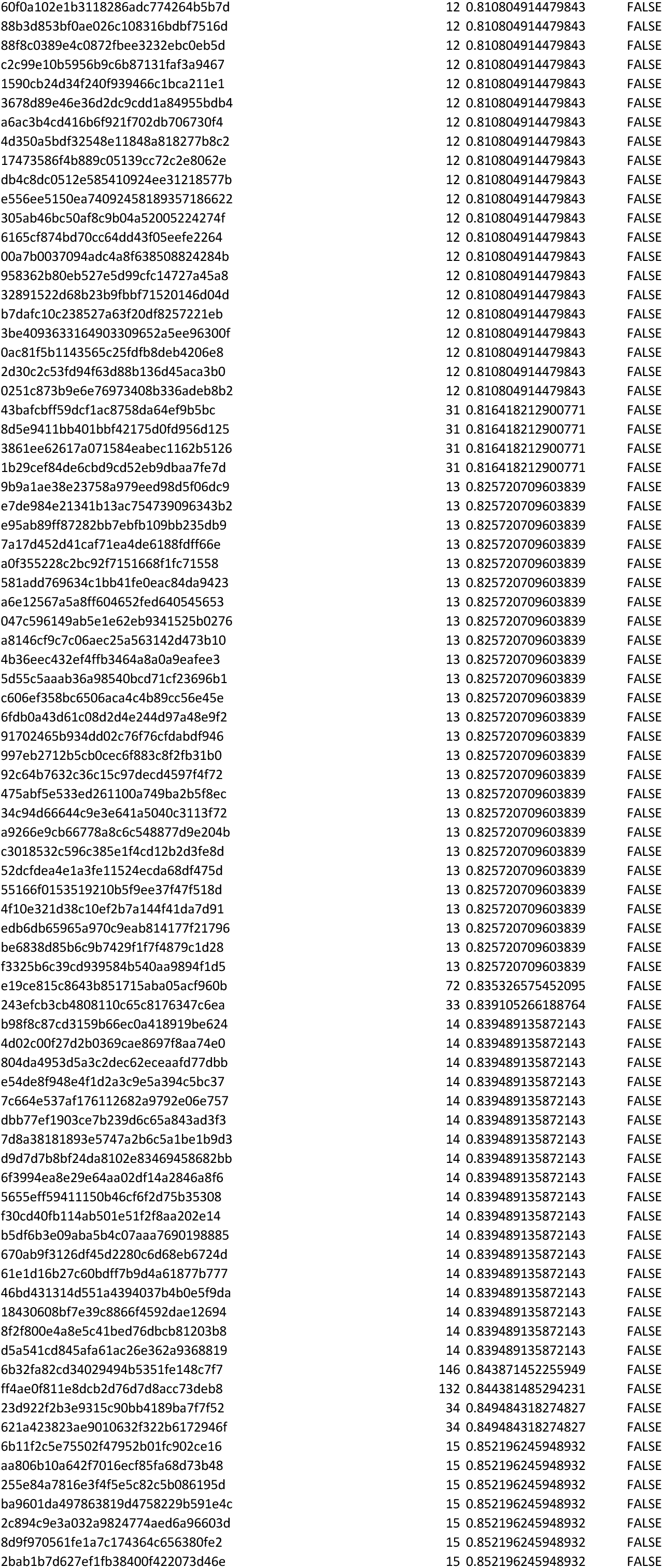
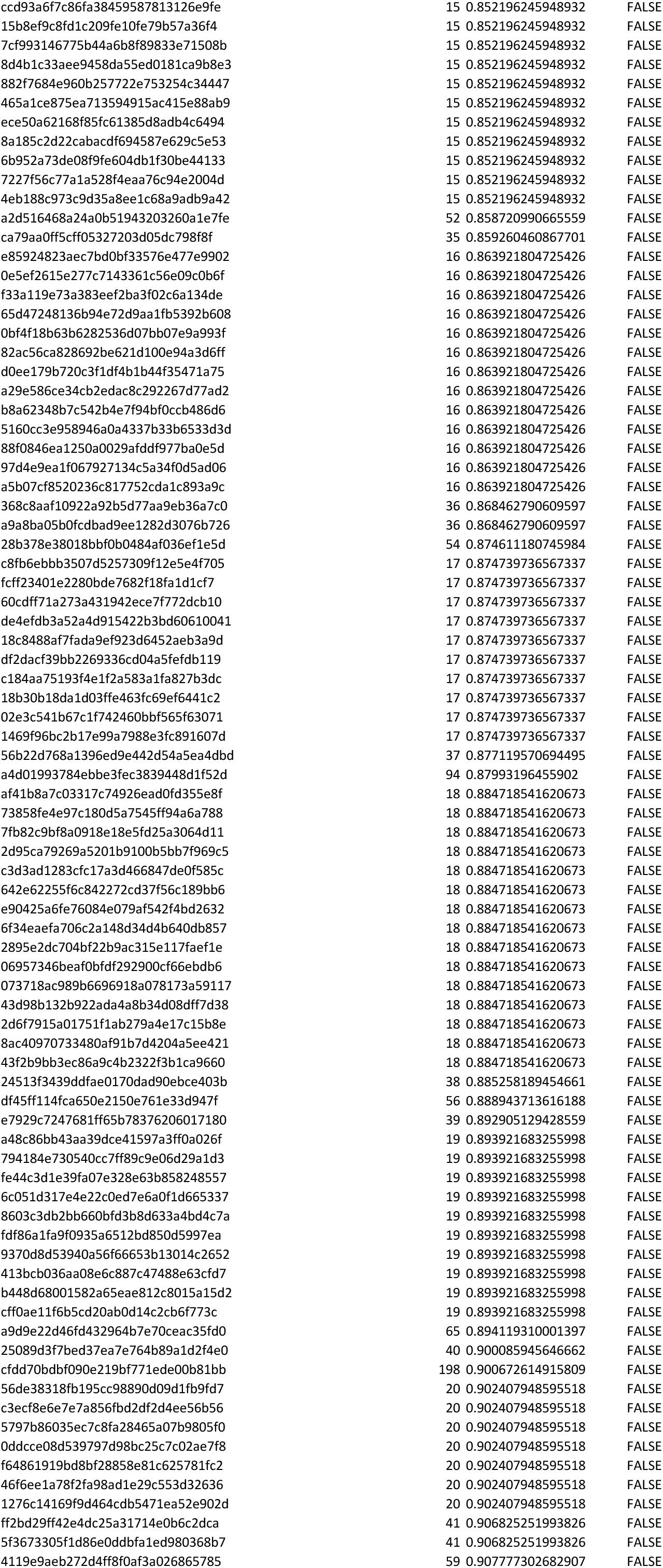
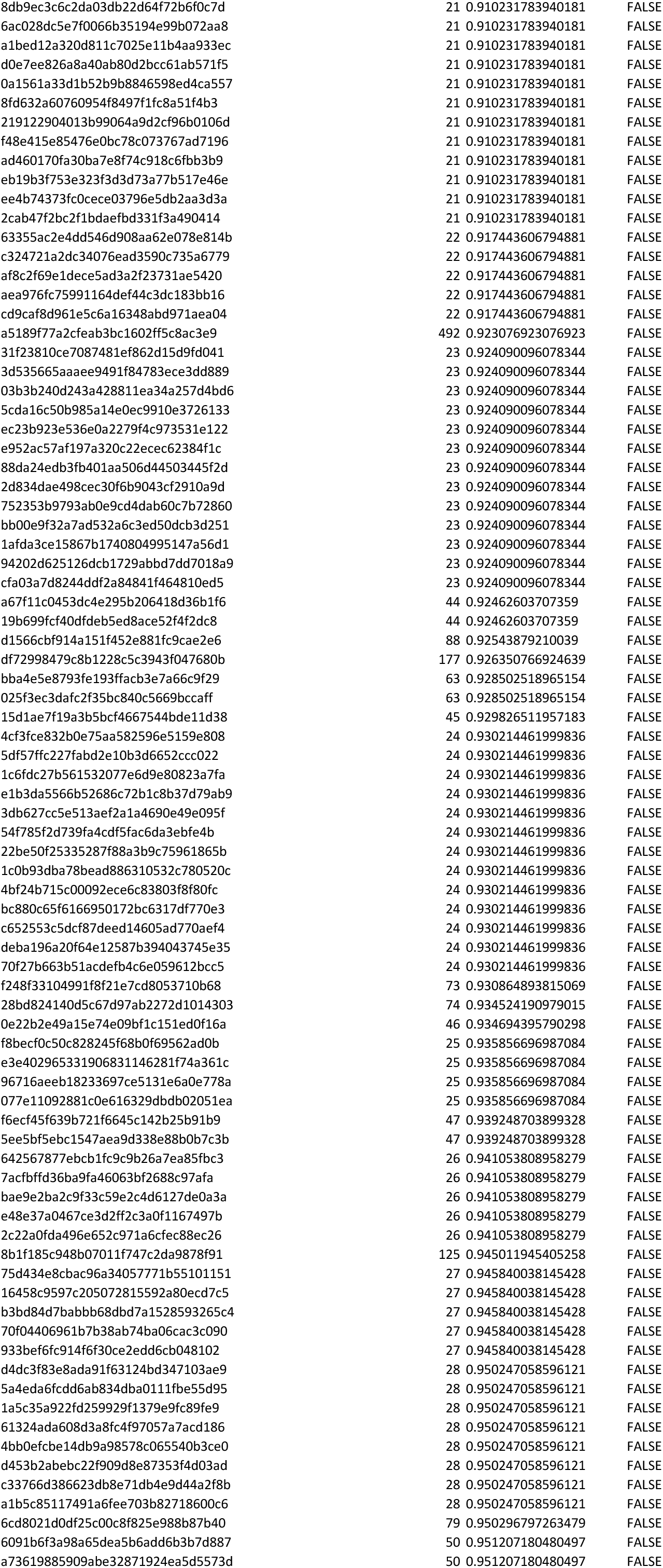
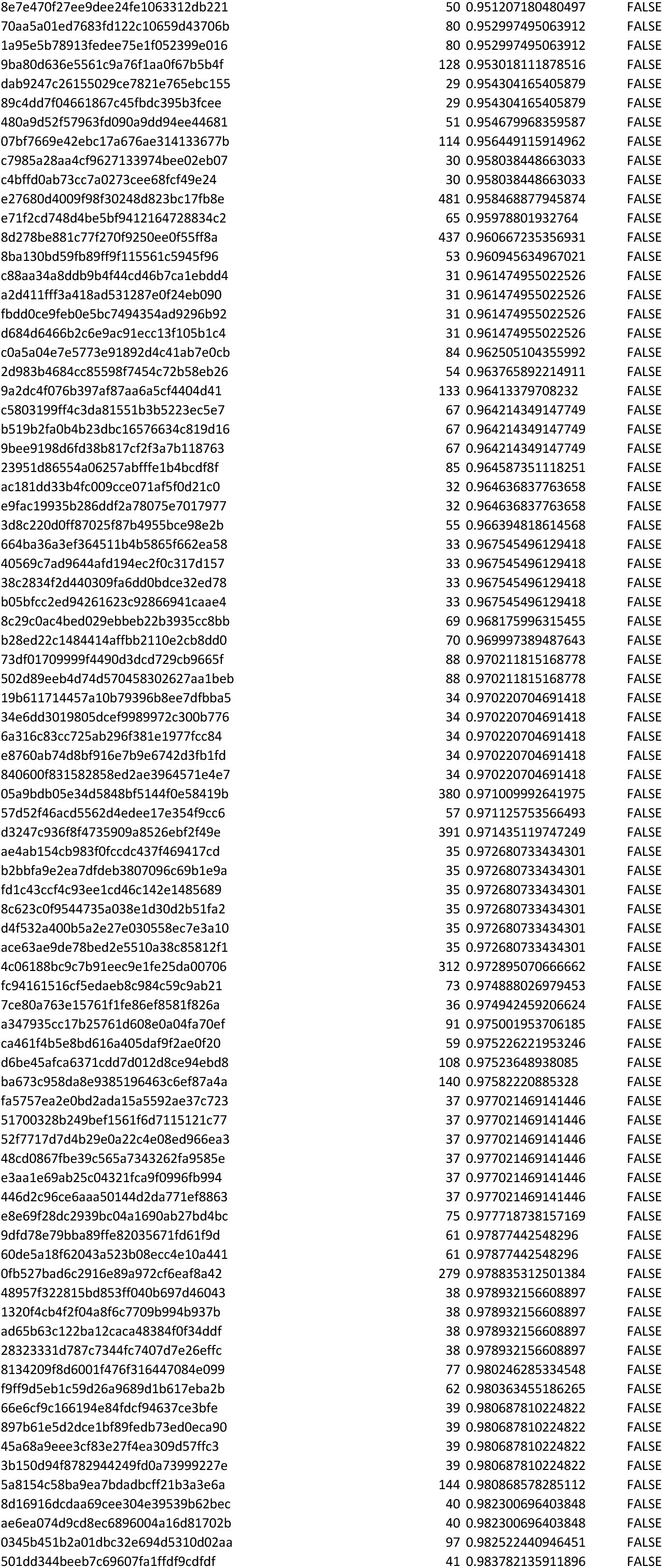
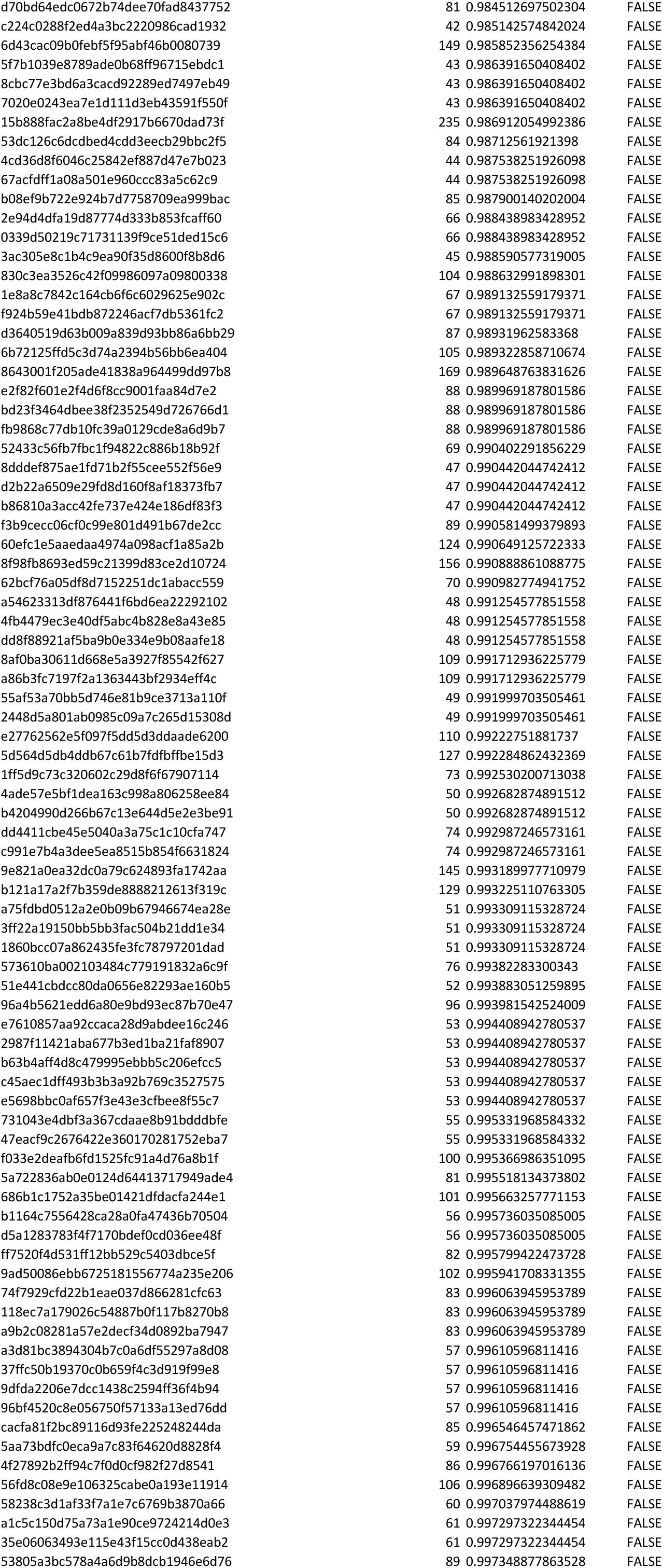
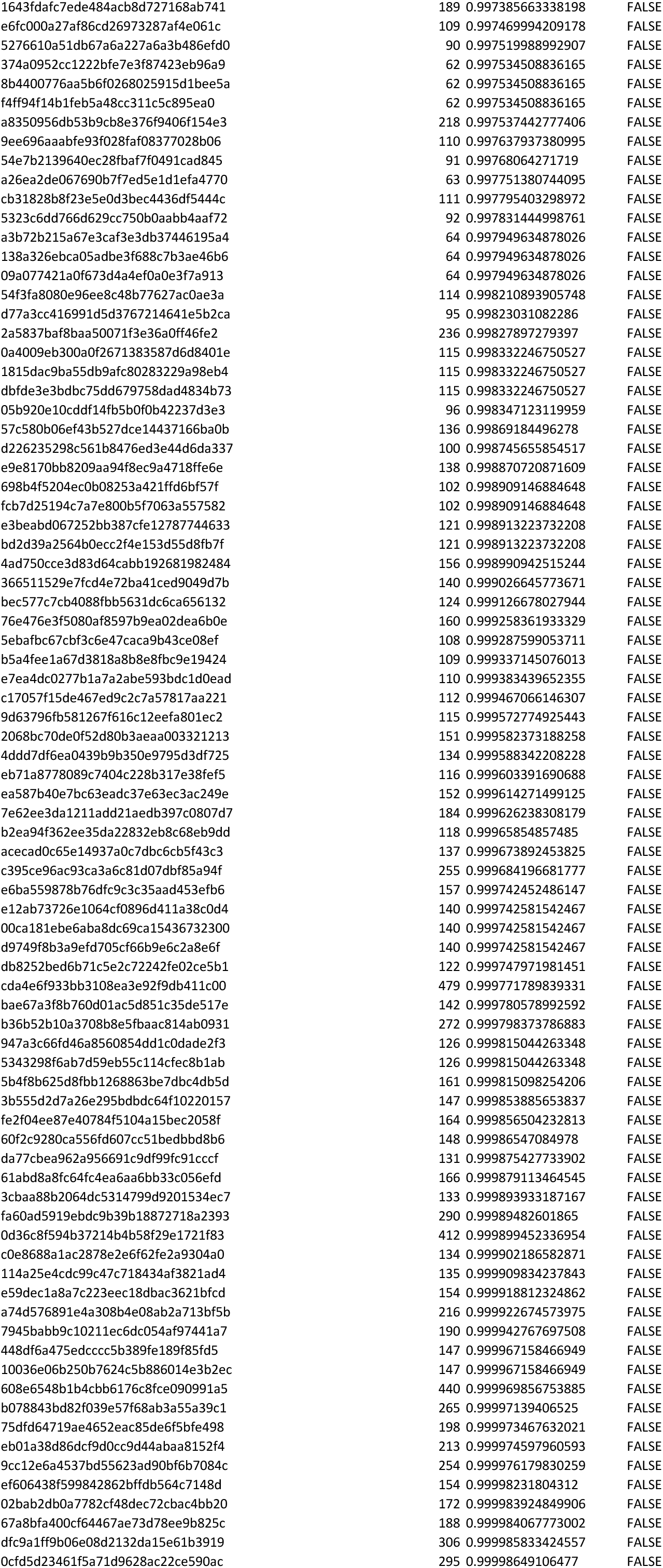
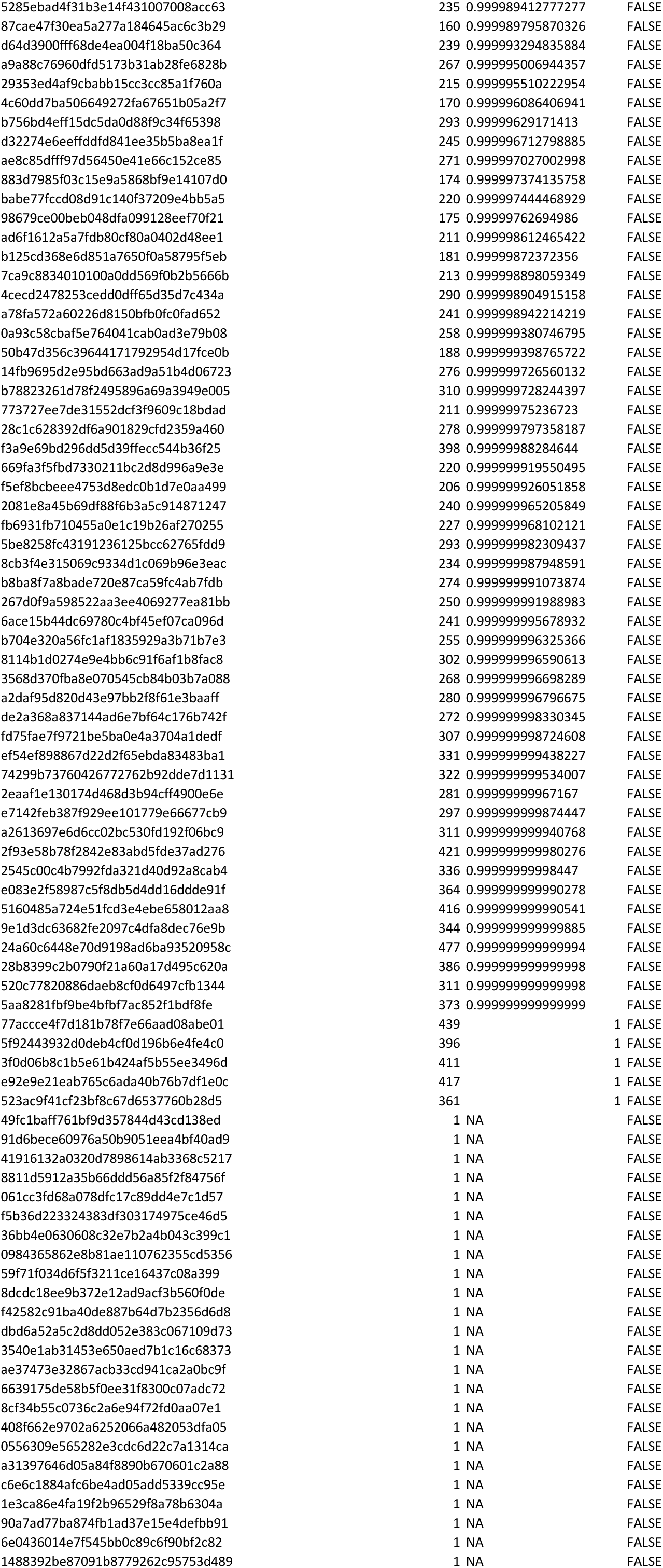
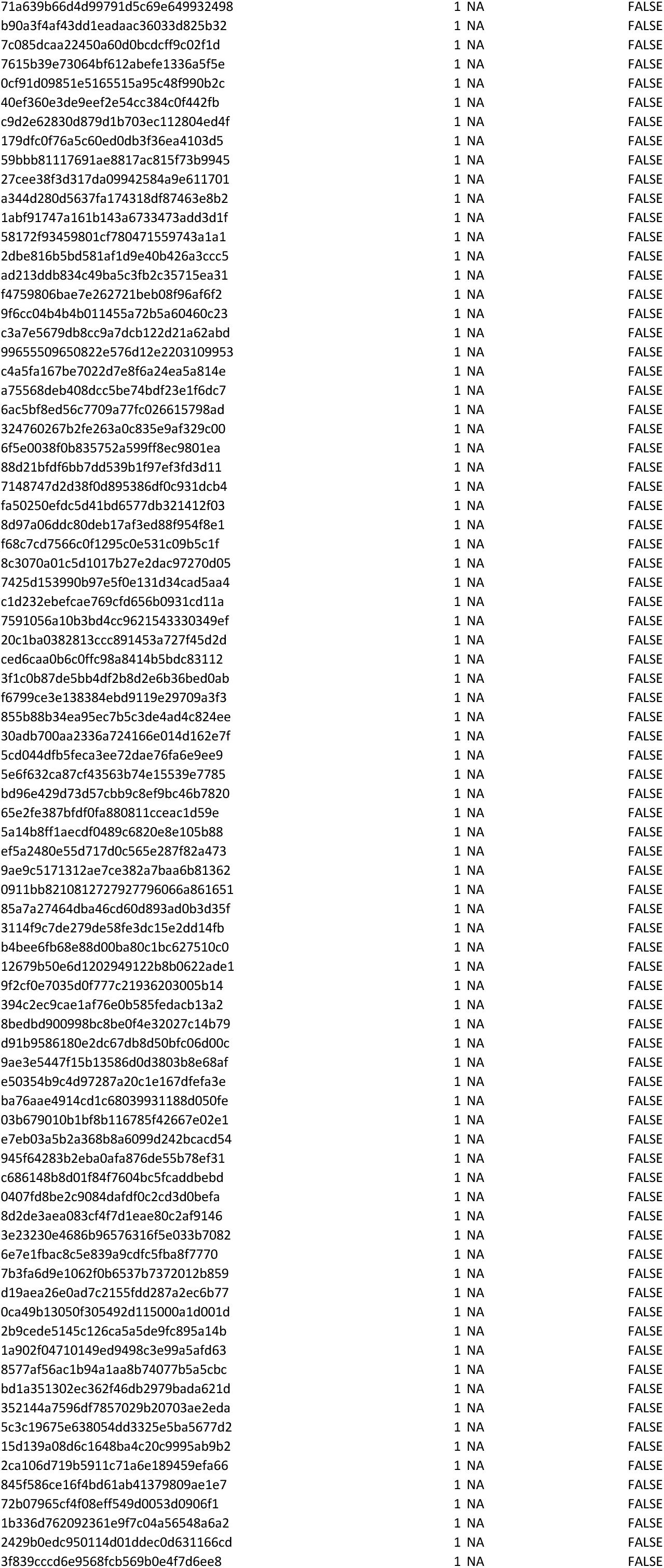
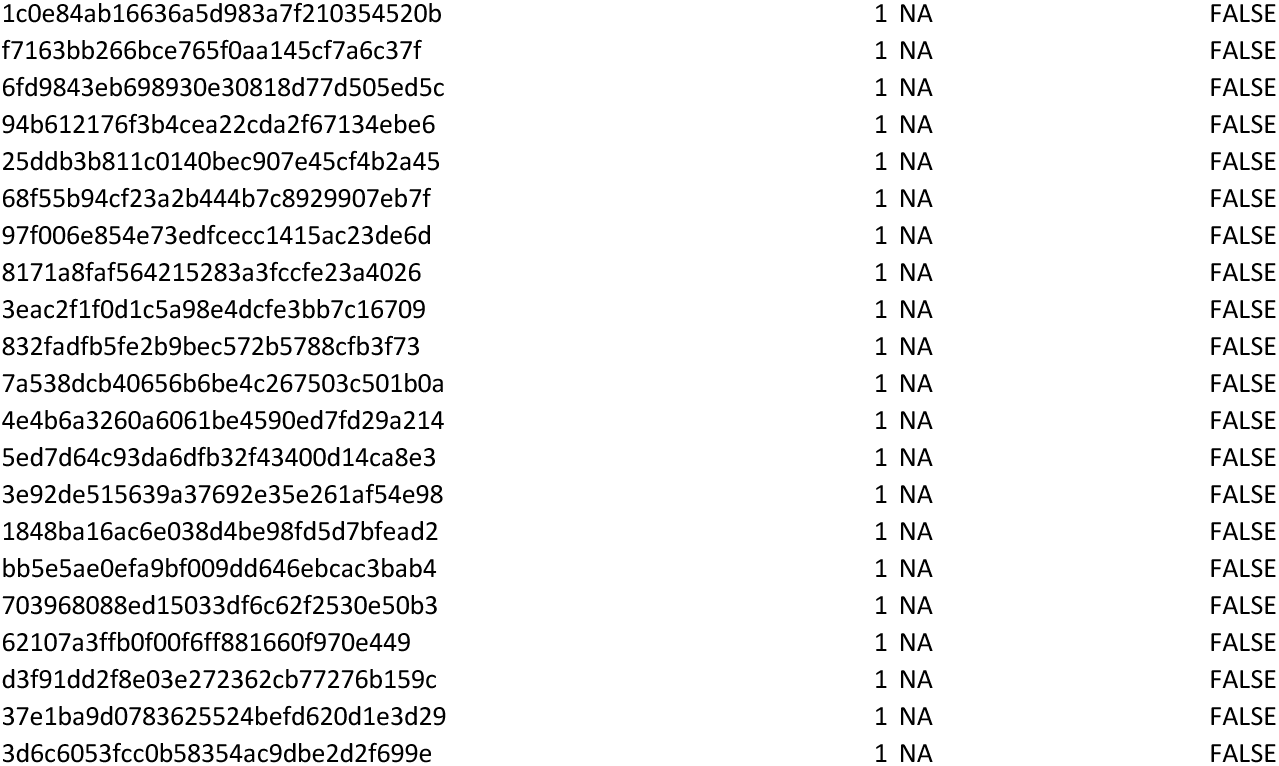
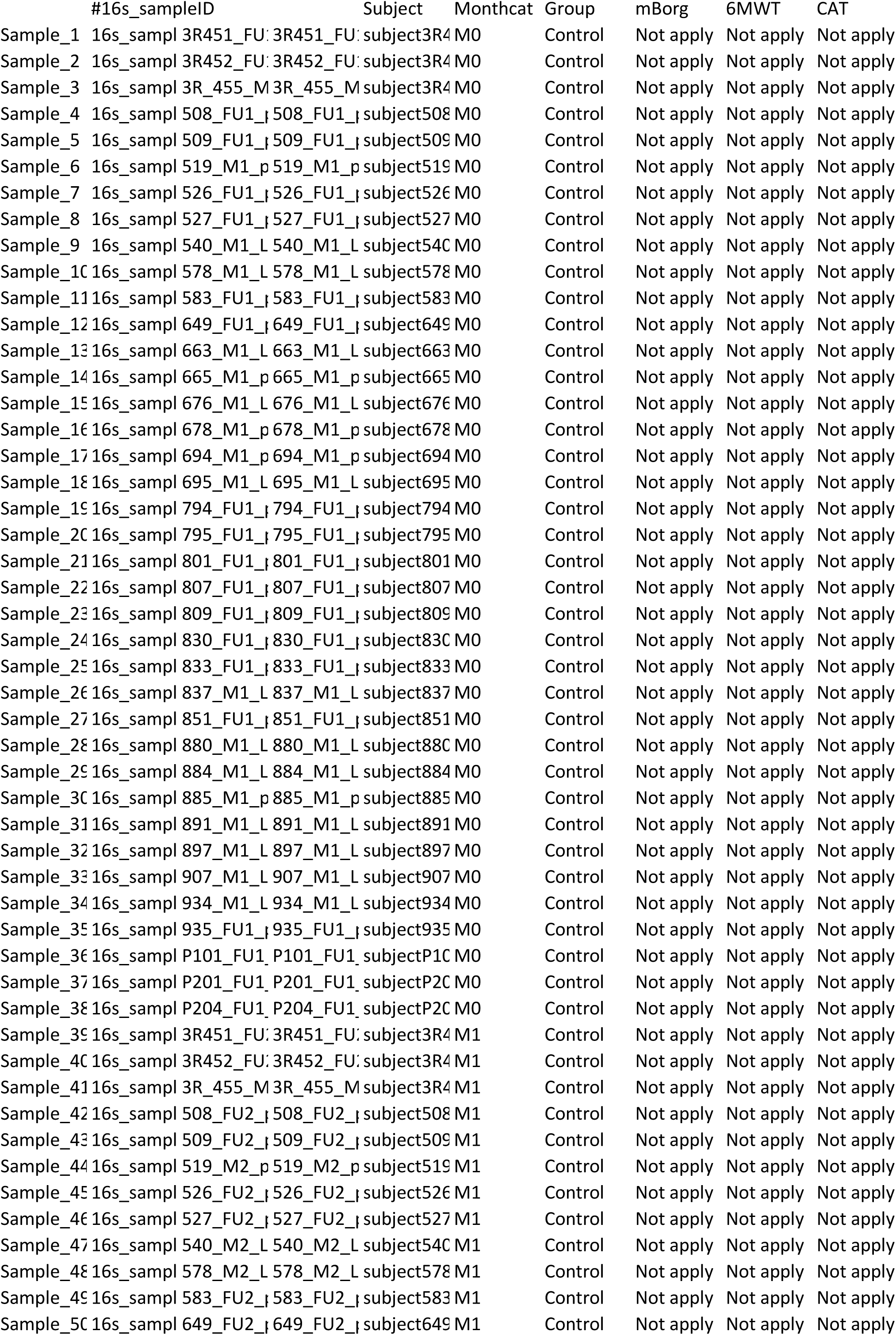
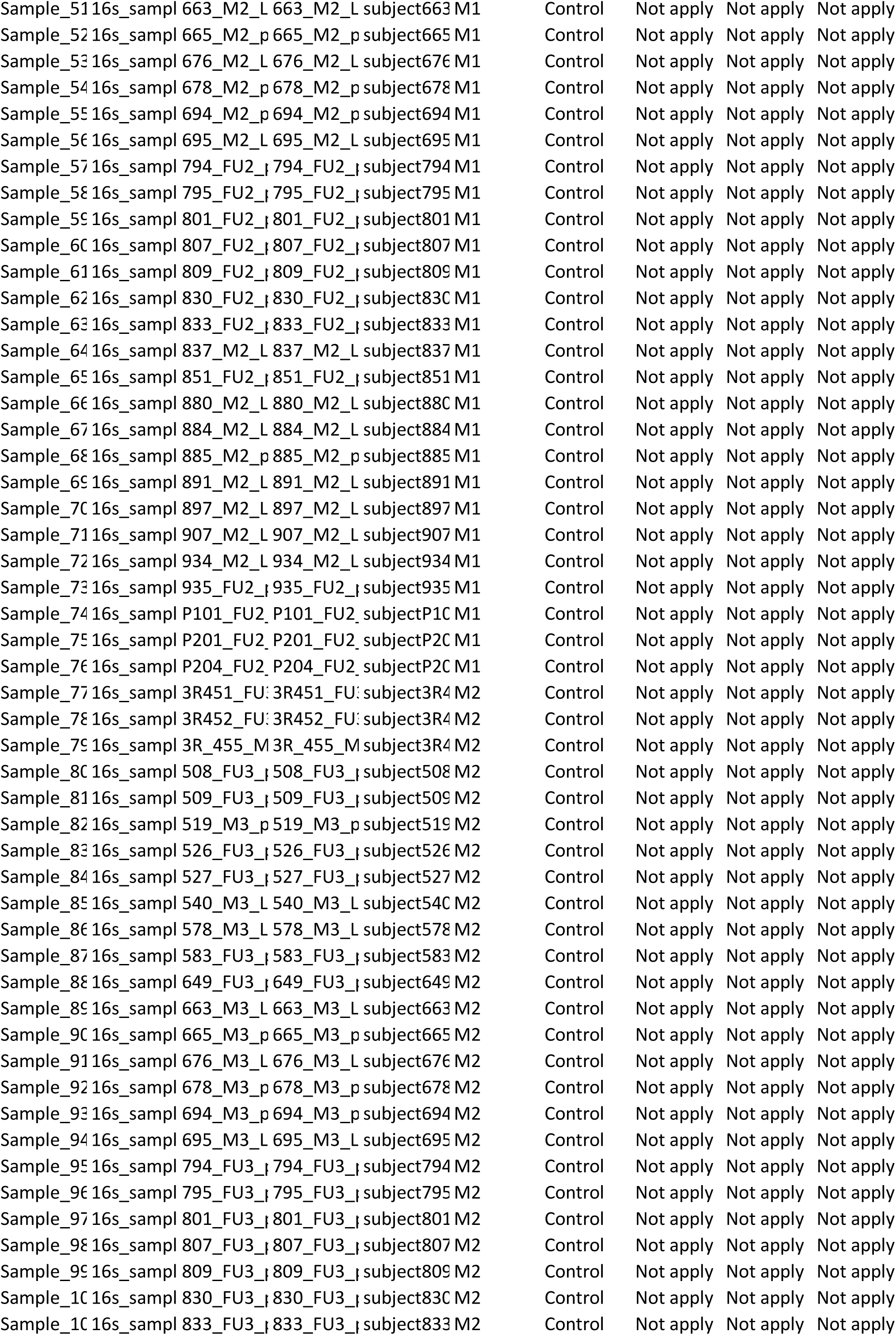
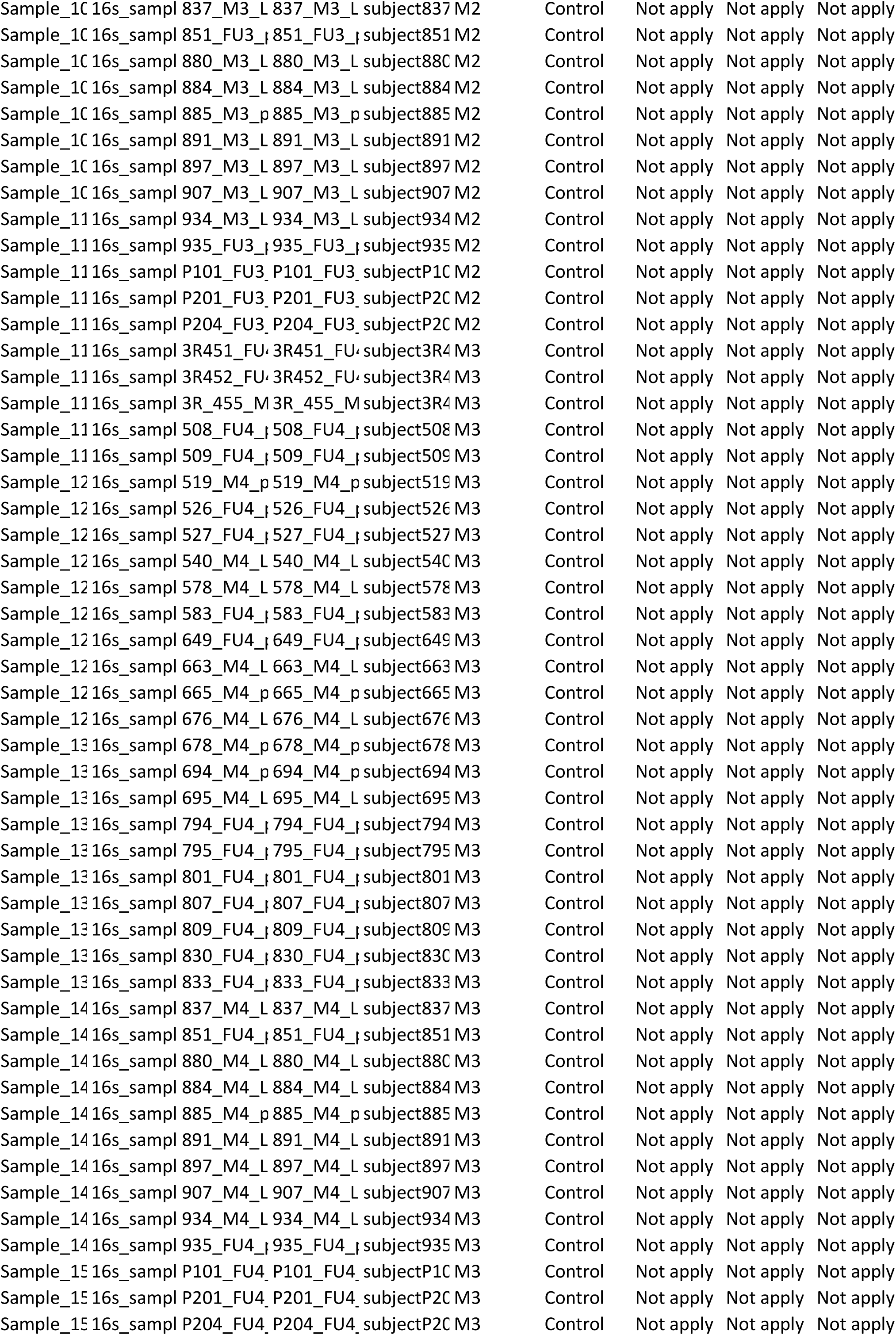
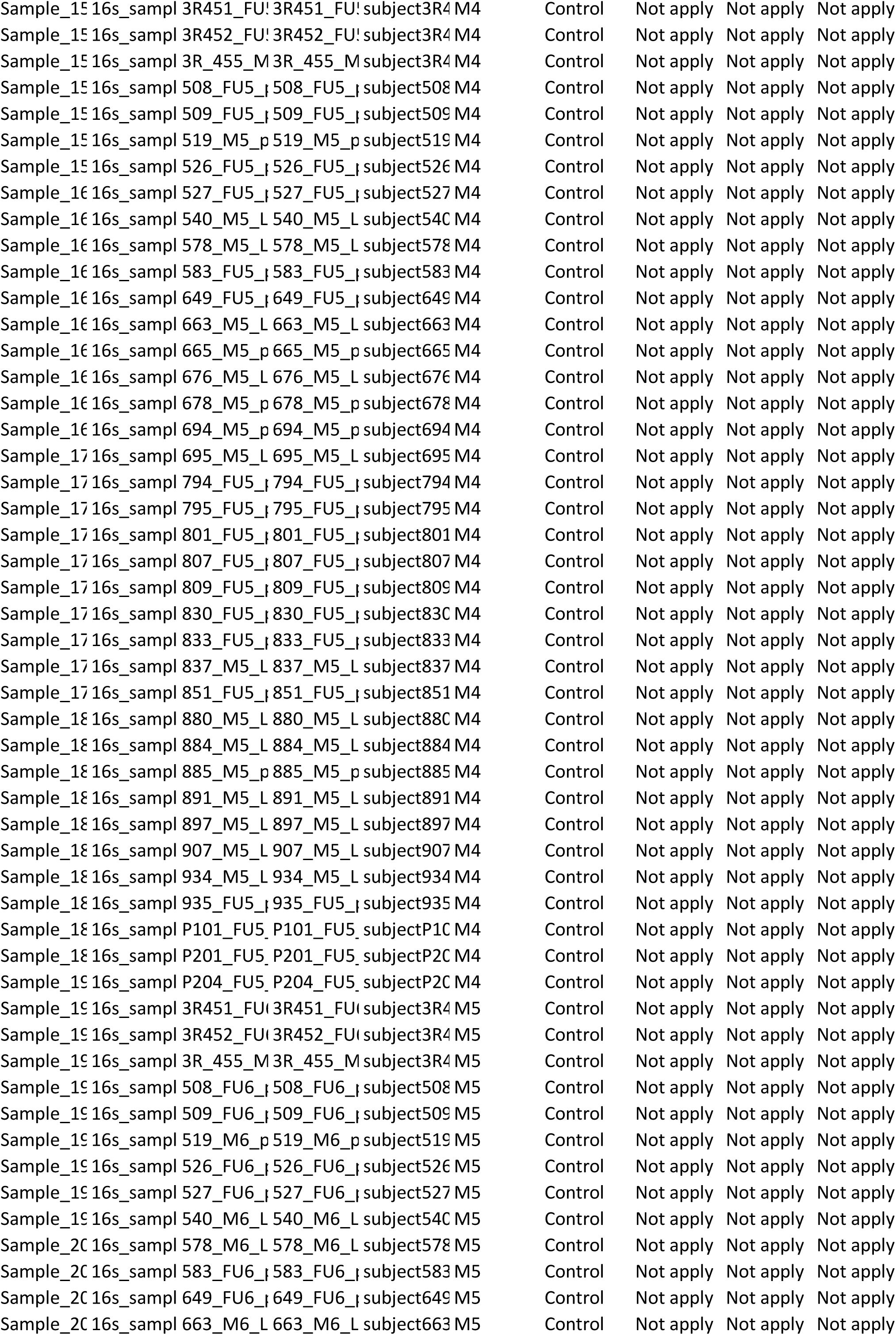
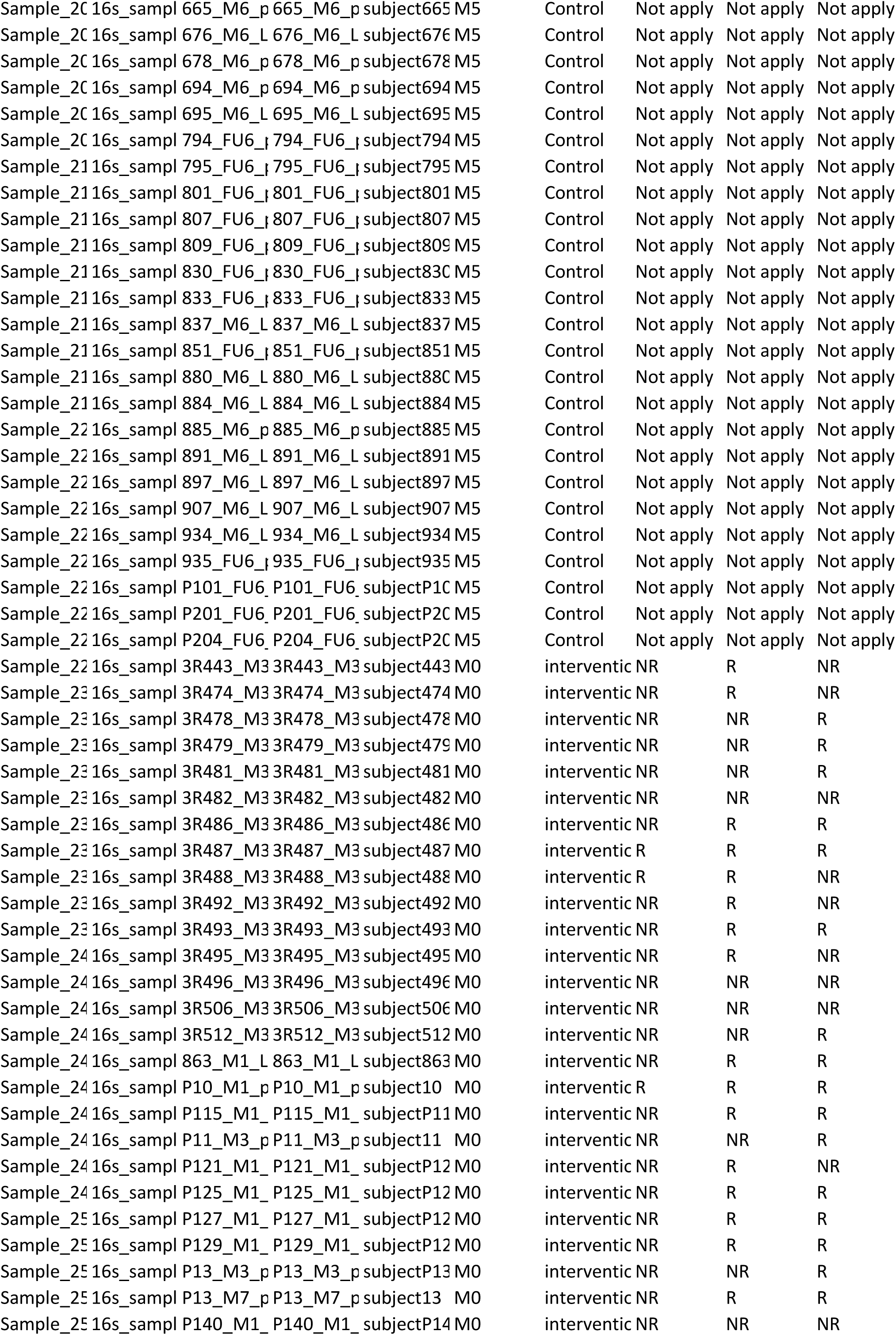
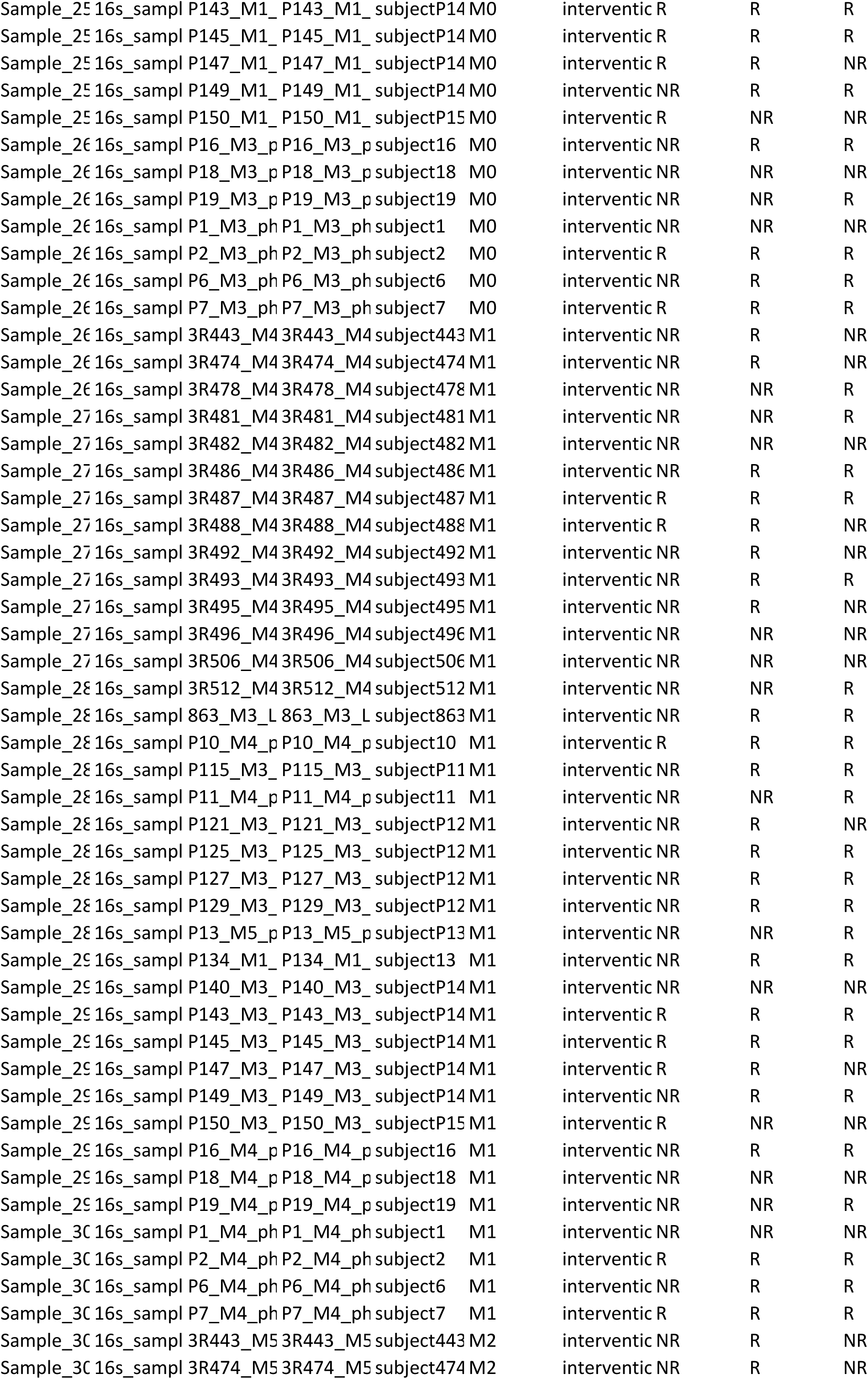
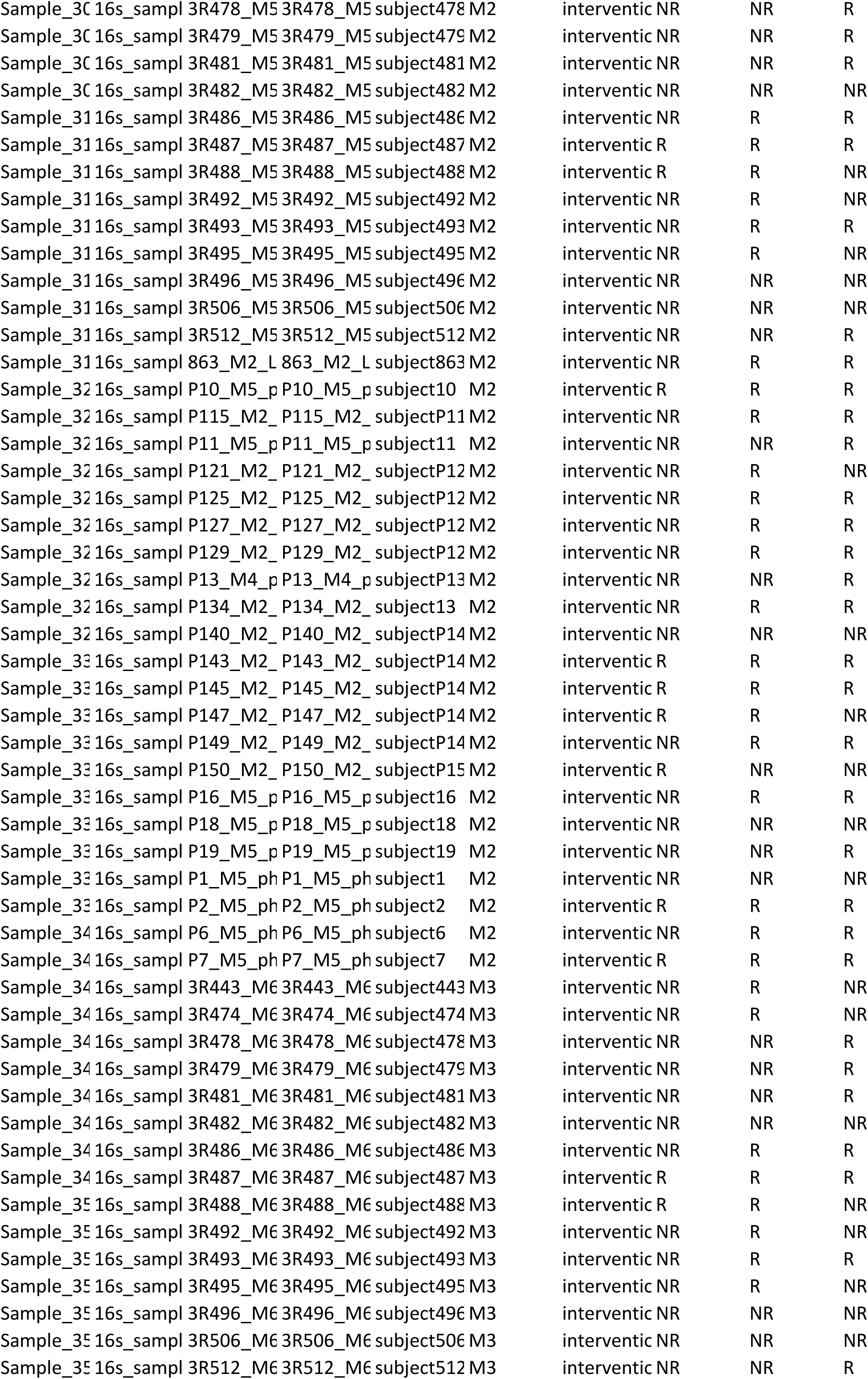
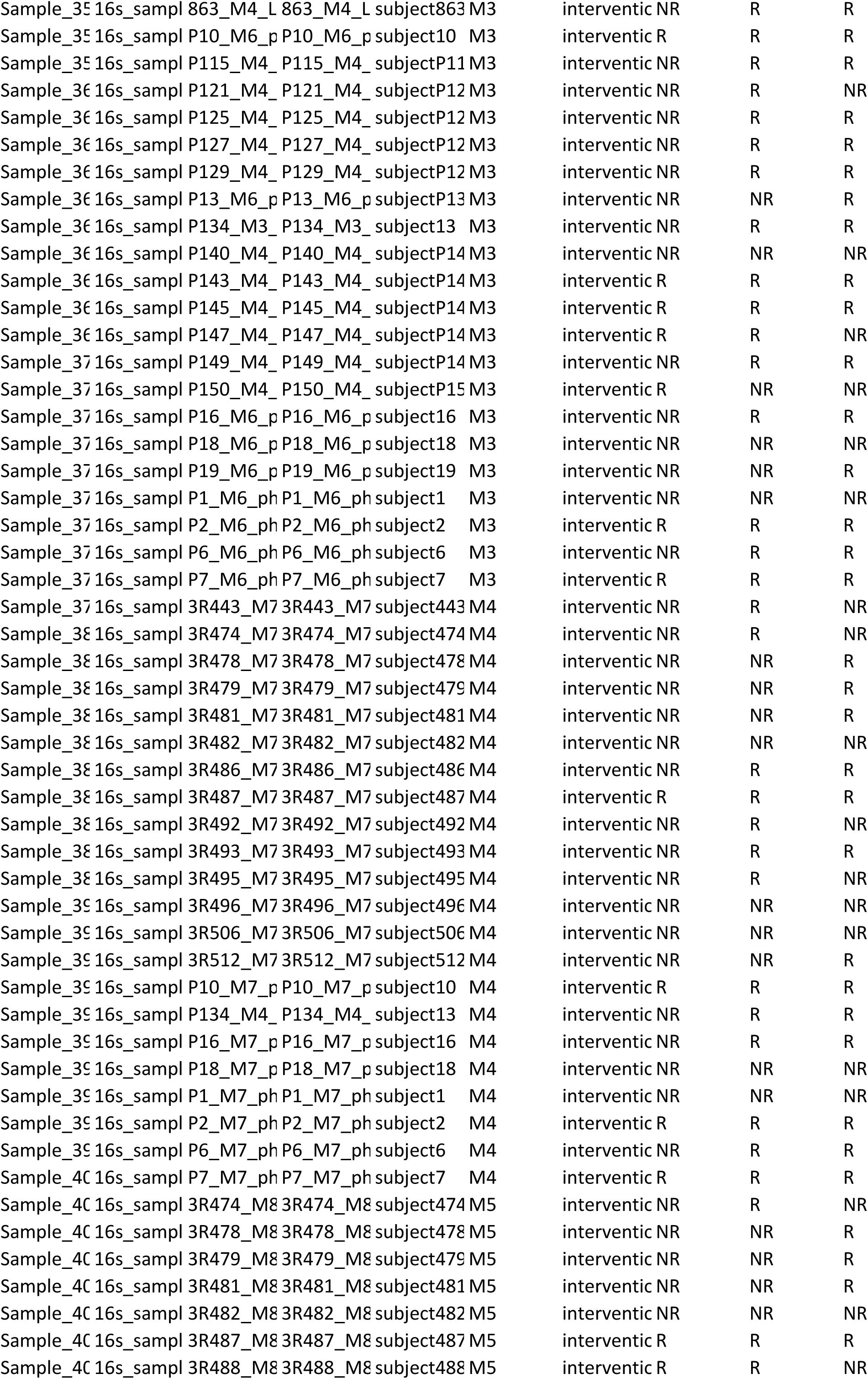
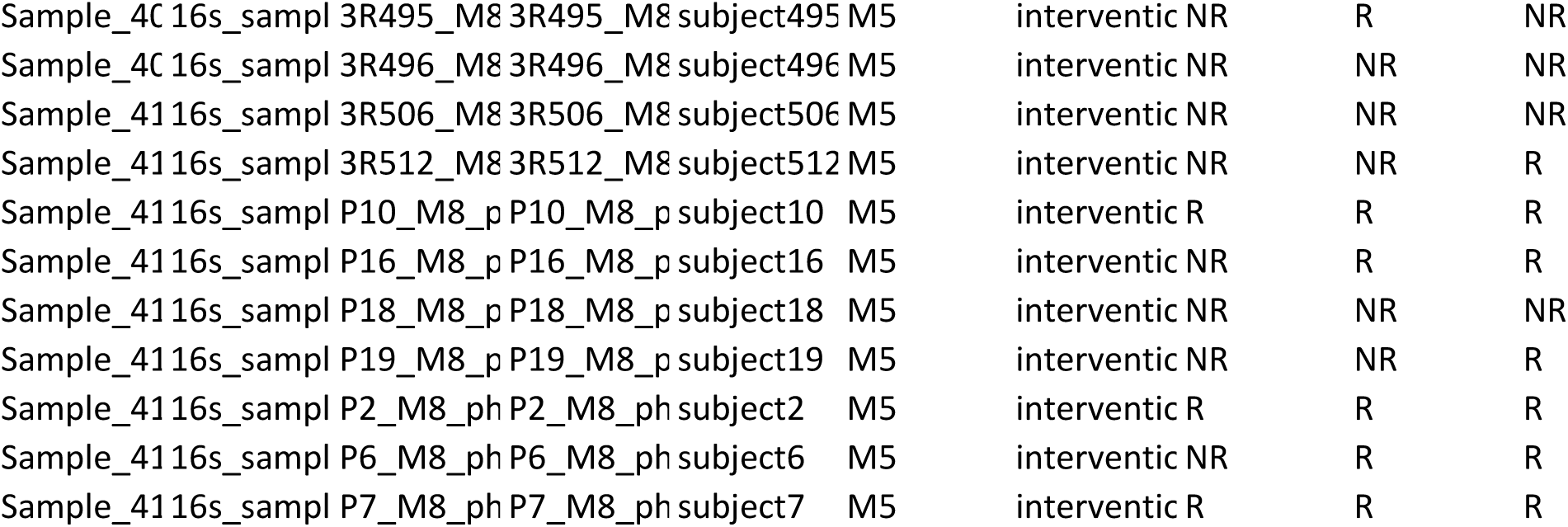

## Notes

**Funding:** This study is integrated in “PRIME – Pulmonary Rehabilitation and microbiota in exacerbations of COPD”, “GENIAL – Genetic and Clinical markers of COPD trajectory”, and “MicroAgeing - The role of microbiota in ageing”, funded by Programa Operacional de Competitividade e Internacionalização - COMPETE, through Fundo Europeu de Desenvolvimento Regional - FEDER (POCI-01-0145-FEDER-007628 and POCI-010145-FEDER-028806), Fundação para a Ciência e Tecnologia - FCT (PTDC/SAU-SER/28806/2017, PTDC/DTP-PIC/2284/2014 and PTDC/BIA-EVL/30212/2017) and under the project UIDB/04501/2020. S. Melo-Dias was supported by Grant SFRH/BD/140908/2018 from FCT. S. Souto-Miranda was supported by Grant SFRH/BD/146134/2019. A. Sousa was funded from national funds through FCT – Fundação para a Ciência e a Tecnologia, I.P., under the Scientific Employment Stimulus - Institutional Call - reference CEECINST/00026/2018.

### Competing Interest Statement

The authors have declared no competing interest.

### Clinical Protocols

https://pubmed.ncbi.nlm.nih.gov/31151409/

### Funding Statement

This study is integrated in PRIME, Pulmonary Rehabilitation and microbiota in exacerbations of COPD GENIAL, Genetic and Clinical markers of COPD trajectory and MicroAgeing, The role of microbiota in ageing, funded by Programa Operacional de Competitividade e Internacionalizacao, COMPETE, through Fundo Europeu de Desenvolvimento Regional - FEDER (POCI-010145-FEDER-028806 and POCI-01-0145-FEDER-007628), Fundacao para a Ciencia e Tecnologia - FCT (PTDC/DTP-PIC/2284/2014, PTDC/SAU-SER/28806/2017 and PTDC/BIA-EVL/30212/2017) and under the project UIDB/04501/2020. S. Melo-Dias was supported by Grant SFRH/BD/140908/2018 from FCT. A. Sousa was funded from national funds through FCT, Fundacao para a Ciencia e a Tecnologia, I.P., under the Scientific Employment Stimulus - Institutional Call - reference CEECINST/00026/2018.

### Author Declarations

A cross-sectional study was conducted. Ethical approvals were obtained from Administracao Regional de Saude Centro (64/2016) and from Centro Hospitalar do Baixo Vouga (08-03-17). Written informed consent was obtained from all participants.

### Summary of Updates

Abstract: Conclusions were rewritten Methods: a full description of how saliva samples were collected and handled was added

